## Supplementary material for "A handheld luminometer with sub-attomole limit of detection for distributed applications in global health": Firefly luciferase titration data

| Dilution number | Dilution factor | Transfer volume (uL) | Diluent volume (uL) | Total volume | Concentration (M) | Moles of enzyme | Result A | Err A | Result B | Err B | Luminometer samples |
|---|---|---|---|---|---|---|---|---|---|---|---|
| Stock | N/A | N/A | N/A | N/A | 1.6 E-4 | | N/A - saturated | N/A - saturated | N/A - saturated | N/A - saturated | N/A - saturated |
| 1 | 100 | 5 | 495 | 500 | 1.60E-06 | 2.40E-11 | N/A - saturated | N/A - saturated | N/A - saturated | N/A - saturated | N/A - saturated |
| 2 | 100 | 5 | 495 | 500 | 1.60E-08 | 2.40E-13 | 42725.29 | 98.19 | 41464 | 85.49 | 15 |
| 3 | 10 | 10 | 90 | 100 | 1.60E-09 | 2.40E-14 | 44865.96 | 84.22 | 42743.51 | 78.81 | 15 |
| 4 | 10 | 10 | 90 | 100 | 1.60E-10 | 2.40E-15 | 19260.89 | 36.43 | 18788.48 | 67.77 | 15 |
| 5 | 10 | 10 | 90 | 100 | 1.60E-11 | 2.40E-16 | 1856.8 | 3.12 | 1741.43 | 3.21 | 15 |
| 6 | 10 | 10 | 90 | 100 | 1.60E-12 | 2.40E-17 | 177.02 | 1.7 | 183.58 | 0.93 | 15 |
| 7 | 10 | 10 | 90 | 100 | 1.60E-13 | 2.40E-18 | 17.88 | 0.76 | 20.25 | 0.75 | 30 |
| 8 | 3 | 33.3 | 66.7 | 100 | 5.33E-14 | 8.00E-19 | 5.01 | 0.84 | 10.85 | 0.74 | 30 |
| 9 | 3 | 33.3 | 66.7 | 100 | 1.78E-14 | 2.67E-19 | 3.69 | 0.84 | 6.8 | 1.04 | 30 |
| 10 | 3 | 33.3 | 66.7 | 100 | 5.93E-15 | 8.89E-20 | -0.26 | 0.39 | 4.81 | 0.38 | 150 |
| 11 | 3 | 33.3 | 66.7 | 100 | 1.98E-15 | 2.96E-20 | 1.45 | 0.41 | 4.09 | 0.43 | 150 |

Enzyme volume (L) 1.50E-05

### Data summary

| | Channel A | Channel B | Average |
|---|---|---|---|
| **avg err_stdev** | 1.98 | 1.40 | 1.69 |
| **avg err_sem** | 0.55 | 0.62 | 0.58 |
| **Slope** | 8.03E+18 | 7.83E+18 | 7.93E+18 |
| **LOD_sem** | 2.04E-19 | 2.36E-19 | 2.20E-19 |
| **LOD_stdev** | 7.40E-19 | 5.38E-19 | 6.39E-19 |

| Dilution number | Dilution factor | Transfer volume (uL) | Diluent volume (uL) | Total volume (uL) | Concentration (M) | Moles of enzyme | Result A | Err A | Result B | Err B | Luminometer samples |
|---|---|---|---|---|---|---|---|---|---|---|---|
| Stock | N/A | N/A | N/A | N/A | 1.6 E-4 | | N/A | N/A | N/A | N/A | N/A |
| 1 | 100 | 5 | 495 | 500 | 1.60E-06 | 8.00E-12 | N/A | N/A | N/A | N/A | N/A |
| 2 | 100 | 5 | 495 | 500 | 1.60E-08 | 8.00E-14 | N/A | N/A | N/A | N/A | N/A |
| 3 | 10 | 10 | 90 | 100 | 1.60E-09 | 8.00E-15 | 45735 | 99.61 | 43566.53 | 89.33 | 15 |
| 4 | 10 | 10 | 90 | 100 | 1.60E-10 | 8.00E-16 | 15264.96 | 48.15 | 16829.5 | 59.99 | 15 |
| 5 | 10 | 10 | 90 | 100 | 1.60E-11 | 8.00E-17 | 1783.38 | 4.77 | 1654.43 | 4.38 | 15 |
| 6 | 10 | 10 | 90 | 100 | 1.60E-12 | 8.00E-18 | 183.07 | 1.08 | 171.24 | 0.94 | 15 |
| 7 | 10 | 10 | 90 | 100 | 1.60E-13 | 8.00E-19 | 17.68 | 0.73 | 21.32 | 0.85 | 30 |
| 8 | 3 | 33.3 | 66.7 | 100 | 5.33E-14 | 2.67E-19 | 6.62 | 1.02 | 12.18 | 0.77 | 30 |
| 9 | 3 | 33.3 | 66.7 | 100 | 1.78E-14 | 8.89E-20 | 4.2 | 0.92 | 9.04 | 0.91 | 30 |
| 10 | 3 | 33.3 | 66.7 | 100 | 5.93E-15 | 2.96E-20 | 1.7 | 1.05 | 7.25 | 0.81 | 30 |
| 11 | 3 | 33.3 | 66.7 | 100 | 1.98E-15 | 9.88E-21 | 2.23 | 0.41 | 7.31 | 0.43 | 30 |
| 12 | 3 | 33.3 | 66.7 | 100 | 6.58E-16 | 3.29E-21 | 3.34 | 0.97 | 4.59 | 0.99 | 30 |
| 13 | 3 | 33.3 | 66.7 | 100 | 2.19E-16 | 1.10E-21 | 3.05 | 1.12 | 7.66 | 0.86 | 30 |

| | |
|---|---|
| Measurement volume (L) | 3.00E-05 |
| Enzyme volume (L) | 5.00E-06 |
| Assay buffer volume(L) | 1.50E-05 |

### Data summary

| | Channel A | Channel B | Average |
|---|---|---|---|
| **avg err_stdev** | 0.75 | 1.42 | 1.09 |
| **avg err_sem** | 0.89 | 0.77 | 0.83 |
| **Slope** | 1.91E+19 | 2.10E+19 | 2.01E+19 |
| **LOD_sem** | 1.40E-19 | 1.10E-19 | 1.25E-19 |
| **LOD_stdev** | 1.18E-19 | 2.02E-19 | **1.60E-19** |
