## Supplementary material for "A handheld luminometer with sub-attomole limit of detection for distributed applications in global health": Fleet calibration data in whole blood

### Luminometer ID

Temp = 20 C

| Concentration (fM) | Moles of enzyme | 1 A | 1 err | 1 B | 1 err | 2 A | 2 err | 2 B | 2 err | 4 A | 4 err | 4 B | 4 err | 5 A | 5 err | 5 B | 5 err |
|---|---|---|---|---|---|---|---|---|---|---|---|---|---|---|---|---|---|
| 10000 | 1.31E-15 | 9968.8 | 17.7 | 8253.8 | 15.0 | 9681.0 | 17.3 | 9130.7 | 16.6 | 9816.7 | 17.2 | 9832.9 | 17.1 | 10036.9 | 18.1 | 9701.9 | 17.7 |
| 1000 | 1.31E-16 | 948.1 | 1.9 | 928.8 | 2.5 | 948.4 | 2.7 | 957.5 | 2.6 | 952.6 | 2.7 | 952.4 | 2.6 | 958.3 | 2.2 | 946.6 | 3.3 |
| 100 | 1.31E-17 | 105.6 | 1.1 | 99.2 | 1.3 | 100.3 | 1.5 | 102.6 | 0.8 | 101.3 | 1.3 | 100.1 | 0.8 | 97.5 | 1.2 | 97.5 | 2.3 |
| 30 | 3.94E-18 | 38.4 | 1.0 | 37.7 | 1.0 | 36.2 | 0.8 | 37.4 | 0.9 | 35.3 | 0.7 | 37.4 | 0.8 | 35.5 | 1.0 | 40.7 | 1.4 |
| 9 | 1.18E-18 | 18.6 | 1.1 | 17.9 | 0.8 | 14.3 | 0.7 | 18.9 | 0.9 | 15.7 | 0.8 | 17.2 | 0.9 | 16.5 | 1.1 | 14.2 | 1.3 |
| 2.7 | 3.54E-19 | 11.3 | 0.9 | 11.2 | 0.8 | 9.6 | 0.9 | 12.1 | 0.8 | 10.0 | 0.9 | 10.0 | 0.7 | 10.7 | 1.1 | 13.7 | 1.8 |
| 0.81 | 1.06E-19 | 9.7 | 0.7 | 12.4 | 0.6 | 5.9 | 0.8 | 10.9 | 0.9 | 9.9 | 0.7 | 9.6 | 0.9 | 10.1 | 1.1 | 11.3 | 0.8 |
| 0.243 | 3.19E-20 | 10.3 | 0.4 | 9.7 | 0.4 | 7.3 | 0.4 | 10.0 | 0.4 | 8.2 | 0.3 | 9.2 | 0.3 | 8.3 | 0.4 | 9.8 | 0.8 |

| Concentration (fM) | Moles of enzyme | 6 A | 6 err | 6 B | 6 err | 7 A | 7 err | 7 B | 7 err | 8 A | 8 err | 8 B | 8 err | 9 A | 9 err | 9 B | 9 err | 10 A | 10 err | 10 B | 10 err |
|---|---|---|---|---|---|---|---|---|---|---|---|---|---|---|---|---|---|---|---|---|---|
| 10000 | 1.31E-15 | 8908.5 | 18.7 | 9362.5 | 19.9 | 10293.2 | 22.1 | 10549.6 | 22.6 | 9155.3 | 1646.8 | 9977.6 | 20.6 | 10121.7 | 20.5 | 10143.6 | 19.6 | 10270.4 | 21.1 | 10560.8 | 23.8 |
| 1000 | 1.31E-16 | 987.4 | 3.0 | 988.6 | 2.4 | 997.5 | 2.9 | 991.8 | 2.7 | 1424.5 | 176.2 | 989.2 | 1.9 | 992.3 | 2.9 | 981.1 | 2.2 | 1001.0 | 2.5 | 1002.2 | 2.9 |
| 100 | 1.31E-17 | 108.1 | 1.3 | 110.6 | 1.4 | 100.0 | 1.3 | 103.2 | 1.3 | -46.3 | 17.7 | 109.4 | 10.0 | 83.5 | 1.1 | 104.3 | 1.0 | 110.7 | 2.4 | 110.6 | 1.1 |
| 30 | 3.94E-18 | 36.1 | 1.0 | 38.4 | 0.8 | 30.8 | 1.2 | 32.4 | 1.2 | -51.2 | 10.9 | 35.7 | 1.0 | 40.4 | 0.8 | 41.9 | 0.9 | 37.5 | 1.0 | 39.9 | 0.9 |
| 9 | 1.18E-18 | 16.2 | 0.7 | 20.5 | 0.8 | 11.7 | 1.0 | 14.4 | 1.4 | -97.7 | 5.4 | 16.3 | 0.9 | 20.7 | 1.9 | 22.0 | 0.8 | 19.3 | 1.2 | 18.4 | 0.8 |
| 2.7 | 3.54E-19 | 7.8 | 0.9 | 12.1 | 0.9 | 4.6 | 1.5 | 10.9 | 2.0 | -73.0 | 7.5 | 13.1 | 1.1 | 15.8 | 0.9 | 16.1 | 0.7 | 11.1 | 1.0 | 12.6 | 0.8 |
| 0.81 | 1.06E-19 | 8.4 | 0.7 | 9.8 | 0.7 | 1.8 | 1.4 | 5.0 | 1.1 | -56.5 | 7.6 | 9.9 | 0.7 | 11.2 | 0.9 | 13.9 | 0.9 | 11.5 | 0.8 | 10.5 | 0.8 |
| 0.243 | 3.19E-20 | 8.4 | 0.4 | 9.4 | 0.3 | 2.1 | 0.7 | 4.8 | 0.6 | -48.9 | 3.3 | 8.6 | 0.4 | 12.0 | 0.4 | 13.1 | 0.3 | 9.8 | 0.6 | 8.0 | 0.4 |

*The shutter on 8A was malfunctioning during this dataset

| | | |
|---|---|---|
| Enzyme diluted in PBSTB (PBS + BSA + tween + whole blood) | | |
| Enzyme master mix volume per point | 1.05E-03 | Mean |
| Samples per point | 8.00E+00 | Std |
| Diluted enzyme volume per sample (L) | 1.31E-04 | CV (%) |

#### Data summary

| | Channel A | Channel B | Average |
|---|---|---|---|
| **avg err_stdev** | 1.87 | 1.04 | 1.45 |
| **avg err_sem** | 0.68 | 0.67 | 0.68 |
| **Slope** | 7.23E+18 | 7.29E+18 | 7.26E+18 |
| **LOD_sem** | 2.84E-19 | 2.76E-19 | 2.80E-19 |
| **LOD_stdev** | 7.78E-19 | 4.26E-19 | 6.02E-19 |
