## Supplementary material for "A handheld luminometer with sub-attomole limit of detection for distributed applications in global health": Bill of materials

### Assembly items

| Description | Internal part | Vendor part | Vendor | Fabrication method | Quantity per | Pack Price | Pack size | Total per |
|---|---|---|---|---|---|---|---|---|
| **Analog PCB assembly - itemized below** | **5-2002** | | **Assembled in house** | Reflow in house | **1** | **$1.00** | **1** | **$267.33** |
| **Digital PCB assembly - itemized below** | **5-2003** | | **Assembled in house** | Reflow in house | **1** | **$1.00** | **1** | **$88.18** |
| Li-Poly battery 4400 mA-hrs | | 354 | Adafruit | Used as purchased | 1 | $19.95 | 1 | $19.95 |
| PMIC board | | 2465 | Adafruit | Used as purchased | 1 | $19.95 | 1 | $19.95 |
| Hxchen 1.5V-4.5V 7500RPM Coreless Micro DC Motor 6x10mm | | B07ST8FML4 | Amazon | Used as purchased | 2 | $7.69 | 5 | $3.08 |
| Raspberry Pi Zero W | | Zero W | Canakit | Used as purchased | 1 | $10.00 | 1 | $10.00 |
| 40-pin receptacle, extra long | | SAM12900-ND | Digikey | Used as purchased | 1 | $104.33 | 10 | $10.43 |
| Tube holder assembly | 2-0143 | Black resin | Formlabs | 3D printed in house* | 1 | $10.00 | 1 | $10.00 |
| 1/10" dia x 1/16" thick magnet with 0.31 lb pull force | | DH11 | K&J Magnetics | Used as purchased | 14 | $120.00 | 150 | $11.20 |
| 18-8 SS dowel pin 1/16" dia x 1.25" long | | 90145A425 | McMaster-Carr | Used as purchased | 1 | $9.52 | 50 | $0.19 |
| Main enclosure - cost for all external 3D printed parts lumped in this line | 3-0682 | | Hawkridge | Multijet fusion 3D printing | 1 | $433.36 | 10 | $43.34 |
| Rear access lid - cost lumped above | 3-0684 | | Hawkridge | Multijet fusion 3D printing | 1 | $0.00 | 10 | $0.00 |
| Front / side panel - cost lumped above | 3-0682 | | Hawkridge | Multijet fusion 3D printing | 1 | $0.00 | 10 | $0.00 |
| Battery clip - cost lumped above | 3-0953 | | Hawkridge | Multijet fusion 3D printing | 1 | $0.00 | 10 | $0.00 |
| 1" x 1/8" x 1/16" (LxWxT) magnet with 2.31 lb pull force | | BX021 | K&J Magnetics | Used as purchased | 8 | $45.00 | 100 | $3.60 |
| A foam gasket between the main enclosure front face and the Pimoroni inky screen for the ULC luminometer* | 3-0883 | 8647K102 | McMaster-Carr | Laser-cut in house* | 1 | $1.00 | 1 | $1.00 |
| Motor spacer - laser cut acrylic | 3-0892 | 8492K133 | McMaster-Carr | Laser-cut in house* | 1 | $1.00 | 1 | $1.00 |
| Inter-PCB standoffs | | 94868A163 | McMaster-Carr | Used as purchased | 4 | $1.00 | 1 | $4.00 |
| Button head M2.5 | | 92095A113 | McMaster-Carr | Used as purchased | 5 | $14.80 | 100 | $0.74 |
| Hex socket head cap screw M2.5x0.45 x 14 Stainless Steel | | 91292A017 | McMaster-Carr | Used as purchased | 1 | $4.63 | 50 | $0.09 |
| PH Countersunk flat head screw M2.5x0.45 x 8 Stainless Steel | | 92125A086 | McMaster-Carr | Used as purchased | 3 | $6.70 | 25 | $0.80 |
| Roller bearings | | 7804K119 | McMaster-Carr | Used as purchased | 4 | $7.03 | 1 | $28.12 |
| Shutter shaft screw | | 92290A850 | McMaster-Carr | Used as purchased | 2 | $12.18 | 50 | $0.49 |
| M2 thin brass nut | | 93187A110 | McMaster-Carr | Used as purchased | 2 | $7.21 | 50 | $0.29 |
| Stainless steel thread-forming screw for brittle plastics, round head, Phillips drive, #4 - 3/8" long. Drill diameter: 0.086" | | 97349A100 | McMaster-Carr | Used as purchased | 3 | $5.67 | 100 | $0.17 |
| M2 SS Nylon insert locknut, pack of 10 | | 93625A101 | McMaster-Carr | Used as purchased | 2 | $7.70 | 10 | $1.54 |
| Stainless steel thread-forming screw for plastics, flat head, Torx drive, #2 - 1/4" long. Drill diameter: 0.076" | | 95893A550 | McMaster-Carr | Used as purchased | 4 | $16.64 | 50 | $1.33 |
| Hex socket head cap screw M2x0.40 x 15 Stainless Steel | | 91292A335 | McMaster-Carr | Used as purchased | 2 | $13.34 | 5 | $5.34 |
| Hex socket head cap screw M2x0.40 x 6 Stainless Steel | | 91292A831 | McMaster-Carr | Used as purchased | 2 | $9.73 | 100 | $0.19 |
| Chamfered plain washer normal M2 Stainless Steel | | 98689A110 | McMaster-Carr | Used as purchased | 6 | $2.86 | 100 | $0.17 |
| Stainless steel thread-forming screw for plastics, flat head, Torx drive, #0 - 3/16" long. Drill diameter: 0.0465" | | 95893A501 | McMaster-Carr | Used as purchased | 6 | $17.40 | 50 | $2.09 |
| Hex socket head cap screw M2.5x0.45 x 10 Stainless Steel | | 91292A014 | McMaster-Carr | Used as purchased | 2 | $6.54 | 100 | $0.13 |
| Male-Female Threaded Hex Standoff, 18-8 Stainless Steel, 4.500 mm Hex, 16 mm Long, M2.5 x 0.45 mm Thread | | 93655A353 | McMaster-Carr | Used as purchased | 2 | $3.72 | 1 | $7.44 |
| M2.5 Stainless steel washer | | 98689A111 | McMaster-Carr | Used as purchased | 7 | $2.86 | 100 | $0.20 |
| Male-Female Threaded Hex Standoff, 18-8 Stainless Steel, 4.500 mm Hex, 8 mm Long, M2.5 x 0.45 mm Thread | | 93655A094 | McMaster-Carr | Used as purchased | 2 | $3.16 | 1 | $6.32 |
| Shutter flag | 3-0717 | | OSH Stencil | Laser-cut steel | 2 | $65.49 | 20 | $6.55 |
| Pimoroni PIM368 Inkyphat eInk Screen B&W | | PIM368 | Pimoroni | Used as purchased | 1 | $24.30 | 1 | $24.30 |
| Aluminum shaft spacer | 3-0716 | | Protolabs | CNC Machined | 1 | $385.00 | 10 | $38.50 |
| Aluminum motor mount | 3-1003 | | Protolabs | CNC Machined | 1 | $574.00 | 10 | $57.40 |
| Follower gear for the luminometer shutter | 3-0889 | M46S32CU | Ultimaker | 3D printed in house* | 2 | $1.00 | 1 | $2.00 |
| Drive gear for the luminometer shutter | 3-0890 | M46S32CU | Ultimaker | 3D printed in house* | 2 | $1.00 | 1 | $2.00 |
| PMIC cover panel | 3-0984 | 8685K41 | McMaster-Carr | Laser-cut in house* | 1 | $1.00 | 1 | $1.00 |
| RPi 40-pin receptacle spacer | 3-0811 | 8560K359 | McMaster-Carr | Laser-cut in house* | 1 | $1.00 | 1 | $1.00 |
| Total ground paint for all 3D printed parts | | B01N01RNFS | Amazon | Sprayed onto parts | 0.1 | $46.00 | 1 | $4.60 |
| Black 3.0 paint for critical surfaces | | Black 3.0 | Culture hustle USA | Sprayed onto parts | 0.1 | $29.99 | 1 | $3.00 |
| Loctite 420 superglue | | 66745A21 | McMaster-Carr | Applied in small quantities | 0.05 | $31.00 | 1 | $1.55 |

| | | | | | | | | |
|---|---|---|---|---|---|---|---|---|
| Reflective mylar film for sample cavities | 3-0710 | B018VI75CS | Amazon | Laser-cut in house* | 0.01 | $30.99 | 1 | $0.31 |
| Dow Corning silicone sealant | | 7425A5 | McMaster-Carr | Applied in small quantities | 0.01 | $11.85 | 1 | $0.12 |
| Light-proofing gasket behind buttons | 3-0825 | 86435K45 | McMaster-Carr | Laser-cut in house* | 0.01 | $55.31 | 1 | $0.55 |
| Light-proofing gasket between PCB and tube holder | 3-0679 | 1460N22 | McMaster-Carr | Laser-cut in house* | 0.1 | $29.43 | 1 | $2.94 |
| *Quantity per device was crudely approximated | | | | | | | | |
| **Total cost per luminometer** | | | | | | | | **$694.53** |

#### Digital PCB - itemized

| Description | References | Value | Part Number | Vendor | Quantity Per PCB | Pack Price | Pack size | Total cost per PCB |
|---|---|---|---|---|---|---|---|---|
| Buzzer | BZ1 | 668-SMI-1027-T-5V-RCT-ND | 668-SMI-1027-T-5V-RCT-ND | Digikey | 1 | $15.42 | 10 | $1.54 |
| Unpolarized capacitor | C1 C4 C5 C7 C9 C10 C11 | 10nF | 478-12682-1-ND | Digikey | 7 | $2.28 | 100 | $0.16 |
| Unpolarized capacitor | C6 C13 | 100nF | 399-20424-1-ND | Digikey | 2 | $5.30 | 10 | $1.06 |
| Unpolarized capacitor | C2 | 2.2uF | 445-5157-1-ND | Digikey | 1 | $1.14 | 10 | $0.11 |
| Unpolarized capacitor | C8 C12 | 10uF | 1276-1948-1-ND | Digikey | 2 | $5.15 | 10 | $1.03 |
| Unpolarized capacitor | C3 | 47uF | 478-11180-1-ND | Digikey | 1 | $8.42 | 10 | $0.84 |
| Ferrite bead, small symbol | FB1 FB2 FB3 | Ferrite_Bead_Small | 399-Z0603C600BPWZTCT-ND | Digikey | 3 | $5.48 | 50 | $0.33 |
| Generic connector, single row, 01x02, script generated (kicad-library-utils/schlib/autogen/connector/) | J2 J3 J5 J6 J8 | H2191-ND | H2191-ND | Digikey | 5 | $2.55 | 10 | $1.28 |
| Connector | J7 | Harting_15110122601000 | 1195-3359-1-ND | Digikey | 1 | $37.75 | 10 | $3.78 |
| Generic connector, single row, 01x08, script generated (kicad-library-utils/schlib/autogen/connector/) | J1 | PMIC_header_right_angle | 732-5341-ND | Digikey | 1 | $6.68 | 10 | $0.67 |
| Inductor | L1 L2 | 732-4872-1-ND | 732-4872-1-ND | Digikey | 2 | $16.48 | 10 | $3.30 |
| MOSFET N-CH 30V 5A SOT23 | Q1 Q2 Q3 | IRLML6344TRPBF | IRLML6344TRPBFCT-ND | Digikey | 3 | $4.76 | 10 | $1.43 |
| Resistor | R7 R8 | 200m | 2019-SR731JTTDR200FCT-ND | Digikey | 2 | $8.37 | 25 | $0.67 |
| Resistor | R1 R6 | 180R | 311-180DCT-ND | Digikey | 2 | $1.51 | 10 | $0.30 |
| Resistor | R2 R3 R9 R10 R11 | 10k | YAG2321CT-ND | Digikey | 5 | $0.59 | 10 | $0.30 |
| Switch | SW4 | AS11CH | 360-2728-ND | Digikey | 1 | $47.39 | 10 | $4.74 |
| Buttons | SW1 SW2 SW3 | B3F-3152 | SW410-ND | Digikey | 3 | $11.97 | 25 | $1.44 |
| 3A, 4.5 to 28V Input, EMI Friendly integrated switch synchronous step-down regulator, pulse-skipping, SOT-23-6 | U2 U3 | 296-46565-1-ND | 296-46565-1-ND | Digikey | 2 | $14.83 | 10 | $2.97 |
| IC MOTOR DRIVER PAR 16HTSSOP | U1 | DRV8833PWP | 296-29434-1-ND | Digikey | 1 | $23.57 | 10 | $2.36 |
| Button caps | #BC1 #BC2 #BC3 | Z6127-ND | Z6127-ND | Digikey | 3 | $20.23 | 25 | $2.43 |
| Hanging connectors | #J1 #J2 #J3 | H2179-ND | H2179-ND | Digikey | 3 | $2.30 | 25 | $0.28 |
| Crimps | #CR1 #CR2 #CR3 #CR4 #CR5 #CR6 | DF13-3032SCFA | DF13-3032SCFA | Digikey | 6 | $8.20 | 100 | $0.49 |
| Fan | #F1 | 381-2453-ND | 381-2453-ND | Digikey | 1 | $321.85 | 10 | $32.19 |
| Flex ribbon cable | | A9CAG-0803F-ND | A9CAG-0803F-ND | Digikey | 1 | $26.56 | 10 | $2.66 |
| Pre-crimped wire (black) | | | H4BXT-10103-B8 | Digikey | 2 | $23.64 | 40 | $1.18 |
| Pre-crimped wire (red) | | | H4BXT-10104-R8 | Digikey | 2 | $23.64 | 40 | $1.18 |
| Fabricated PCB - board only | | | | OSH Park | 1 | $117 | 6 | $19.50 |
| **Total per digital PCB** | | | | | | | | **$88.18** |

### Analog PCB - itemized

| Description | References | Value | Part Number | Vendor | Quantity Per PCB | Pack Price | Pack Size | Price per PCB |
|---|---|---|---|---|---|---|---|---|
| Unpolarized capacitor, small symbol | C12 C13 C18 C19 C20 | 10nF | 490-16599-1-ND | Digikey | 5 | $0.45 | 50 | $0.05 |
| Unpolarized capacitor, small symbol | C2 C3 C5 C6 C15 C16 | 220nF | 587-5958-1-ND | Digikey | 6 | $3.63 | 50 | $0.44 |
| Unpolarized capacitor, small symbol | C4 | 330nF | 445-11296-1-ND | Digikey | 1 | $1.90 | 10 | $0.19 |
| Unpolarized capacitor, small symbol | C1 C7 C8 C9 C10 C11 C14 C17 | 1uF | 587-6315-1-ND | Digikey | 8 | $6.18 | 100 | $0.49 |
| Generic connector, double row, 02x06, | J1 | Harting__1521012240 1000 | 1195-5658-1-ND | Digikey | 1 | $49.46 | 10 | $4.95 |
| Inductor | L1 | | 732-4872-1-ND | Digikey | 1 | $16.48 | 10 | $1.65 |
| Resistor | R5 | 180R | RG1608N-181-W-T1 | Digikey | 1 | $7.28 | 10 | $0.73 |
| Resistor | R16 R17 | 470 | MCT0603-470-CFCT-ND | Digikey | 2 | $1.34 | 10 | $0.27 |
| Resistor | R1 R18 | 1k | Y1636-1K-ND | Digikey | 2 | $128.07 | 10 | $25.61 |
| Resistor | R12 R13 R14 R15 R23 R24 R25 R26 R27 R28 | 1k | 408-2063-1-ND | Digikey | 10 | $19.11 | 100 | $1.91 |
| Resistor | R8 R9 R10 R11 | 2k | 804-1167-ND | Digikey | 4 | $403.60 | 50 | $32.29 |
| Resistor | R2 R6 R7 | 25k | PLT0603-25KACT-ND | Digikey | 3 | $42.44 | 10 | $12.73 |
| Resistor | R4 R20 R22 | 100k | A139985CT-ND | Digikey | 3 | $6.68 | 10 | $2.00 |
| Resistor | R3 R19 R21 | 2.7M | 511-1712-1-ND | Digikey | 3 | $1.36 | 10 | $0.41 |
| SiPM Sensor | SiPM1 SiPM2 | MICROFC−60035−SMT | 863-MFC60035SMTTR1 | Mouser | 2 | $657.10 | 10 | $131.42 |
| Single Ultra-Low Offset Voltage Operational Amplifier, DIP-8/SOIC-8 | U2 | ADA4522-1 | ADA4522-1ARZ-ND | Digikey | 1 | $27.25 | 10 | $2.73 |
| | U5 | ADA4522-4 | ADA4522-4ARZ-RLTR-ND | Digikey | 1 | $109.16 | 10 | $10.92 |
| IC VREF SHUNT 1V SOT23-3 | U3 U4 | ADR512 | ADR512ARTZ-REEL7CT-ND | Digikey | 2 | $52.34 | 25 | $4.19 |
| | U7 | ADS131M08IPBS | 296-ADS131M08IPBS-ND | Digikey | 1 | $95.85 | 10 | $9.59 |
| 250-mA Ultra-Low-Noise Low-IQ LDO, 3.0V, SOT-23 | U6 | LP5907MFX-3 | 296-40357-1-ND | Digikey | 1 | $5.92 | 10 | $0.59 |
| | U1 | LT3464 | LT3464ETS8#TRMPBFCT-ND | Digikey | 1 | $46.92 | 10 | $4.69 |
| Bare analog PCB, fabricated | | | | OSH Park | 1 | $117 | 6 | $19.50 |
| **Total per analog PCB** | | | | | | | | **$267.33** |
