## Supplementary material for "A handheld luminometer with sub-attomole limit of detection for distributed applications in global health": Luminometer build guide

### SiPM luminometer Build Guide

Bioengineering Platform
Chan Zuckerberg Biohub | San Francisco, CA 94103

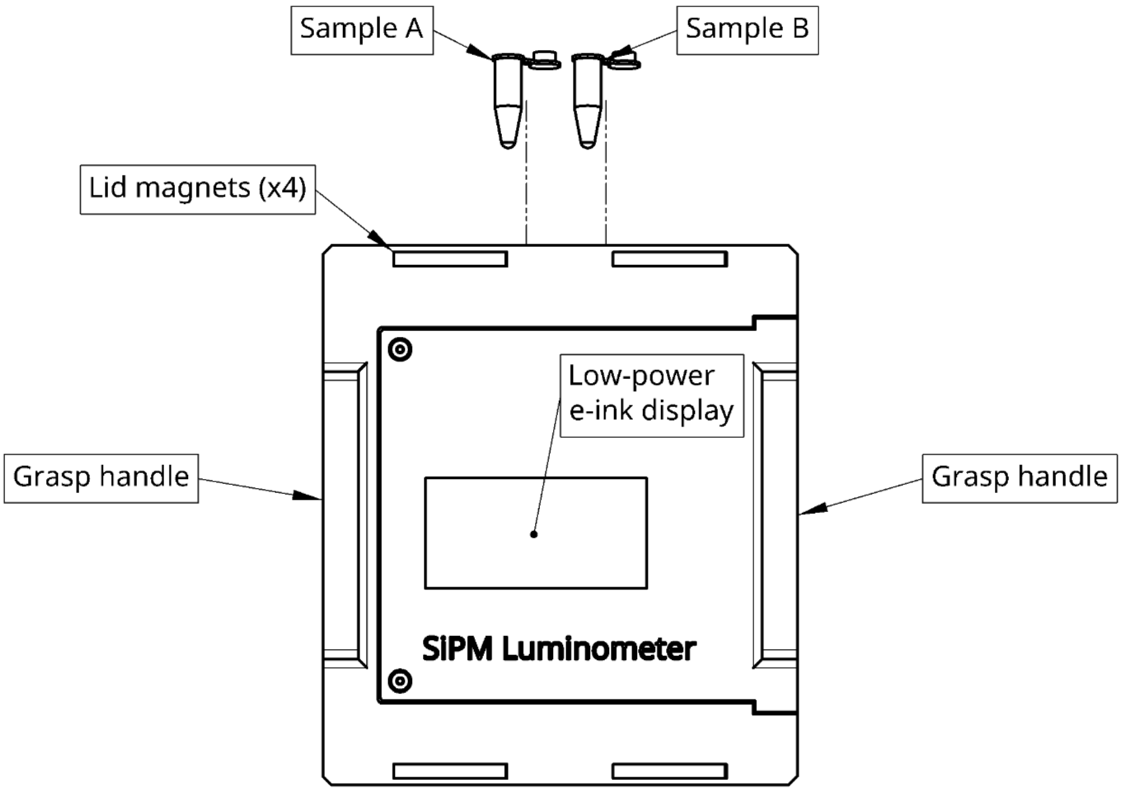


A cloud-based mechanical CAD model of the design is provided here:
https://tinyurl.com/3sh6myts

### Table of Contents


**Table of Contents** 2

**Materials and Facilities Required** 3

**Assembly instructions** 4

PCBs and Cabling 4

Digital and analog board reflow process 4

Digital PCB, post-soldering assembly steps 6

Assembly of the extra-long receptacle headers and Inky display 7

Prepare the Raspberry Pi Zero W 8

Connecting the PCBs temporarily for testing 8

No sensor test 9

Analog PCB, bottom side (low-temperature paste) 10

Powerboost 1000C cable assembly 11

Mechanical assembly 11

Powerboost cover panel fabrication 11

Preparation of tube holders for painting 11

Preparation of the main enclosure parts 12

Total ground painting 12

Black 3.0 painting 14

Post-paint assembly steps 16

Fan installation 18

Shutters 19

Shutter drivetrain 19

Pre-assembly with PCBs 21

Shutter wiring 26

Final Assembly 27

Mounting the electronics 27

Battery installation 27

Install the tube holder assembly 28

Installation of cover panels 29

Supplementary Photos 30

### Materials and Facilities Required

In addition to all the items listed in the Bill of Materials (BOM), building the devices requires the following equipment and facilities:

- Top and Bottom side laser-cut solder stencils for each PCB (Vendor used: OSH Stencil)
    - Note: .gbr and .pdf files for solder paste stencils were exported from Kicad were uploaded to the vendor's website and ordered)
- Reflow oven (Model used: LPKF Protoflow S)
- Solder paste stencil printer (Model used: Neoden FP2636)
    - Note: the stencil printer is optional and can be replaced with careful hand-alignment of the stencil to the PCB. It is common to hand-align stencils and secure them with tape prior to application of solder paste.
- Solder paste:
    - Chipquik SMDLTLFP500T3C (low-temperature)
    - Chipquik SMD291SNL(high-temperature)
- Fine-tipped tweezers
- Conductive, grounded work surface
- Stereoscopic microscope (Not essential but highly recommended)
- Spool of lead-free solder for hand-soldering
- Soldering iron station
- Solder smoke exhaust fan (recommended)
- Hand tools:
    - Set of metric allen keys
    - Set of small Torx wrenches (T1-T8)
    - Ball peen hammer
    - Wire strippers, flush cutters, pliers, tweezers
    - Wire crimping tool
    - Set of miniature flat wrenches (Moody #58-0161)
    - Modified 4mm miniature flat wrench (thinned down on a stone grinder)
- Loctite 420 (cyanoacrylate glue)
- Total ground conductive spray paint
- Black 3.0 acrylic paint
- Fine-tipped paint brushes
- Isopropanol
- Lint-free absorbent wipes

### Assembly instructions

#### PCBs and Cabling

##### Digital and analog board reflow process

**Important notes:**

- Always work on an ESD-safe work bench when manipulating electronics components.
- First, work with high-temperature solder paste to assemble and reflow the components on the top side of the board.
- Subsequently, use the low-temperature solder paste to reflow components on the bottom side without affecting the top side.
- Only surface-mount components get reflowed. All through-hole components should be hand-soldered after the reflow process has been completed on both sides.
- Before reflowing the bottom side of the analog board (which includes the two SiPM sensors), we **highly** recommend completing the remaining assembly steps in the electronics section and performing the “No sensor test” in order to validate the functionality of the rest of the system prior to installing the sensors.

**Step-by-step reflow instructions for each PCB:**

1. Ensure the correct reflow profiles are set up on the oven, for each type of solder paste.
2. Visually inspect the fabricated boards for manufacturing defects.
3. Remove the solder paste from 4C storage and allow it to warm to room temperature.
4. Follow instructions for the stencil printer to install and constrain the bare PCBs (doing one at a time) with the top side facing up.
5. Align the laser-cut stencil to the PCB, following the stencil printer’s instructions.
    a. Note: This can also be done by hand on a clean benchtop, using tape to secure the PCB and stencil in place.
6. Once aligned, apply solder paste to the top of the stencil and squeegee the paste across the stencil using firm pressure.
    a. Note: solder printer came with a mock plastic credit card to be used as a solder squeegee.
7. Carefully lift the stencil vertically from the PCB surface such that the solder paste pattern is not disrupted.
8. Inspect the bare PCB to confirm the placement of the solder paste on top of the pads.
9. Carefully place the PCB under the stereoscope for assembly of the components.
    a. Tip: a stereo microscope on an articulating arm was used in this case, allowing the PCB to sit on an ESD-safe work surface.
10. Using fine tweezers and the PCB layout as a reference, place each surface mount component on the board in its designed location and orientation, using extra care not to smudge the patterned solder paste, and without disrupting placed components.

11. Carefully place the PCB in the reflow oven and begin the profile.
12. Allow the PCB to fully cool before removing.
13. Inspect all components and their solder joints under a stereomicroscope, checking for poor connections or shorts (bridges of solder between pads can occasionally occur.
14. Repeat the process for the back side of the PCB, using low-temperature paste.
15. After both sides of the board have been reflowed and inspected, proceed with hand soldering all the through-hole components.
16. The analog PCB should appear as follows:
    a. **Note**: do not reflow the bottom side of the analog PCB until the ‘No sensor test’ has been completed.

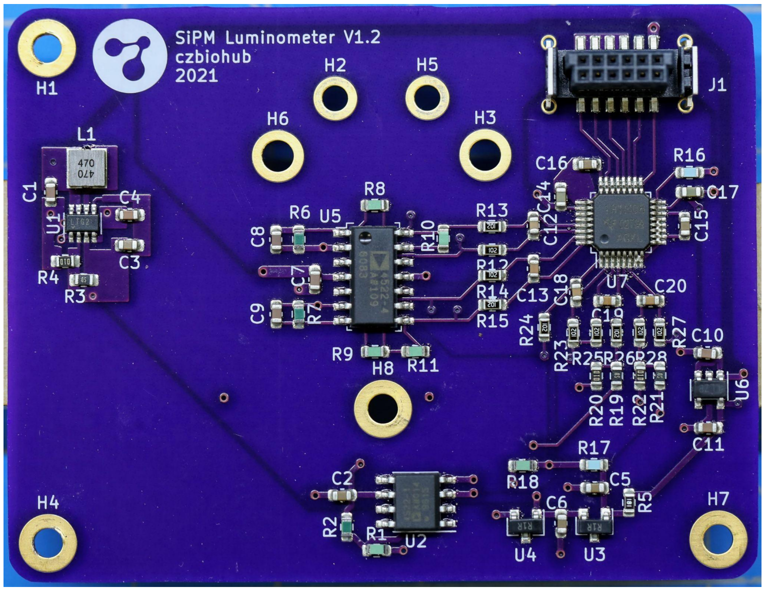


**Digital board, component-specific tips:**

1. For the RPi 40-pin male header (J4):
    a. Ensure male pin header is firmly seated and properly normal to the board.
    b. First solder two pins on opposite sides, checking for proper seating of the pins.
    c. Then proceed with soldering the remainder.
2. Fan header (J5):
    a. Follow orientation shown in the OnShape model
3. Shutter headers (J2,3):
    a. Follow orientation shown in OnShape model
4. UI Button switches (SW1,2,3):
    a. Ensure they are firmly seated against the board before soldering
5. Snap on button caps only after switches are soldered (#BC1,2,3)
6. Power switch (SW4):
    a. There is one extra hole in the switch footprint - ignore it.
7. Headers (J6,8) are unused in this design iteration.

#### Digital PCB, post-soldering assembly steps

8. Connect an M2.5 x 8 mm (FF) standoff on the bottom side to an M2.5 x 16 mm (MF) standoff on the top side, using a single washer between the board and the 8 mm FF standoff.

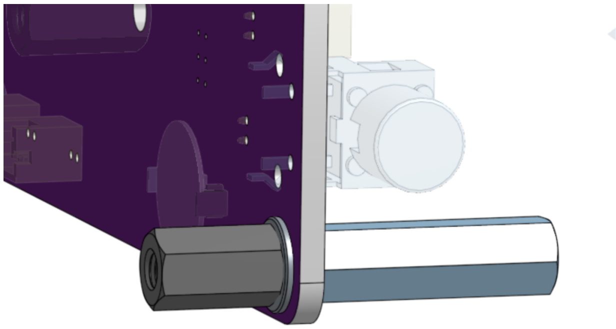

9. Install the remaining standoffs (two locations), using an M2.5 socket cap head screw (L=4mm) from the other side. Tighten firmly as we later connect to the other side of the standoff and it should not loosen. The PCB should appear as follows:

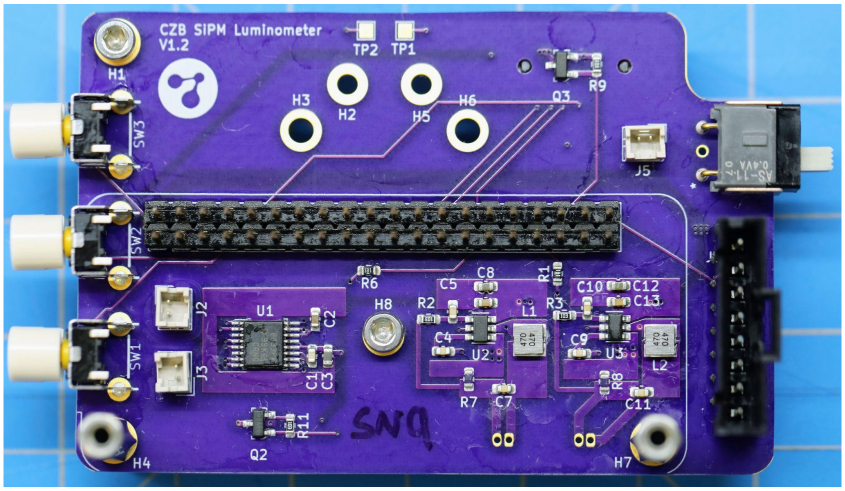


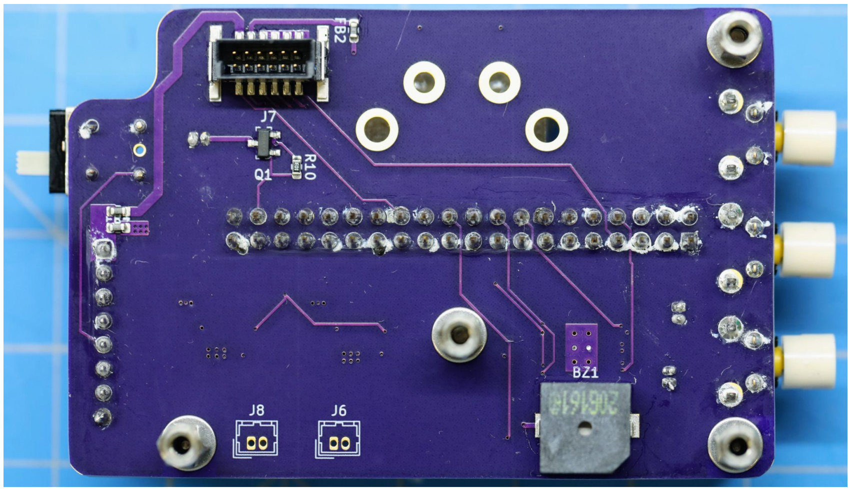

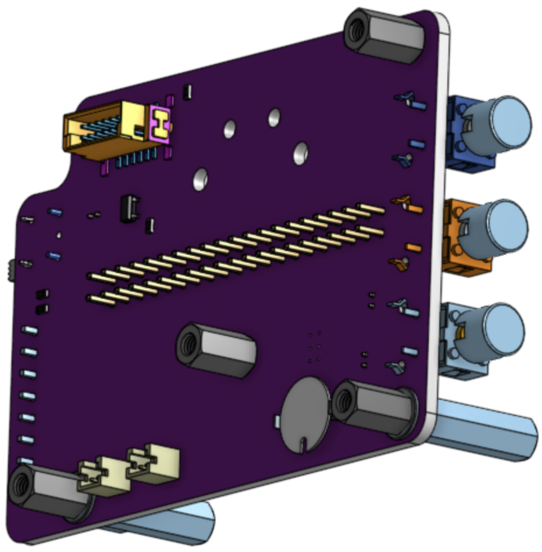

#### Assembly of the extra-long receptacle headers and Inky display

- Install laser-cut header spacer (3-0811) onto the long pins of SAM12900-ND prior to soldering.
- Solder the extra long 40-pin receptacle (SAM12900-ND) to RPi, ensuring it is totally flush and fully-seated against the PCB.
    - Be very careful: solder pin base only. Do not damage RPi components with iron.
- See OnShape for orientation: the pins should protrude out the top of the board

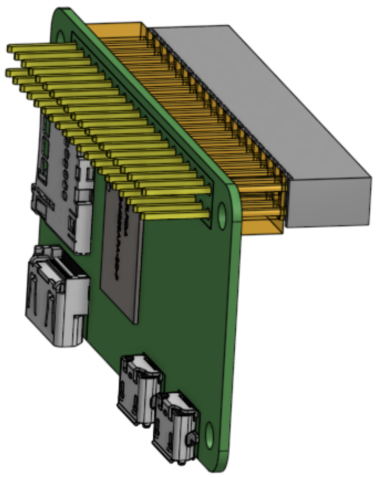

- Install Inky pHat onto the extended GPIO pins
- Pi + Inky sub-assembly:

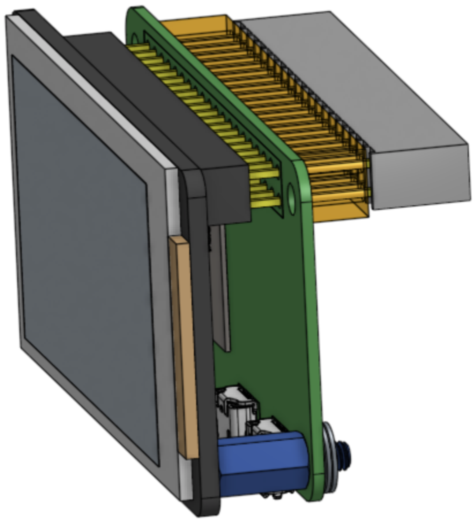

- Install the M2.5 x 8mm M/F standoffs into the lower holes of the RPi. These will later be tightened into the M2.5 x 16 mm standoffs that extend off the digital board.

#### Prepare the Raspberry Pi Zero W

1. Flash a 16GB microSD card with Raspbian image.
2. Follow online instructions for adding local Wifi information to the SD card (local network credentials required).
3. Install the SD card into the Raspberry Pi.
4. Power on via USB cable
5. Verify that you can ssh into device:
    a. *ssh pi@freshlumi1.local*
    b. pw: *fresh1*
6. Power down the Raspberry Pi:
    *a. sudo poweroff*

#### Connecting the PCBs temporarily for testing

The two PCBs can be connected via the Harting board-to-board connectors shown in orange. It is recommended that screws are added to the standoffs for support, to avoid strain on the connectors. They are also useful as they protect the sensors from contacting any surfaces:

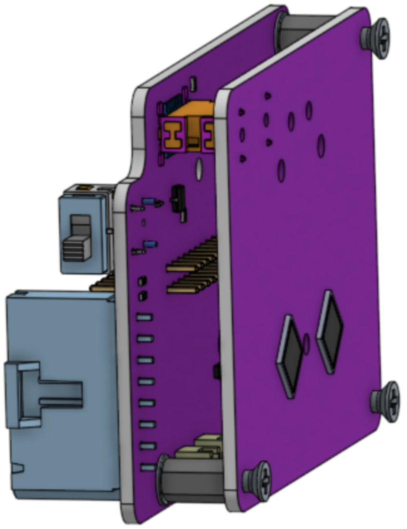

Now, connect the Pi + Inky sub-assembly to the PCBs. The 40-pin receptacle takes some gentle but firm pressure to seat and properly connect:

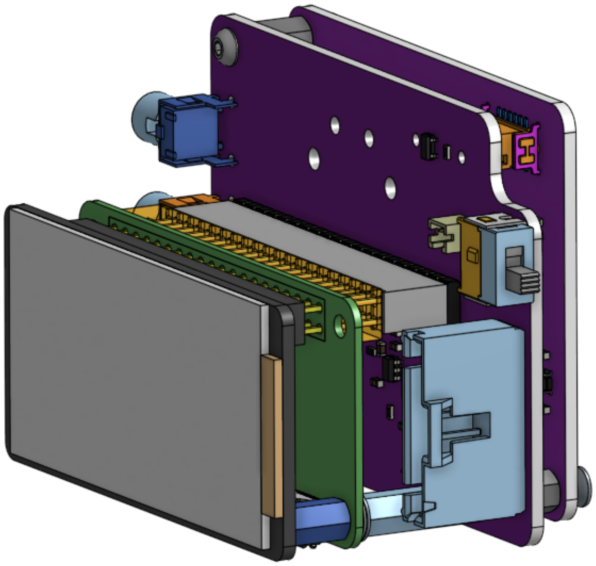

Note: The Raspberry Pi + Inky display + SAM12900-ND sub-assembly will be removed again in later steps in order to make connections to the digital PCB.

#### No sensor test

This can be performed now that the raspberry pi is set up, and by connecting it to the PCBs. The no sensor test verifies the parameters of the amplifier in the absence of any dark current. This measurement is otherwise impossible to perform without introducing an open circuit between the sensors and the amplifier.

- Power on the device by connecting it to a micro-USB power supply
- Navigate to the diagnostic menu using the buttons
- Record all values from the diagnostic menu
- CHA and CHB raw values should both measure between -0.433 and -0.429 V. These are the baseline values of the amplifier in the absence of any sensor current.

After completion of this test, the sensors can now be reflowed onto the bottom side of the analog board and the electronics re-assembled.

#### Analog PCB, bottom side (low-temperature paste)

Follow the same procedures as soldering the other PCB sides.
→ The sensors are fragile before being installed into the PCB, so handle them with care.
→ Ensure the sensors are well-centered prior to reflow.

After the sensors have been reflowed, the PCB should appear as follows.

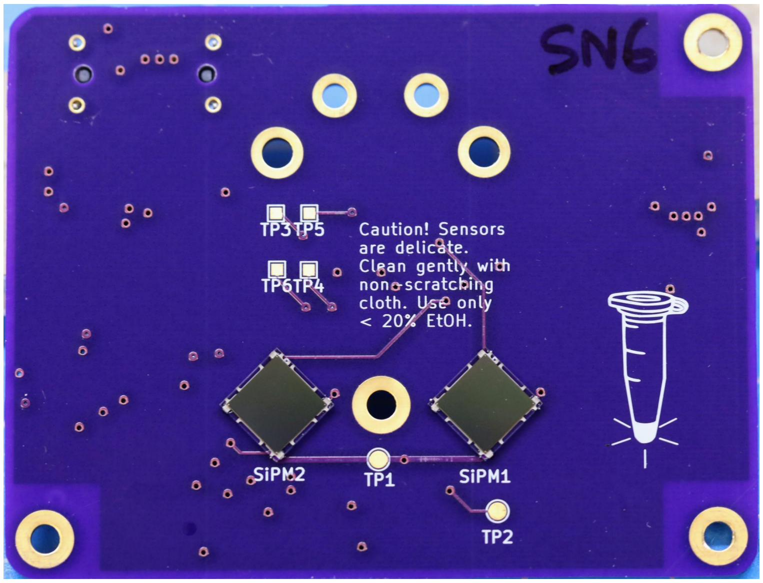


Note the orientation of the sensors is indicated by subtle markings on the perimeter.

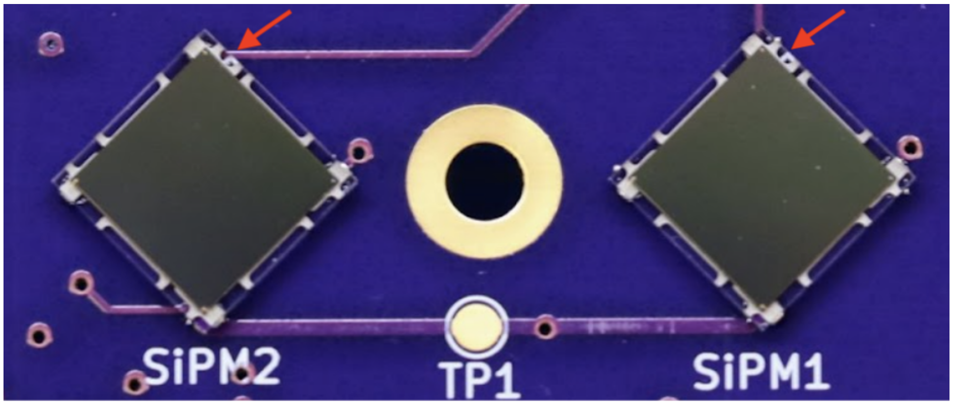

After the sensors have been reflowed, the PCBs should be re-assembled with standoffs in the same configuration as the 'No sensor test'.

##### Powerboost 1000C cable assembly

Make the cable to connect the Powerboost 1000C to the Digital PCB:

1. Cut four 26 AWG insulated wires to 20 cm length: red, blue, black, green
2. Strip 1.5 mm segments from the ends of one side, and a 4mm segment from the other end of each wire.
3. Solder the wires into the following sockets of the Powerboost 1000C chip:
    a. 5V output (**Red**)
    b. LBO (**Blue**, Low battery out)
    c. GND (**Black**)
    d. EN (**Green**, Enable)
4. Install wire crimps (DK A105462CT-ND) on the other side using a crimp tool.
5. Insert into connector (DK 487526-7-ND) into the following positions:
6. **Red** = 1, **Blue** = 3, **Black** = 4, **Green** = 5
7. Solder a 10k resistor inline with the blue (Low battery - LBO) wire. Heat shrink.
8. Twist the wires together in a bundle.
9. Powerboost 1000C screws installed
10. Slip incoming wires through the slot in the main enclosure.
11. Fasten thread-forming screws to secure the powerboost 1000C to the enclosure.

#### Mechanical assembly

##### Powerboost cover panel fabrication

The cover panel (3-0984) is a 1 mm thick, laser-cut polycarbonate panel (recommended material due to its increased strength over acrylic).

1. Spray paint one single side a piece of 1mm thick stock polycarbonate sheet using total ground paint. The painted area should be larger than the ~86 x 44 mm outline of the part.
2. Laser cut the part, as obtained by exporting the back side of the part (3-0984) from OnShape as .dxf.
3. Optional: Indicator text and transparent boxes can also be etched into the panel using a raster scan on the laser cutter:

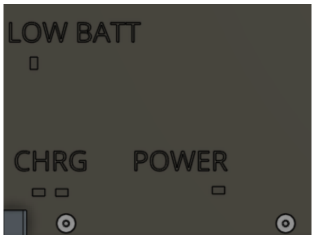

4. Countersink the holes to a 2.9 mm diameter using a standard countersink bit.

#### Preparation of tube holders for painting

1. Clean the tube holder lid (3-0678) and base (3-0677) with compressed air to remove any dust from the fabrication process.
2. Glue all magnets (K&J DH11, qty. 14) into the base (3-0677) and lid (3-0678) using Loctite 422.

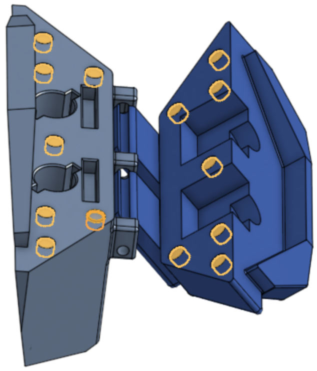

3. Drill out PCR tube holes with 0.25" drill bit.
4. Clean all dust/debris with compressed air.

#### Preparation of the main enclosure parts

- Press-fit and glue the magnets (K&J BX021, qty. 8) into the main enclosure and rear access lid using Loctite 422. The fit of the magnets may be tight - be careful not to crack the enclosure parts when applying pressure. If necessary, sand the faces of the magnets until they fit snugly but without needing excessive pressure to press in. Remove all dust from the magnets after sanding. Sanding also increases the strength of the bond when gluing.
- Note: the orientation of the magnets must be such that matching pairs attract each other.

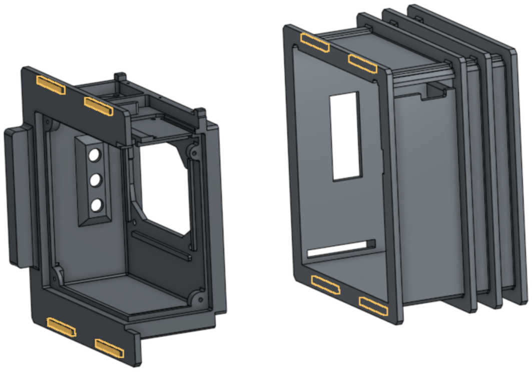

#### Total Ground painting

Two coats may be required to obtain complete coverage:

- Enclosure - front/side panel (3-0861, all surfaces)

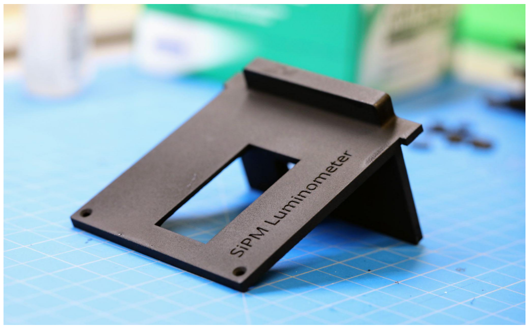


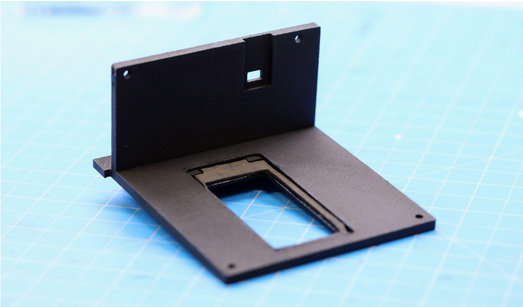

- Enclosure - main (3-0682, all surface except masked region shown below)
    - Before painting, mask Powerboost mounting surface with tape

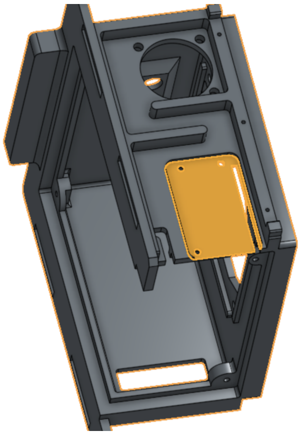

- Enclosure - rear access lid (3-0684, all surfaces)

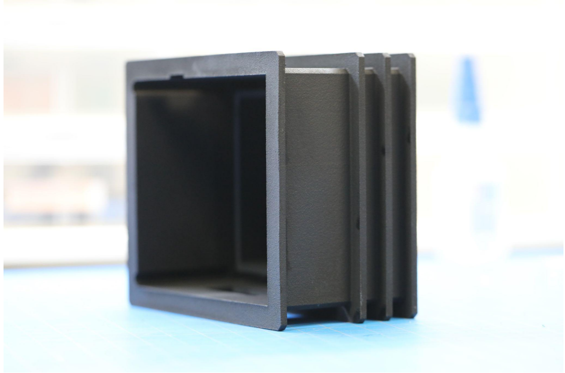

- Tube holder lid (3-0678, all surfaces) and base (3-0677, all surfaces)

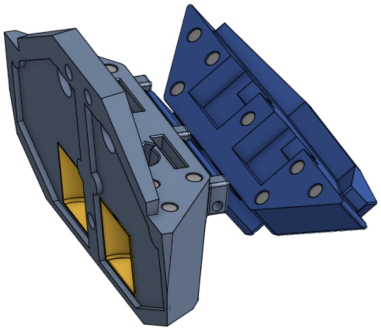

- Shutter flag (3-0717):
    - Gently sand the stainless steel surfaces with 300 grit sandpaper
    - Clean all dust and debris and allow to fully dry prior to painting

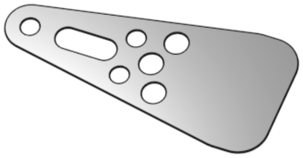

#### Black 3.0 painting

Note:

- Ensure the Total Ground paint has fully dried for > 24 hrs before applying Black 3.0.
- Standard craft supplies (artistic paint brushes) can be used for applying the paint

- Enclosure - rear access lid (3-0684), only the interior surfaces:

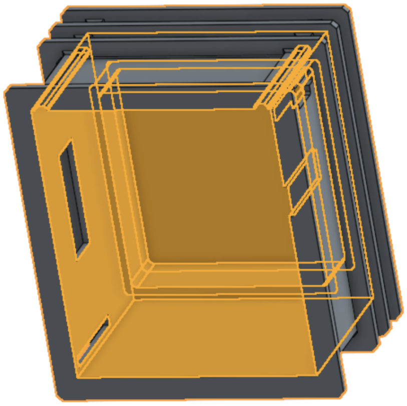

- Enclosure - main (3-0682), only selected surfaces (3):

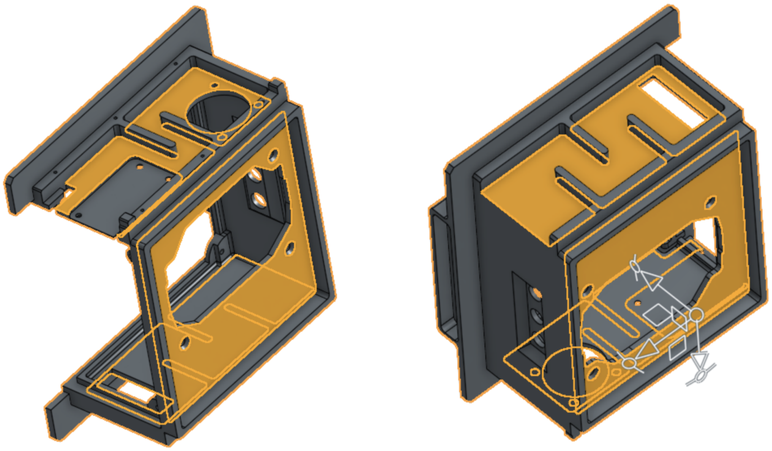

- Tube holder base (3-0677), sample cavity and shutter face:

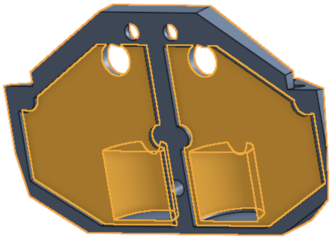

  - Tube holder after painting with total ground and Black 3.0:

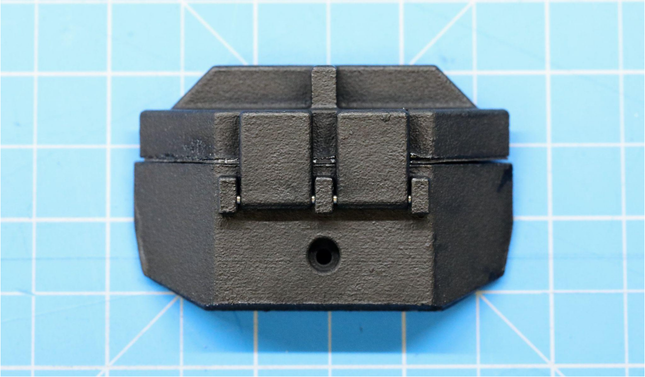

- Shutter flag (3-0171):
  - Paint only the bottom half with Black 3.0, as the top hole is used for mounting and Black 3.0 paint is not durable. The below photo was captured using very bright illumination, showing how much darker black 3.0 is than total ground.

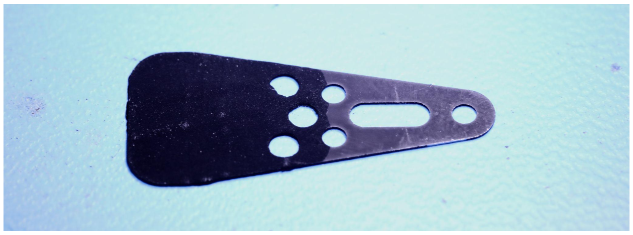

#### Post-paint assembly steps

- Glue the button gaskets (3-0825 shown in orange) onto the inner surface of the main enclosure (3-0682) using Loctite 422. Ensure it sits flush on the face:

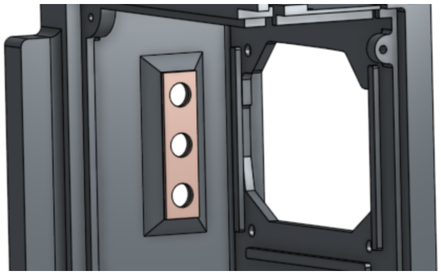

- Glue the screen gasket (3-0883 shown in blue) onto the enclosure front/side panel (3-0861) using Loctite 422.

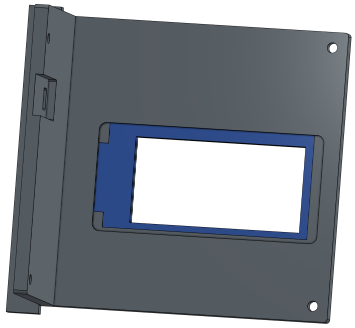

- Glue the tube holder lid gasket (3-0680 shown in silver) to the tube holder lid (3-0678) using Loctite 422, ensuring it is completely flush.

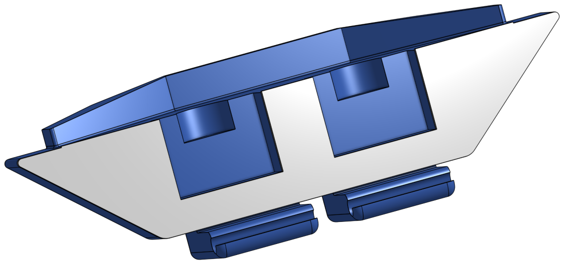

- Carefully drill open the shutter clearance holes in the tube holder base (3-0679) with a #1 drill to 5.8 mm.

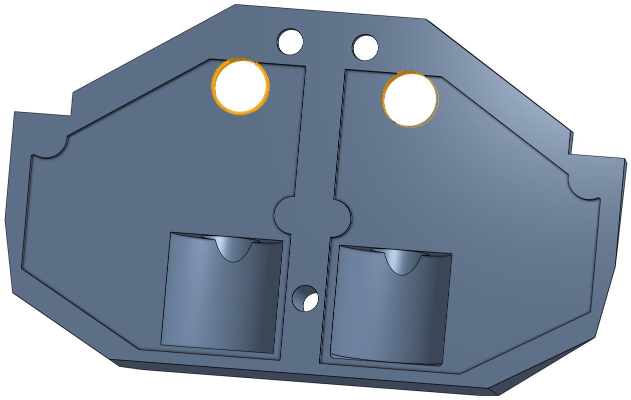

- Glue the tube holder base PCB gasket (3-0679 shown in grey) to the tube holder base (3-0677) using Loctite 422.

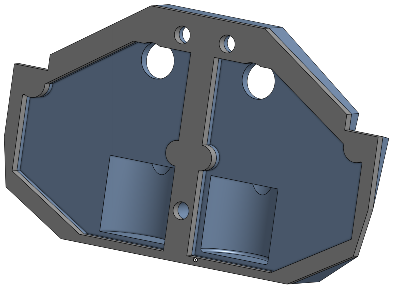

- Close the tube holder lid (3-0678) onto the tube holder base (3-0677). Slide the pin (McMaster-Carr 90145A425) through the hole through the clamps on the lid, resting in the slot. Cover the hole with silicone sealant (Dowsil 700).

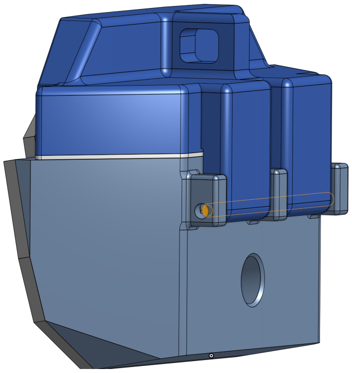

- The tube holder lid should now pivot about the installed pin

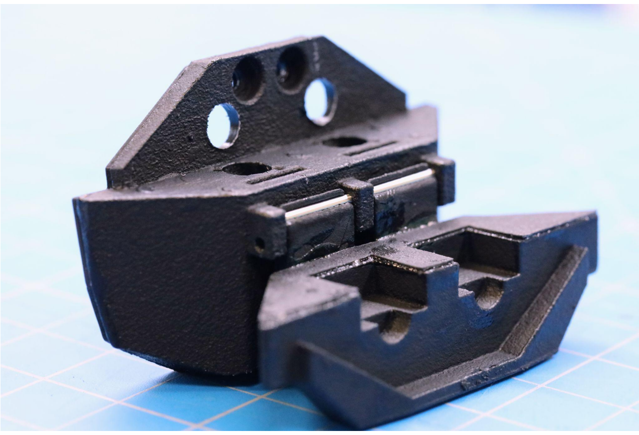

- The reflective mylar strip (3-0710) is laser cut into two pieces, a rectangle and a semicircle. Glue the mylar into the tube holder base (3-0677) cavities using Loctite 422. Press with the mylar glue guide (3-0999) to keep in place.

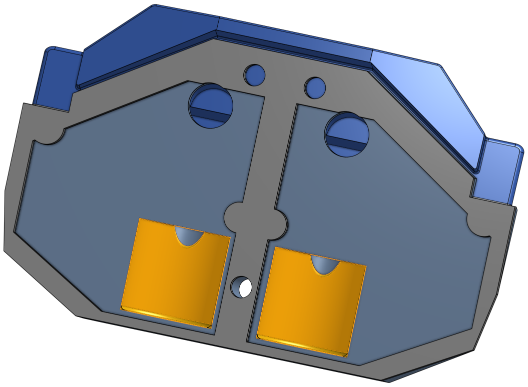

#### Fan installation

1. Insert pre-crimped wires (H4BXT-10103-B8 (black) and H4BXT-10104-R8 (red)) into the connector (H2179-ND), respecting the polarity of the connector as specified in the PCB schematic.
2. Extend the length of the cable to 12 cm by soldering additional lengths of hook up wire. Ensure the joints are strong and well-covered with heat shrink tubing.

3. Solder the wires to the fan connectors. Only 2 out of three terminals are used. See image for reference.
4. Mount the fan onto the main enclosure, 3-0682 using #4 thread-forming screws, qty 3 (McMaster-Carr 97349A100). Note that the enclosure holes are used as clear holes, and the screws form threads into the fan's plastic holes.

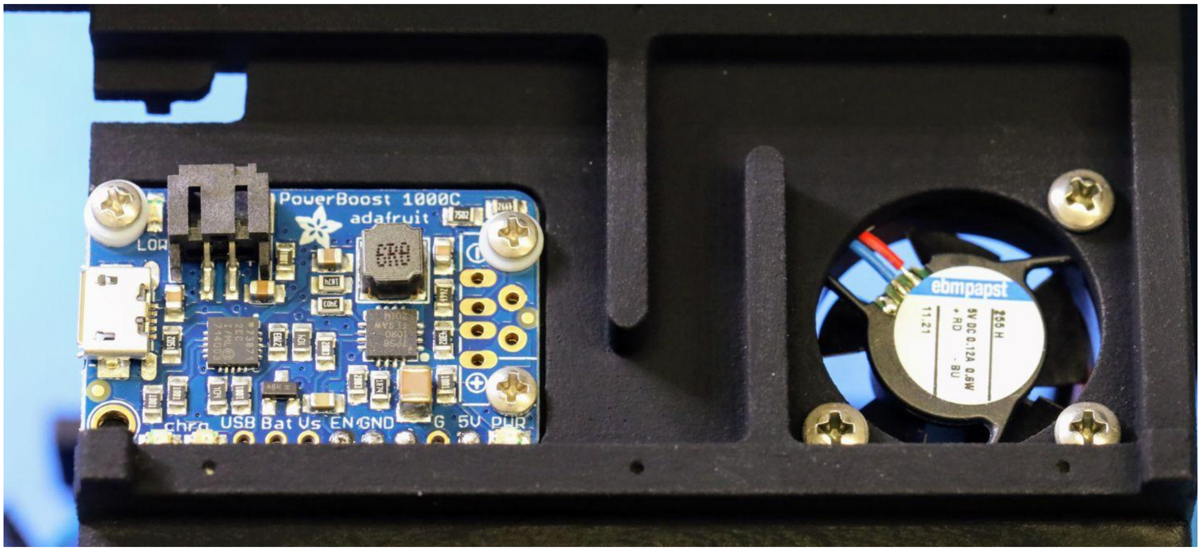

Main enclosure with Powerboost 1000C and fan installed.

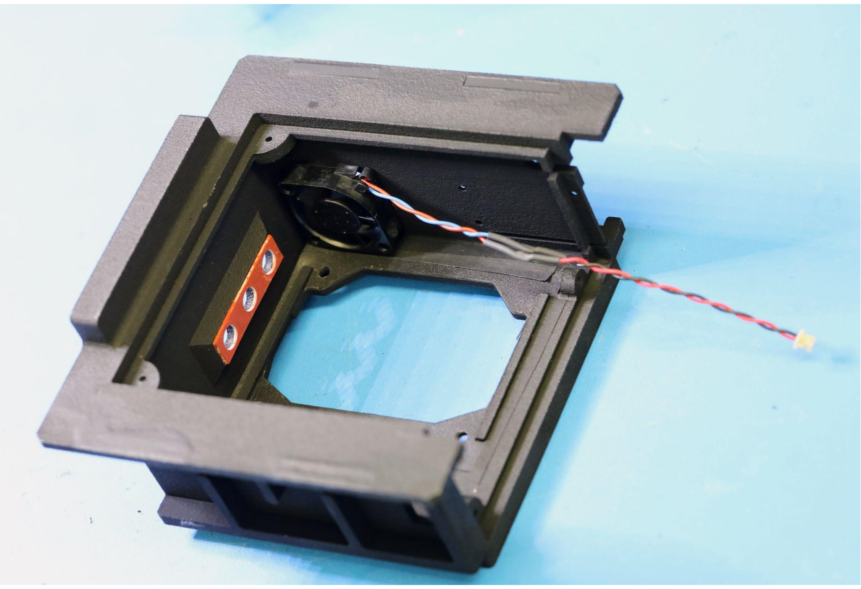
Main enclosure with button gasket and fan installed

#### Shutters

The shutters are the most difficult and intricate portion of the instrument, and require careful assembly.

#### Shutter drivetrain

1. Press-fit the bearings (McMaster-Carr 7804K119, qty 4) into the shaft spacer (3-0716). Note that the motor mount spacer may need gentle reaming in order to permit entry of the bearing. If the spacer pocket is machined too large for a tight fit:
    a. The bearing may be inserted and aligned against the back wall. While maintaining alignment pressure, *gently* tap the thin portion of the wall inwards with a ball peen hammer until a very small deflection is achieved and the bearing is held in place firmly in the pocket.

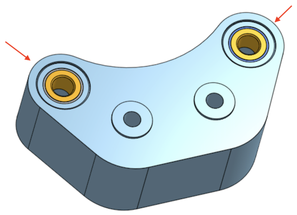

2. Mount the motors (Amazon B07ST8FML4, qty. 2) into the motor mount (3-1003, qty. 1) using M2.5 x 10 socket cap hex screws. **Only tighten the screws until the motors are snug: do not crush the motors:**

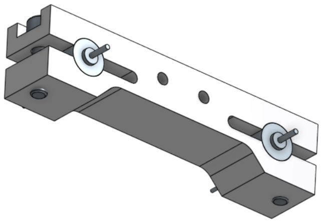

3. Drill out the central bore of the drive gears (3-0890, qty. 2) to 0.8 mm using a miniature 0.8mm bit.

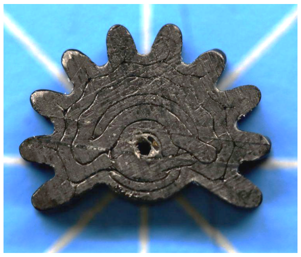

4. Clean the motor drive shafts *sparingly* with isopropyl alcohol to prepare the surface for gluing. Use a lightly-soaked tissue to perform the cleaning: **do not allow alcohol to wick inside the motor**.
5. Press-fit the mounted motor shafts onto the drive gears .
    a. We recommend that this is done with the gear sitting flush against a flat table, and downward force applied to the motor mount assembly from above, pushing on the body of the motor. **Application of force to the motor mount can result in dislocation of the motor from its case.**

b. 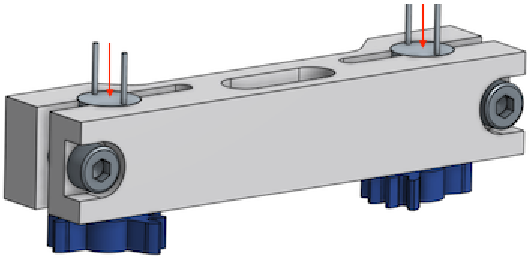

c. 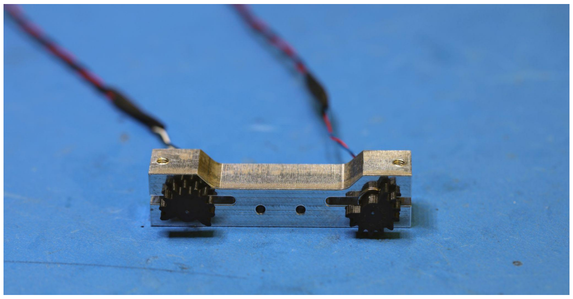

d. 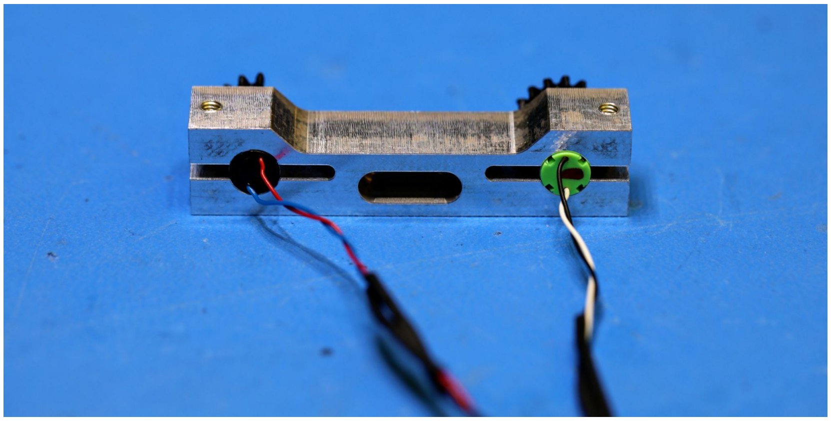

6. Using a 2 uL pipette, carefully apply 0.3 uL of Loctite 420 to the shaft/gear interface that is farthest from the motor. The glue will wick into the press-fit shaft surface. **If too much glue is applied, it could wick into the motor, rendering it unusable.**
7. Use M2 x 18mm socket cap hex screws to self-tap into the follower gears (3-0889, qty. 2). Apply ~1 uL of Loctite 420 to the interface formed by the head of the screw with the gear.

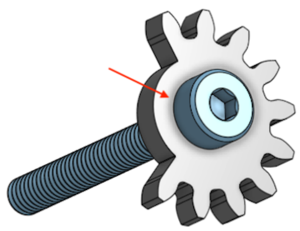

#### Pre-assembly with PCBs

1. Mount the shaft spacer (3-0716) *temporarily* to the digital PCB using 2x short M2 screws.

2. Attach the analog board (with its M2.5 x 8mm hex standoffs, qty. 3) to the digital board via their interconnects. This connection is not load-bearing, so additionally secure the boards from behind *temporarily* with 3x M2.5 screws, fastened from the back side of the analog board.
    a. The shaft spacer is now clamped between the two boards via pressure from the temporary screws.

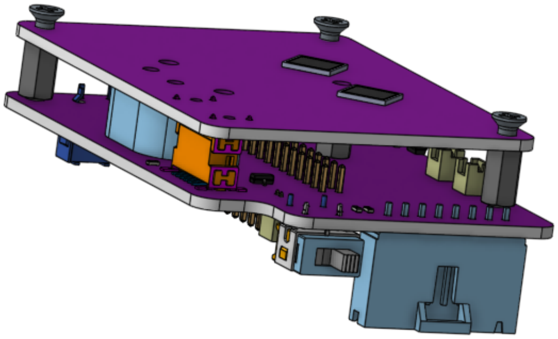

3. Next, remove temporary screws from the shaft spacer on the digital side so that it is held only by clamping pressure between the PCBs.
4. Insert follower gears and shafts through assembly so they thread through the shaft spacer. Include 2x M2 washers per shaft, between the follower gear and the PCB:

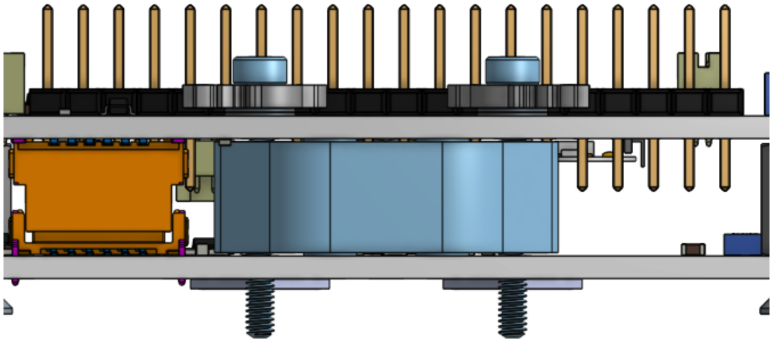

5. Secure the motor mount, including 3-0892 motor mount spacer, and using M2 screws of length 16 mm, qty. 2:
    a. Position the assemblies as shown in the photo, to avoid parts from falling due to gravity.
    b. Rotate the follower gears to the angles shown in the photo, such that they will mesh correctly with the drive gears. Notably, the toothless gaps need to be positioned such that they will not interfere with the motor mount spacer (3-0892) once it pivots into place.
    c. Pivot the motor mount assembly downwards into place so that:
        i. The drive gears mesh with the follower gears
        ii. The follower gears to not interfere with the motor mount spacer
        iii. The 16 mm long M2 screws enter the PCB clear holes and thread into the tapped holes in the shaft spacer (3-0716).

d. 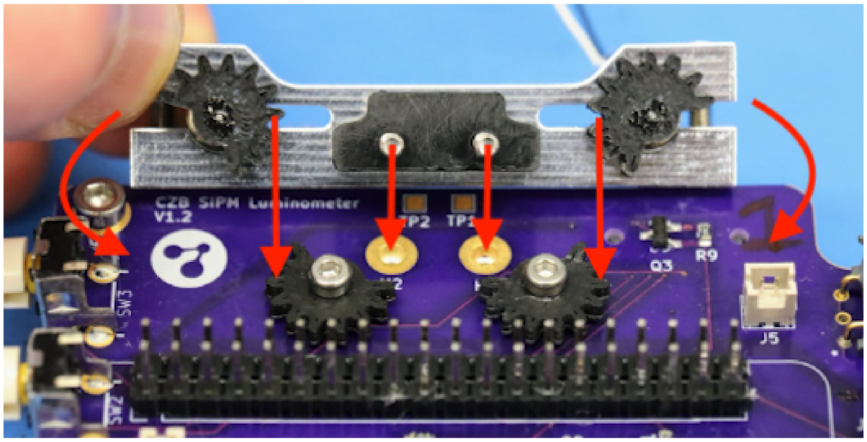


e. 

f. 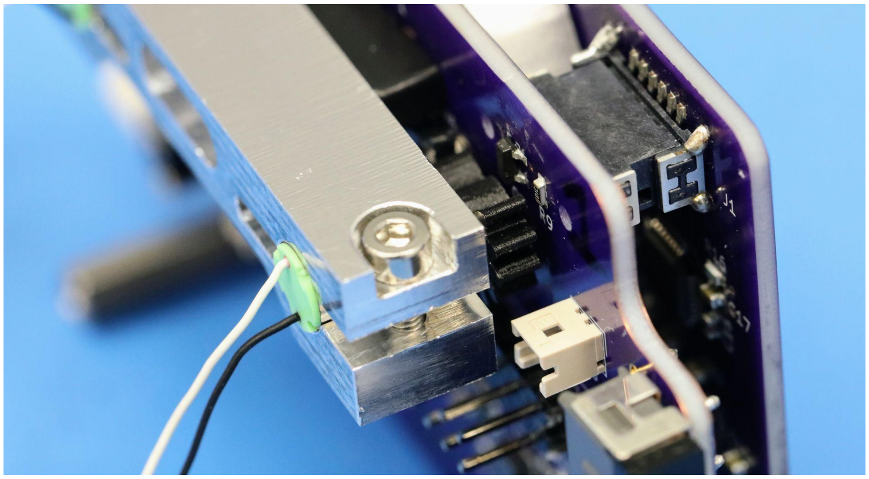

g. Side view of mounted motors

6. Tighten the screws slowly, rotating gears into position such that they mesh.
7. Ensure both gear systems are able to rotate freely without friction within their designed range of motion. Misalignment of the shaft spacer can cause the system to bind up.
8. Attach one M2 washer and one M2 thin brass hex nut to the protruding shutter screw in order to secure the assembly against falling apart.
    a. Adjust the brass nuts until they are adjacent to the PCB but not exerting pressure.

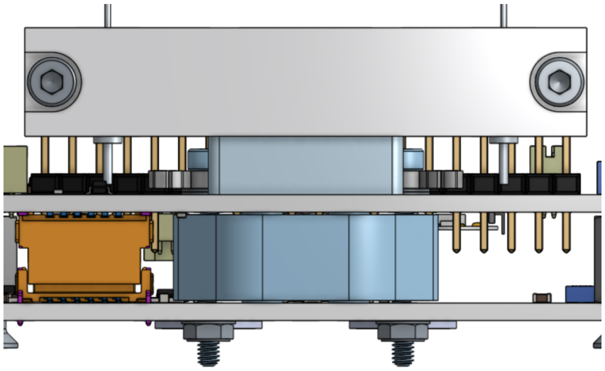

b.

9. Install the shutter flags onto the 18 mm length M2 screws.

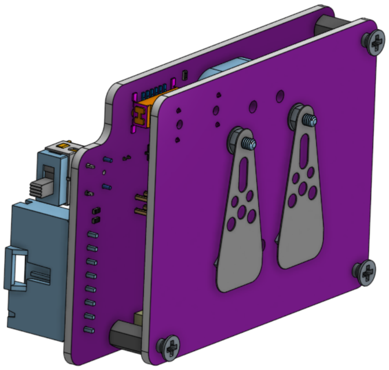

10. Compress the shutter flag between the first brass nut and a second brass nut
    a. Use the laser-cut shutter alignment tool (3-1015) to lock the shutter angle in place for tightening of the M2 lock nuts.

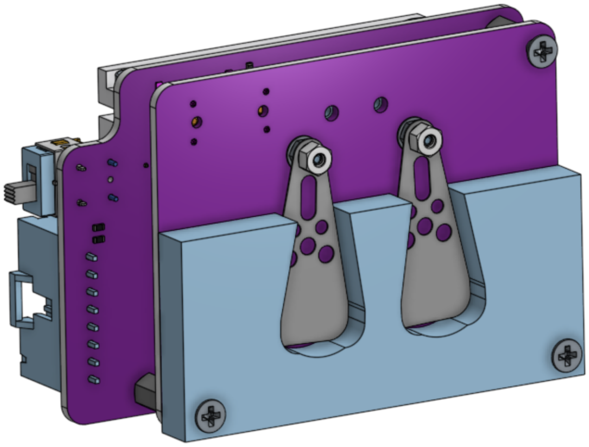

11. **Caution:** Tightening the lock nuts is tricky because you must use two miniature 4 mm wrenches, one on each nut (one regular wrench, and one modified) to lock the shutter flags in place without applying torque to the plastic gears on the other side of the assembly. This must also be done while keeping the gears in the correct position, as shown below. Locking the shutter flags in place sets the angle of the flag with respect to

the drive system, and must allow for the correct range of motion. It can help to wrap an elastic band around the follower gears to bias the position of the gears against the stop, in the shutter closed position.

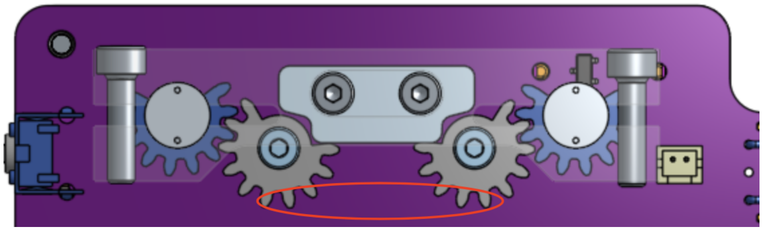

12. Tighten the nuts to compress the shutter flag while it is in the closed position. Take care not to scratch the sensors.
13. Ensure that the flag is compressed between the nuts, and such that they do not exert pressure on the PCB and can rotate freely.
14. Repeat for the other shutter.
15. Remove the shutter flag alignment tool.
16. After removing the alignment tool, check that the angular range of motion allows full blocking/unblocking of the sensor.
    a. Closed: 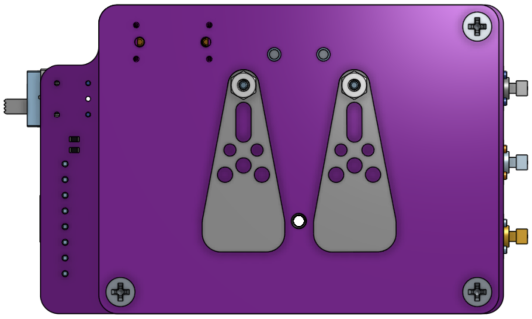
    b. Open: 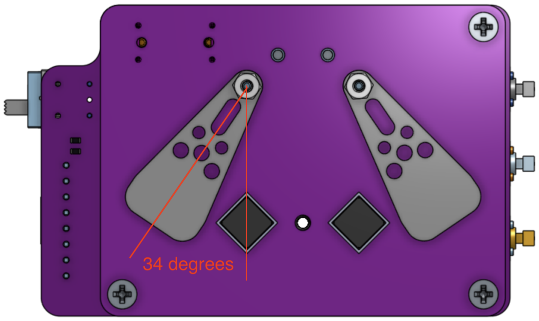

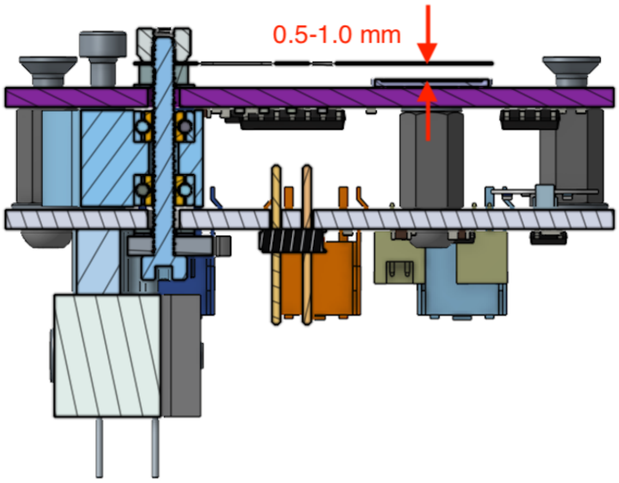


Note: there should be 0.5-1.0 mm of clearance between the shutter flag and the sensor. If the shim steel flags are bent or damaged, they may collide with the tube holder or the sensor. Before proceeding, verify that the spacing is between 0.5 and 1.0 mm. It is possible to bend the shutter flags *very* gently to correct **small** deflections. Otherwise, it is recommended to start with freshly-cut, unwarped parts.

#### Shutter wiring

1. Insert the pre-crimped wires (H4BXT-10103-B8 and -R8: **black** and **red**, respectively) into the two connectors (H2179-ND), minding the polarity. Use **red** wire for both ‘1’ outputs, and **black** wire for both ‘2’ outputs:

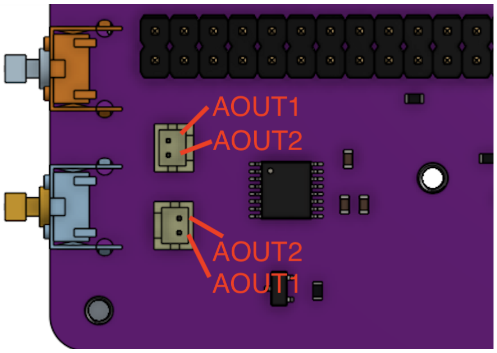


2. Complete the wiring from the hanging connectors to the shutter motors by soldering a junction, joining the shutter motor wires to the free ends of the connector wires. The motors have either **red** and **blue**, or **black** and white outputs. Connect **red** to **red**, and **blue** to **black**, or if they are **black** and white, connect white to **red**, and **black** to **black**.

### Final Assembly

#### Mounting the electronics

1. Insert the electronics assembly into main enclosure, gently sliding the buttons through button gasket on enclosure, and fasten on the rear side with the three M2.5 countersunk screws:

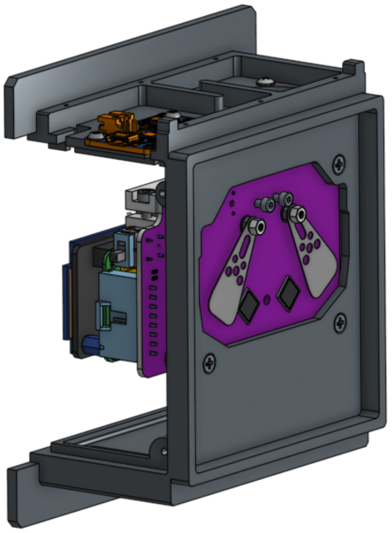

2. Fasten the PCB assembly to the enclosure via 3x M2.5 countersunk screws from the rear side.
3. Plug the fan connector into J5 on the digital PCB.
4. Plug the Powerboost 1000C cable into J1 of the digital PCB.
5. At this point, the RPi + Inky display sub-assembly can be re-installed onto the 40-pin GPIO header on the digital PCB.

#### Battery installation

6. Insert the battery (Adafruit #354) below PCB assembly:
    a. The battery is a tight fit and can be wiggled in with a small amount of force. Do not apply excessive force.
    b. The battery retention clip can be inserted by flexing the teeth inwards and sliding them past the internal retention rail inside the main enclosure. The clip itself is a flexural element and can be deflected during installation. The battery should be held in place securely via tension from the clip.

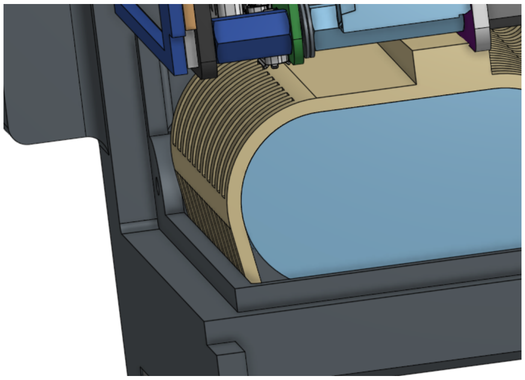

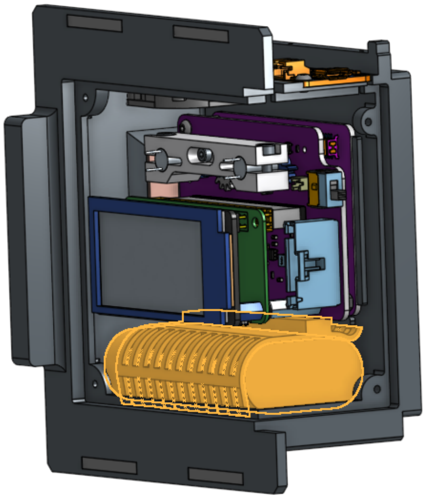

7. Ensure the main power switch (SW4) is turned off (down position).
8. Connect battery cable to the JST connector on the Powerboost board.

#### Install the tube holder assembly

The tube holder assembly can be installed after the shutters are completely assembled. Use qty. 2 M2 x 6 mm length screws for the upper mounting holes, and qty. 1 M2.5 x 14 mm length screw for the lower mounting hole. Before fully tightening, ensure that the shutter motion is not inhibited.

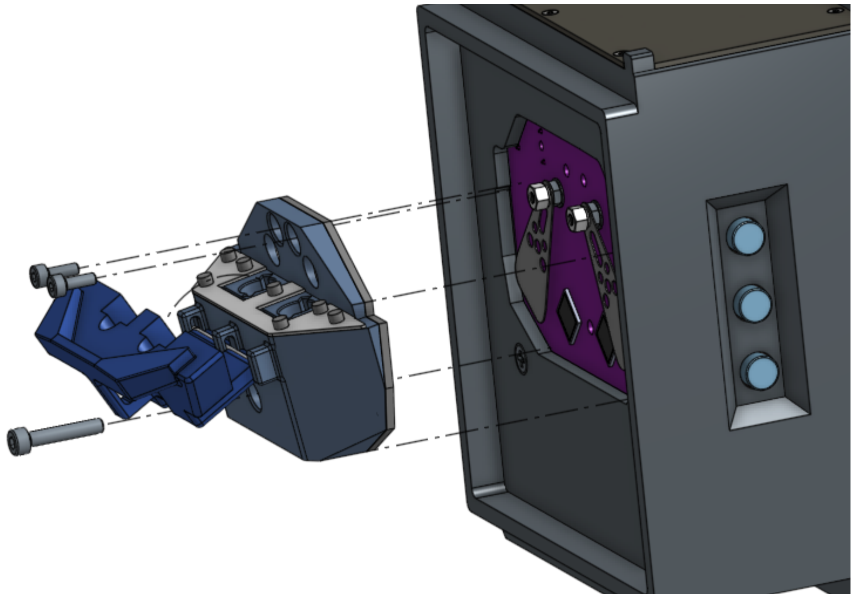

#### Installation of cover panels

1. Attach front/side panel 3-0861 using 4x #2-¼” long thread-forming screws (McMaster-Carr 95893A550).
    a. Front/side panel installed:

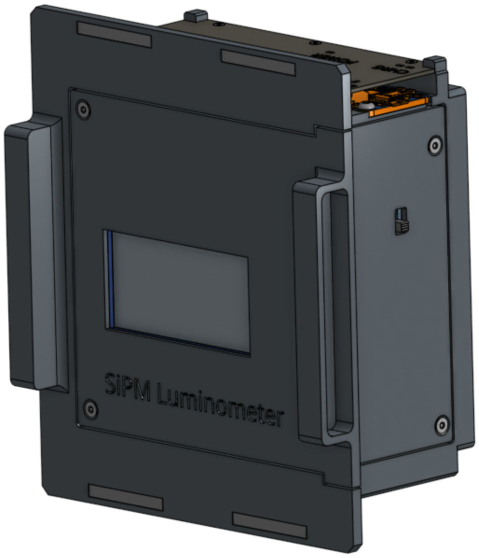


2. Install the powerboost cover panel using 6x #0-3/16” length thread-forming screws, (McMaster-Carr 95893A501)
    a. Powerboost cover panel installed:

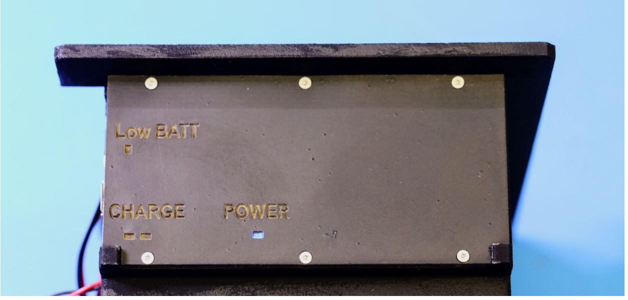


Finished!

#### Supplementary Photos

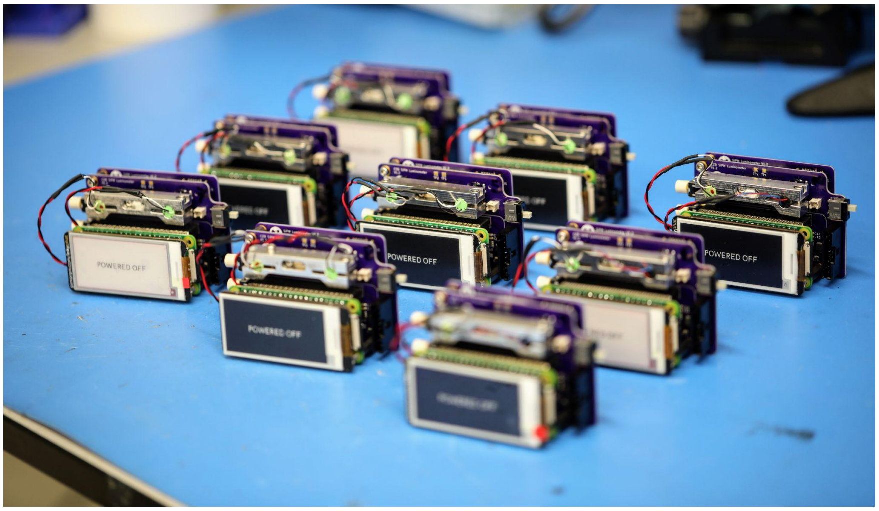

Fleet of assembled luminometer PCBs with shutters

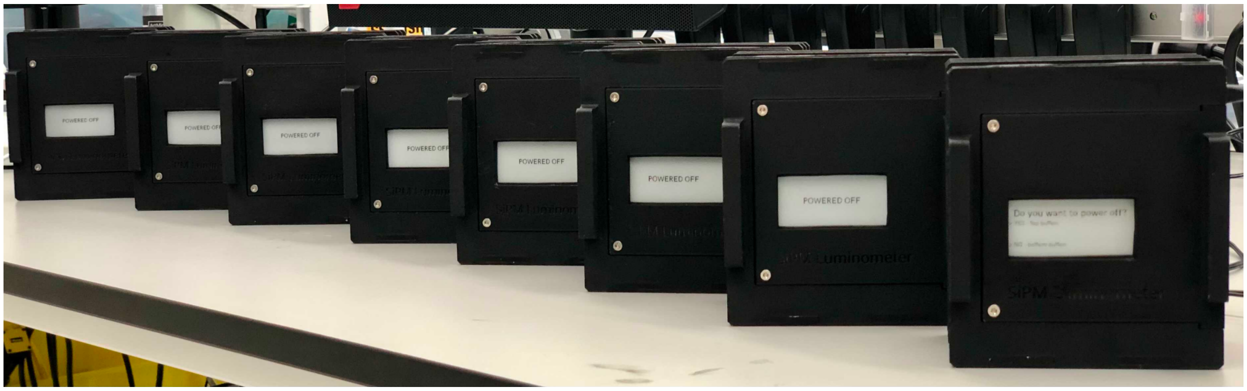


Eight completed luminometers.

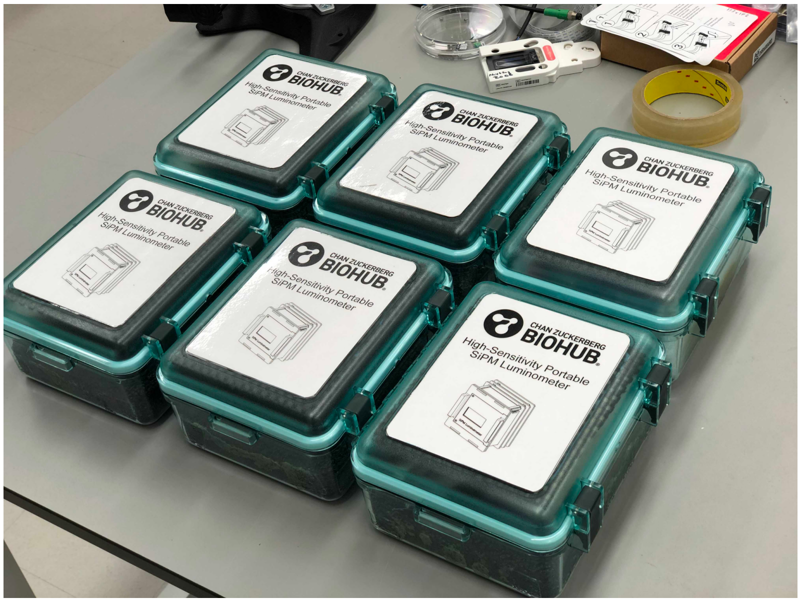


Portable luminometers in shock-resistant, waterproof carrying cases.
