## Supplementary material for "A handheld luminometer with sub-attomole limit of detection for distributed applications in global health": Luminometer user guide

### High-sensitivity Portable SiPM Luminometer

### User Guide

Last updated: September 22nd, 2021
Bioengineering Platform
Chan Zuckerberg Biohub | San Francisco, CA 94103

https://tinyurl.com/3p8p7axm

**Warnings** 3

**System Diagrams** 4

- Front view 4
- Exploded View 5
- Sample View 6
- User interface view 7

**Theory of Operation** 8

**Operation Guide** 10

- Common Actions and the Menu System 10
  - Action - charging the device 10
  - Action - powering the device off 11
    - Graphical Summary: Powering OFF 12
  - Action - powering the device ON 12
  - Action - waiting for the screen to update and pushing buttons 13
  - Action - pre-measurement checks 13
  - Action - performing a measurement 14
    - Graphical summary: Performing a measurement 16
  - Action - aborting a measurement-in-progress 16
  - Action - performing a temperature calibration 17
    - Graphical summary: Performing a temperature calibration 18
  - Action - choosing a calibration 19
  - Action - Viewing System Status 20

**Troubleshooting** 21

### Warnings

- These devices were designed and built at Chan Zuckerberg Biohub (CZB) as a tool for research purposes only, and provided free of cost to direct research collaborators.
- These devices are not for clinical use.
- These devices are not waterproof. Direct or indirect exposure to water may result in permanent damage or total failure of the device, as well as injury or harm to the user.
- Do not operate in direct sunlight. Although the design is intended to be light-proof, direct sunlight is bright enough to nonetheless cause problems with measurement.
- Do not let dirt, dust, or debris fall into the sample holder cavities.
- Each device contains a 4400 mA-hr / 3.7 V lithium polymer battery. Electrical shorting of any part of the instrument may result in the battery being shorted out, resulting in leaking and/or explosion/fire. Keep away from liquid and never short out components inside the device.
- There are no user-serviceable components inside the device. Users should never open the device to expose the internal electronics unless under the direct guidance of CZB staff.

### System Diagrams

#### Front view

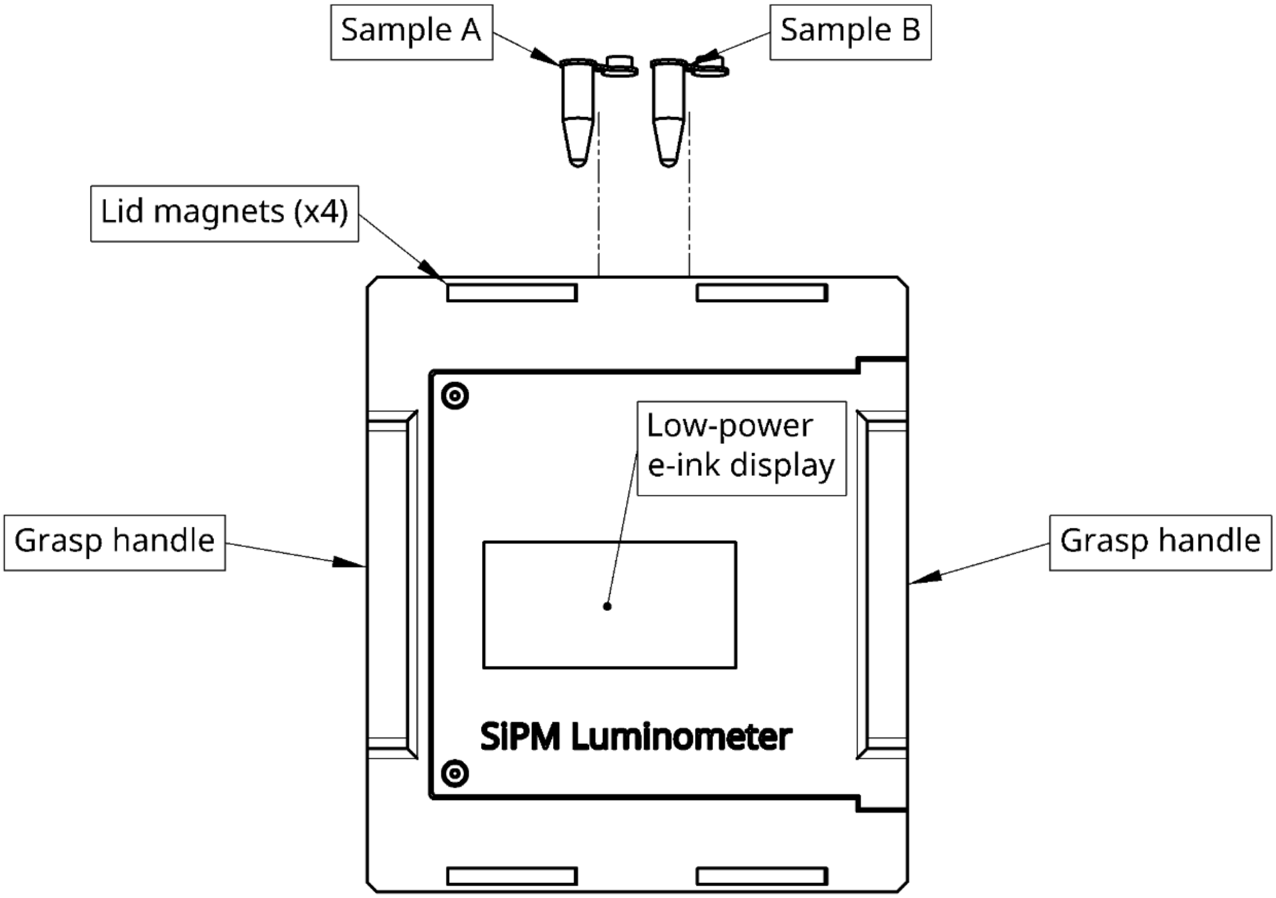


**Figure 1:** ***Front view.*** *The luminometer has two measurement channels. As viewed from the front, channel A is on the LEFT, and channel B is on the RIGHT. See Exploded View, Sample View, and User Interface View for details of operation*

#### Exploded View

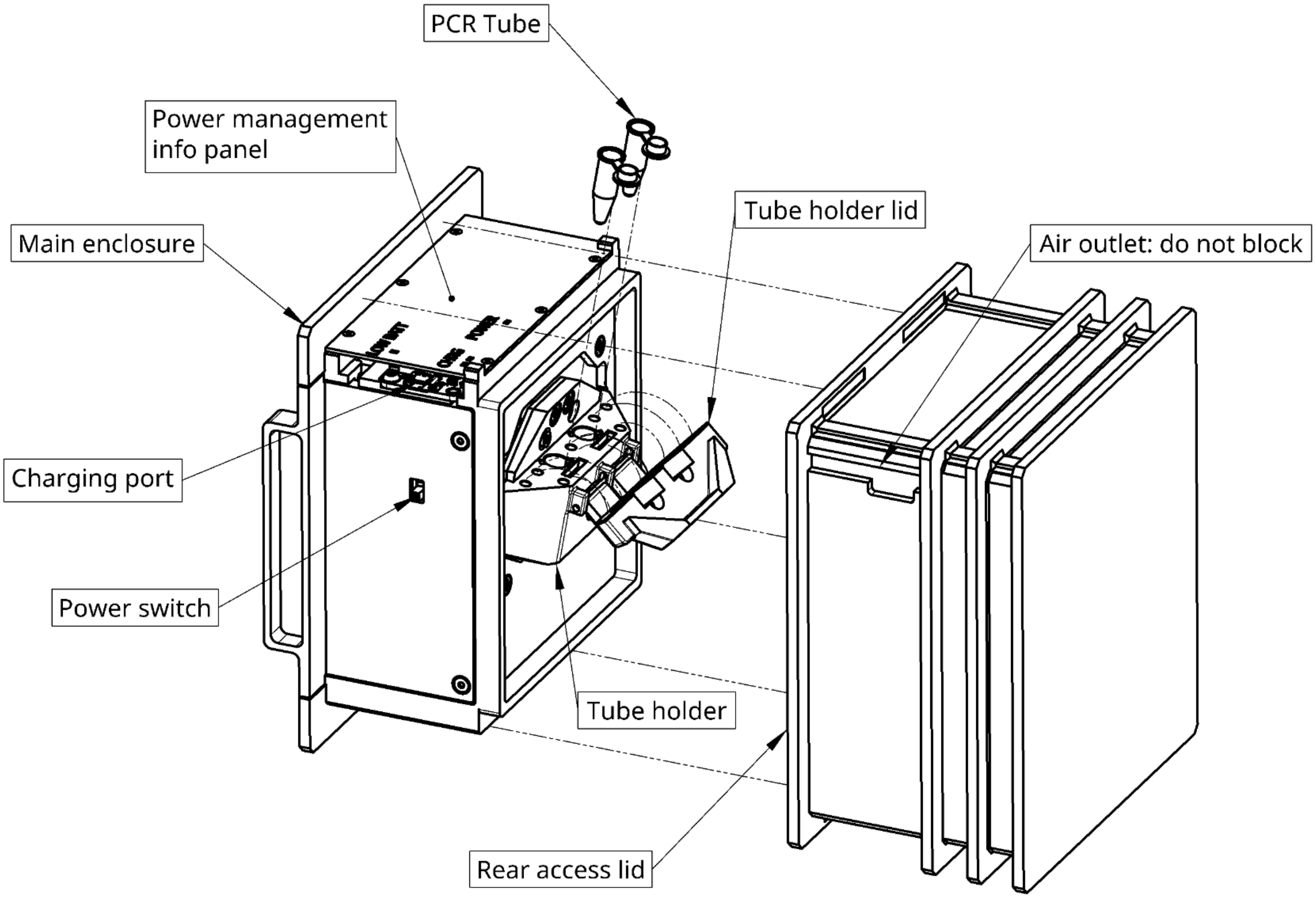


***Figure 2: Exploded view.*** *The luminometer consists of a main enclosure (left assembly) containing all the electronics, a small computer, battery, display, shutter mechanism, and sample interface. A rear access lid serves to light-proof as well as protect the assembly from damage. Once closed, the rear access lid is held in place magnetically. The tube holder houses PCR tubes for measurement, and includes a hinged lid which swings open and closed for sample access.*

#### Sample View

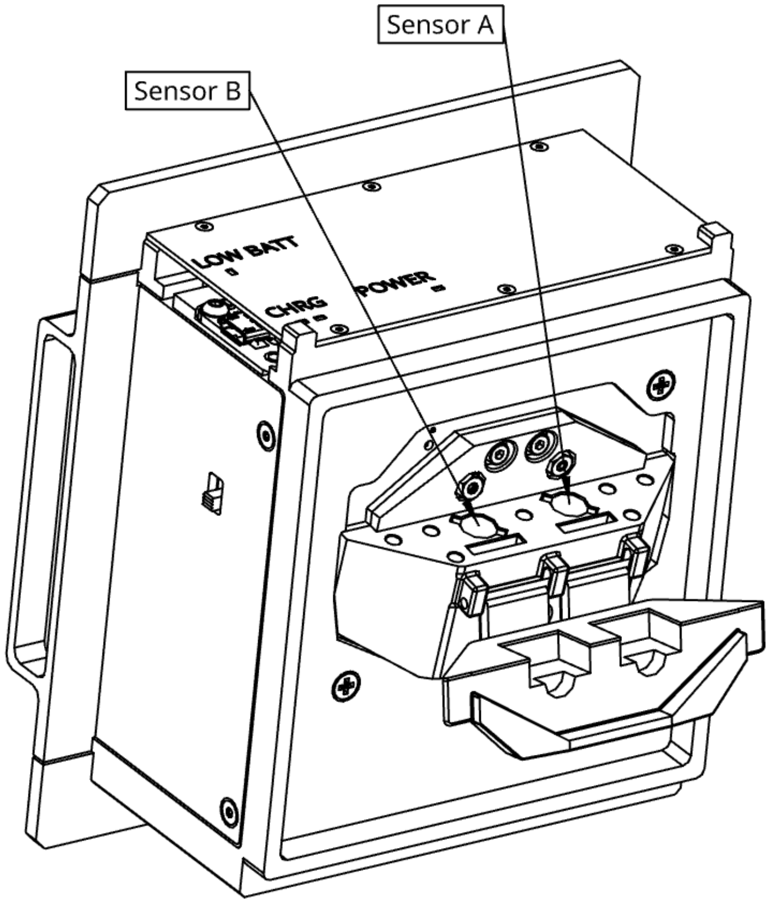


***Figure 3: Sample view.*** *The luminometer has two measurement channels: While facing the sample interface, sensor A is on the right, and sensor B is on the left.* ***WARNING! Never allow objects, dust, or debris to fall into the sensor cavities! Keep the tube holder lid close at all times, except when loading or unloading samples.***

#### User interface view

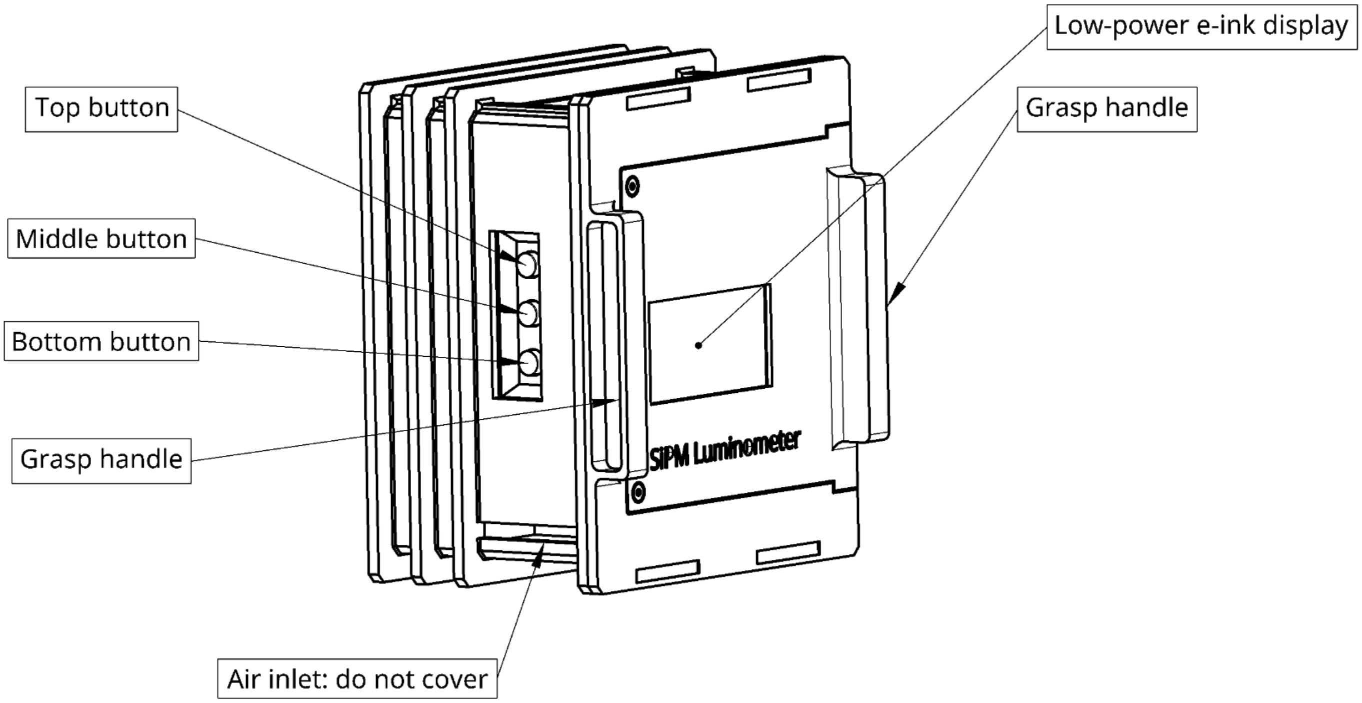


***Figure 4:*** *User interview view. The device has a minimalistic user interface consisting of three buttons and a low-power e-ink display.*

### Theory of Operation

This luminometer uses high-sensitivity Silicon Photomultiplier (SiPM) sensors to detect low levels of light. SiPM sensors consist of arrays of internal Avalanche Photodiodes (APDs) with extremely high internal gain, resulting in a large signal amplification for each photon that is detected. In other words, the sensor is able to produce large output signals from very tiny amounts of light. On short timescales, these sensors can even detect single photons.

One drawback of any light sensor is dark current -- that is, a signal that appears in the absence of light hitting the sensor. In practice, this means that all measurements we make consist of the actual luminescence signal (small) added on top of a dark offset signal (large). This dark signal depends directly on the temperature of the sensor, which is an uncontrolled parameter in the absence of temperature stabilization. Without mitigation, the uncontrolled drift of the dark signal would dwarf the tiny signals we are trying to measure. The dark signal can be thought of as being composed of two components: an average value that drifts slowly over time, and a residual random noise component that is fast, and averages to zero over time.

This luminometer is engineered for high-sensitivity luminescence applications, in which a slow, steady trickle of photons hits the sensor. The task is to distinguish a steady trickle from the dark offset and noise of the sensor itself, which requires integrating the signal for long enough to overcome the random noise of the dark signal. Unfortunately, dark signal drift prevents us from taking the most simple approach of just integrating the signal directly.

To compensate for the drift of the dark signal, a swinging shutter arm was introduced. The shutter repeatedly blocks and unblocks the luminescence light from hitting the sensor, allowing us to perform repeated dark offset measurements over time, as we accumulate enough signal from the sample. Each period of time the shutter is either open or closed, a series of data samples are acquired and averaged, giving us a sequence of measurements, each corresponding to alternating open and closed periods. To subtract the dark offset, each shutter open datapoint is subtracted by the mean of its flanking closed periods. Using both flanking closed periods fully compensates for linear drift in the dark signal. To achieve this, every measurement is programmed to start and end with shutter-closed periods. For example, if the total measurement time is set to 5 samples, then there will be 5 shutter-open periods and 6 shutter-closed periods: _-_-_-_-_-_, (where - represents open, and _ represents closed).

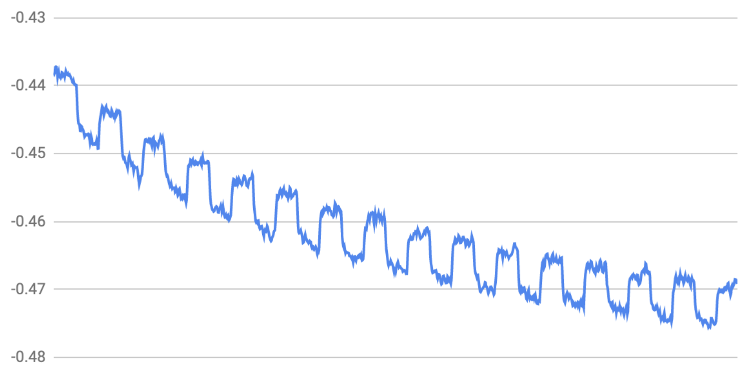


***Figure 5**: **raw signal trace (for example only, not displayed to the user)**. The shutter modulates the raw signal, allowing drift to be rejected. The user is provided with the mean demodulated signal value computed over the requested number of samples.*

After the subtraction is completed, the signal is now gated (Figure 4) and the averaged data points can themselves be averaged together to produce a single number. And because our final answer is the result of averaging multiple independent samples, the standard error of the mean can be reported as a statistical measure of the uncertainty due to counting statistics of dark current shot noise.

Finally, the sensors themselves were found to emit a tiny amount of light. This tiny amount of light reflects off the surfaces of the shutter, sample tube, and reflective cavity and is in turn detected again as a false signal. It was found that this false signal is also a function of temperature, and relates linearly to the dark current. We have strong empirical evidence that the sensor's dark current itself is responsible for the spurious emission of light. We are able to measure and model this effect, and compensate for it by using a calibration routine. As a result, the luminometer is almost entirely free of temperature-dependent measurement bias.

### Operation Guide

#### Common Actions and the Menu System

##### Action - charging the device

- It is important to fully charge the luminometer's battery prior to use. We recommend charging overnight, and powering down during periods without usage.
- Battery life:
    - The idle lifetime (no measurements performed) is approximately 10 hours.
    - The active lifetime (continuously measuring) is approximately 3 hrs.
    - In a typical use-case with intermittent measurements, we expect the device should last at least 5 hours.
- Charge the device by inserting the provided micro-USB into the slot (Figure 6). You can plug or unplug the device at any time. Power will not be interrupted to the device by plugging or unplugging. However, charging will complete more rapidly if the device is powered off.
- Once you plug in the charging cable, an orange LED will light up on the power board to indicate that the device is charging.
- When the battery is fully-charged, a green light on the power board will illuminate.
- When the battery is low, the red LED on the power board will illuminate.
- The red LED may begin to flash on and off with the shutter operation. The device can still be used while this occurs.
- If the red LED turns on continuously, the battery is low and the device must be charged.
- **Do not operate the luminometer if the red LED turns on continuously or if the device indicates LOW BATTERY. The data will not be reliable without sufficient battery power to operate the shutters.**

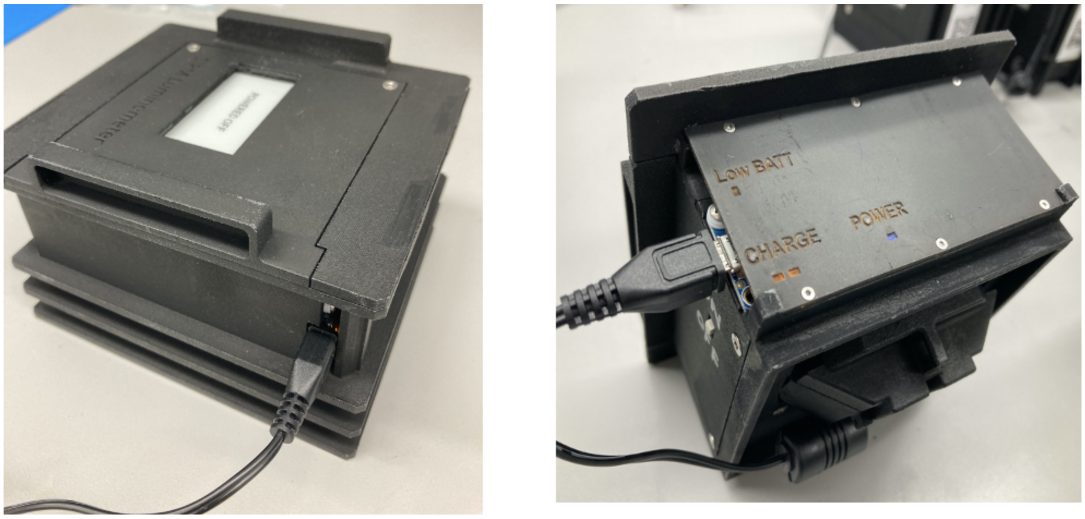


***Figure 6****: Location of the micro-USB port used to charge the luminometer.*

#### Action - powering the device off

(Graphical summary in Figure 8)

1. **Power down the software:**
    a. Press and hold the bottom button for 5 seconds from any screen to power off the software (except during a measurement)
2. **Tap the top button to confirm:**
    a. This will bring you to a confirmation screen (Figure 7):
        i. Press the top button to shut down the software (the screen will refresh and display "POWERED OFF"). You will hear the fan shut down. After shutting the software down, the blue LED on the power board will still remain on. The device is entering a low-power state.
        ii. If you change your mind and do not want to shut down, press the bottom button to cancel the shutdown and go back to the main menu.
3. **Wait 15 or more seconds.** The small computer inside the device needs 15 seconds to turn off. Similar to a desktop computer, you should not pull the plug while it is running. After 15 seconds, the device is now in a low-power state where the internal computer is off, but some of the electronics remain powered.
4. **Turn off the power switch to fully power off the device.**

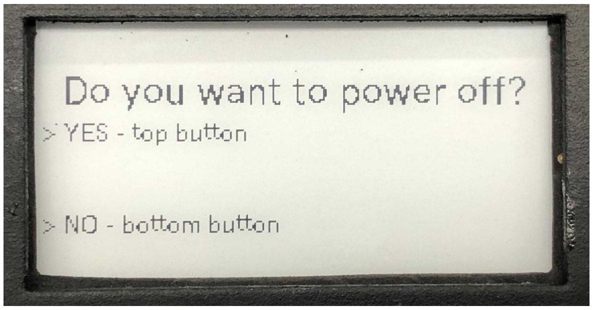


***Figure 7**: The confirmation screen for powering off the device.*

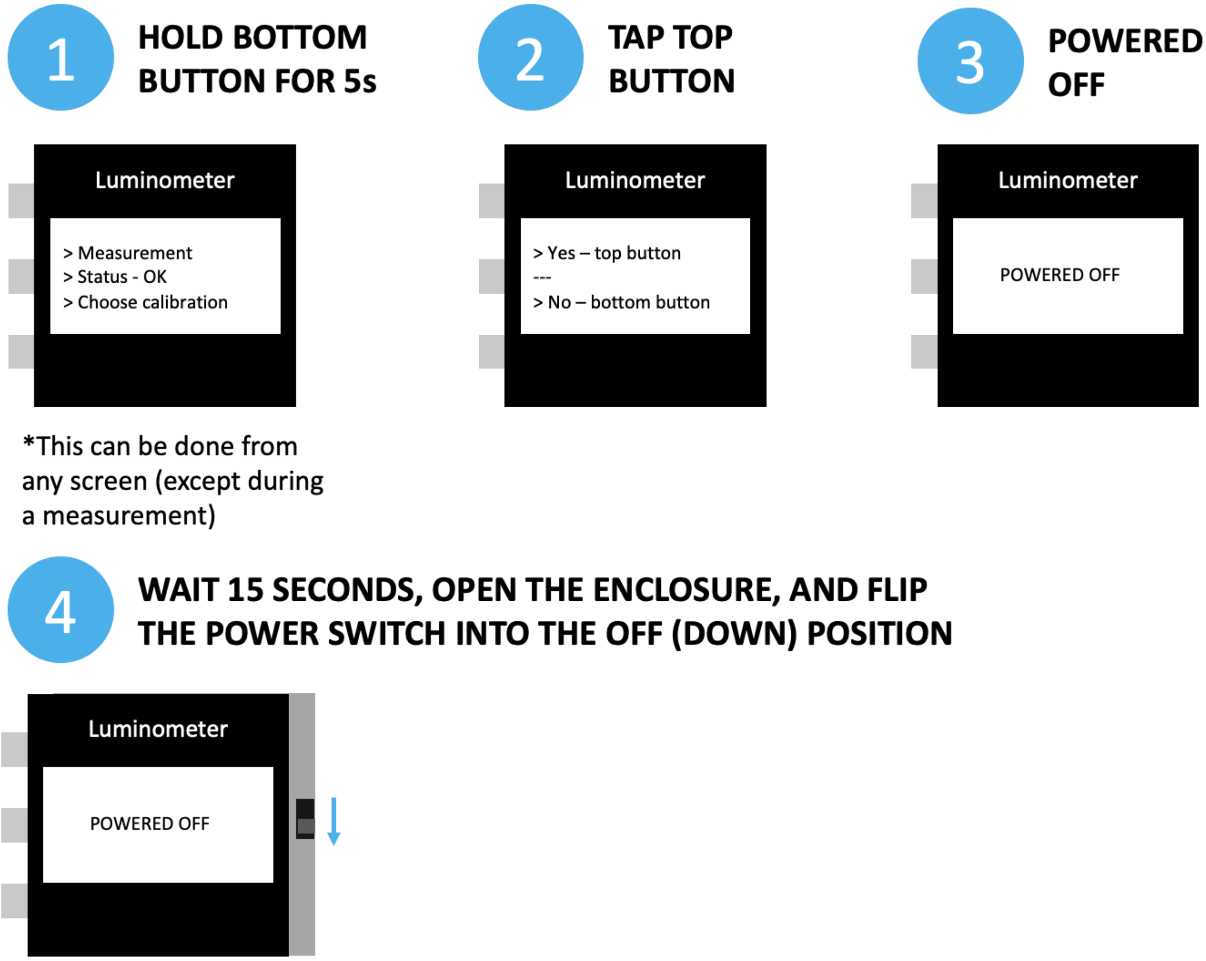


***Figure 8**: Graphical Summary for Powering OFF*

##### Action - powering the device ON

If the power switch is already in the 'OFF' position:

1. Simply slide the power to the 'ON' position and wait for 60-80 seconds.
2. You will start to hear the device fans turn on and the screen will flicker on and display the main menu.

If the power switch has been left 'ON' but the screen displays 'POWERED OFF', then the device has been left in the low-power state (software has shut down but the power switch was left 'ON'). To power on:

1. Cycle the power switch 'OFF' then 'ON'. **Do this only one time.**
2. The device will take **60-80 seconds** to power on.

##### Action - waiting for the screen to update and pushing buttons

- The device uses a low-power e-ink screen to save battery power.
- This screen has a refresh time of 5-6 seconds.
- After pressing a button input, *even if it appears that the screen has fully displayed, it may take another second before the device will register your next button press.*
- Pressing the button before the screen has fully settled will not do anything.
- A good rule is to allow the screen to fully display and then wait an additional 1 second before pressing a button.

##### Action - pre-measurement checks

- During measurements, always **keep the device on a flat surface, and UPRIGHT (screen vertical, Figure 9)**. Do not bump or disturb the device during measurements.
- When inserting a PCR tube, ensure it is pushed all the way down into the tube holder, such that the rim firmly contacts the top surface of the tube holder.
- Always use the exact recommended model of clear, thin-walled PCR tube. Changes to the make/model of PCR tube may alter the accuracy and/or precision of measurements due to differences in shape/size of the tube, which alter the optics of the measurement cavity. If absolutely necessary to change tubes, it is necessary to repeat the thermal compensation calibration. **Never use tinted or colored tubes**.

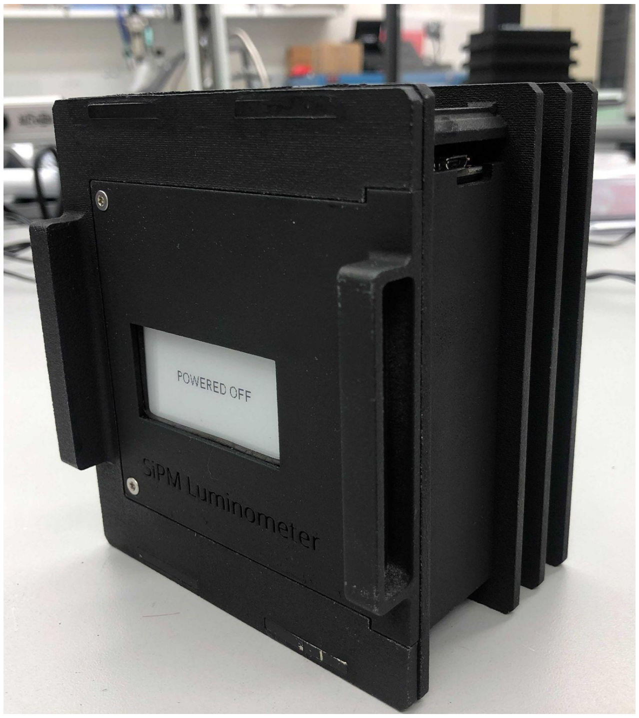


***Figure 9:*** *Always ensure the device is facing upright as shown, during a measurement*

#### Action - performing a measurement

(Graphical Summary in Fig 13)

**IMPORTANT: READ BEFORE USING**

- **Use only the ‘Timed’ mode. For the Spluc assay, an exposure of 30 samples is recommended. Do not use ‘Autoexposure’ mode at this time as it is experimental.**
- **Do not operate in direct sunlight. Direct sunlight is bright enough to cause problems with measurements due to light leakage. In general, be as consistent as possible with ambient lighting when making direct comparisons between samples.**

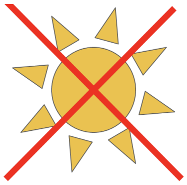

**Starting a measurement**

1. From the main menu screen, press the top button to enter the measurement screen.
2. From the measurement screen (Figure 10):
    a. Top button: perform an auto-exposure measurement
        i. The device will run for a minimum of 7 seconds, or until one of the following conditions is met:
            1. The Signal-To-Noise (SNR) has exceeded the target value (a hard-coded value). SNR is defined as the mean of all the gated shutter-open samples divided by the standard error of the mean of the same datapoints. The SNR improves over time, as the counting statistics improve with more integrated signal.
    b. Middle button (can be tapped or held. The device will beep for every second the button is pressed and held.):
        i. Tap the button to run a 5 sample measurement
        ii. Hold the button for 1s to run a 15 sample measurement (1 beep)
        **iii. Hold the button for 2s to run a 30 sample measurement (2 beeps)**
        iv. Hold the button for 3s to run a 150 sample measurement (3 beeps)
        v. Hold the button for 4s to run a 300 sample measurement (4 beeps)

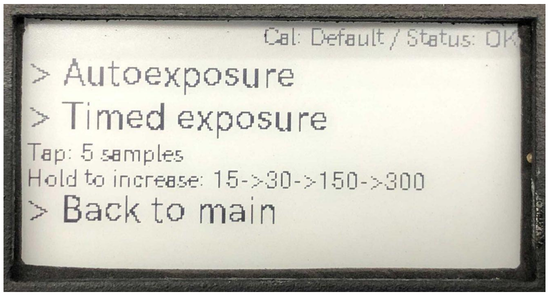


***Figure 10:*** *The measurement menu screen*

**During a measurement**

- The device will begin making a "tick-tock" sound as the shutters open and close across the sensor
- The screen will update every few seconds with the latest measurement results, Figure 11.
- The standard error of the mean (s.e.m.) estimate will initially begin at '+/- inf', indicating that at least 3 samples need to be collected prior to estimating the s.e.m..

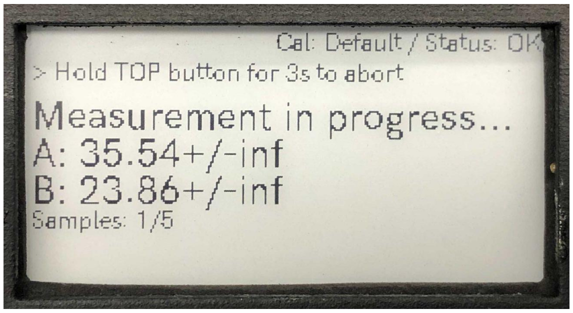


***Figure 11****: A 5 sample measurement in progress. Note that at the very beginning of a measurement, the screen may display '+/- inf'. This is expected and will go away as the measurement continues.*

- **Measurement complete**
  - Once the measurement is complete, the device will beep, the shutters will stop clicking, and the screen will update to display "Final: Yes" with the final measurement values, Figure 12.
  - **It is the operator's responsibility to record all measurements from the result displayed on the screen. Although an internal measurement log is kept on board the device, it is not accessible to the user via the button/screen interface.**
  - For convenience, pressing the middle button from the final measurement screen will repeat the measurement immediately. This is useful when taking multiple measurements in succession using the same number of sample measurements (i.e., 5, 15, 30, 150, or 300). Load in the new PCR tubes **then** press the "Redo measurement" middle button.

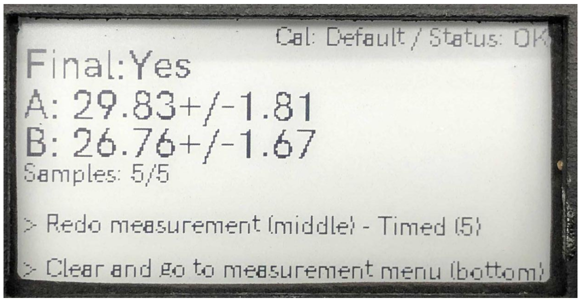


***Figure 12:*** *A completed 5 sample measurement*

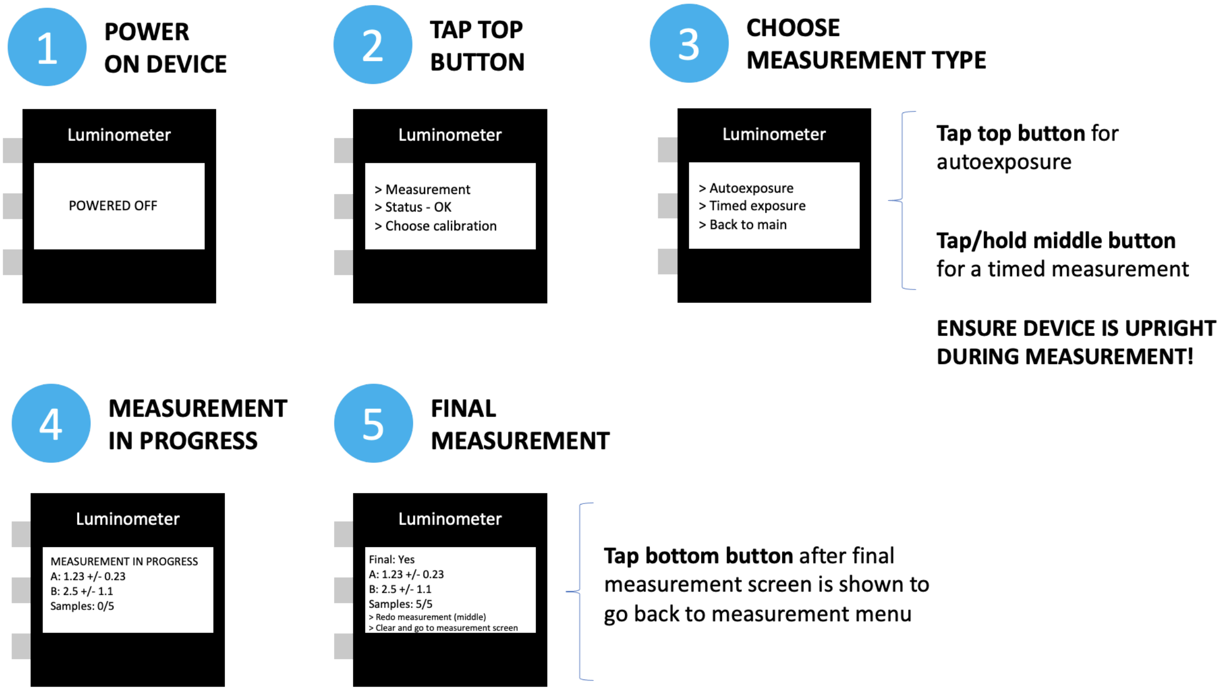


***Figure 13:*** *Graphical summary: Performing a measurement*

##### Action - aborting a measurement-in-progress

- Press and hold the top button for 3 seconds while a measurement is in progress to cancel it and return to the measurement screen.

##### Action - performing a temperature calibration

(Graphical Summary in Figure 16)

As described in the Theory of Operation, the device produces measurements that are fully independent of ambient temperature, by virtue of a calibration procedure. A “Default” calibration is stored on the device and is not altered by user-performed calibrations. The default calibration can always be restored by following the Action - choosing a calibration procedure.

The “Default” calibration was performed at CZ Biohub on a negative control sample prepared as described in the Protocols document.

In the event a custom calibration is required for measurements using a different assay, the “Custom” calibration feature may be used to store a custom calibration for future use.

NOTE: The device will only store one custom calibration, which will be overwritten each time the procedure is performed.

**Temperature calibration pre-flight checklist:**

- Ensure the luminometer has a fully-charged battery.
- Ensure you have access to a 4°C refrigerator.
- Ensure you have access to an incubator or oven, pre-heated to 37-40°C. Ambient temperature may be used, so long as the ambient temperature is the same or higher than will be used during experiments, and is at a minimum temperature of 35°C.
- Two PCR tubes, each filled with a negative control sample, using **exactly** the same volume of liquid as will be used for the experiments, and prepared exactly the same way as experimental tubes.
- The negative controls should have exactly the same composition as the real samples, but with no light emission (ex: no luciferase control).
- The negative control samples should be prepared in exactly the same manner as the experiment. For example, if there is a centrifugation step to pellet cells, perform this step in the same manner as real samples.

**Running the calibration**

1. Power the device ON. It will enter the main menu.
2. Place the device in a 37-40°C incubator for 1 hr.
3. Navigate to the calibration menu by tapping the bottom button while on the main menu, Figure 14.

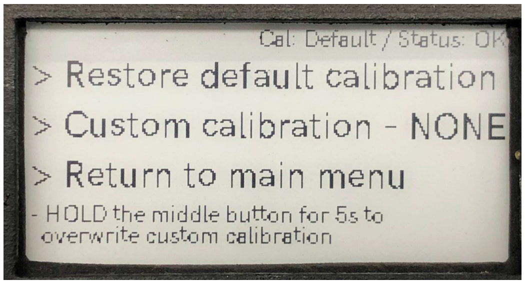


***Figure 14:*** *The calibration menu*

4. Choose to start a custom calibration by holding the middle button down for 5 seconds.
    a. Ensure your negative control sample is prepared.
    b. Ensure the liquid is at the bottom of the tubes, and is totally free of bubbles, droplets, or other abnormalities.
    c. Place the negative control samples inside the tube holder of the device. The calibration will run on both channels at the same time.
5. **STOP** - are you sure you want to overwrite the custom calibration (Figure 15)?
    a. If yes, confirm the start of the routine by tapping the top button.

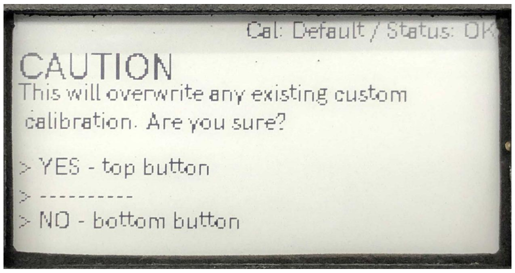


**Figure 15:** A warning screen before overwriting the calibration data

6. Immediately transfer the device into a 4C refrigerator leaving it upright and motionless until the Final Measurement screen is displayed. This will take approximately 40 minutes.
    a. Once the custom calibration measurement is running you will hear the shutters clicking. The screen will display a "Measurement in progress" screen.
7. Upon completion, the device will display a "Final: yes" screen. You can now return to the main menu. The temperature calibration is complete and has been saved to the device.
    a. Note: The final measurement at the end of the calibration is meaningless and can be ignored.
8. **Important: It is essential that after removing the device from the refrigerator that it is powered down due to condensation of ambient humidity that will occur. The humidity may damage internal electronics if they are powered on. Do not power the device back on until it has spent >1 hr at ambient temperature.**

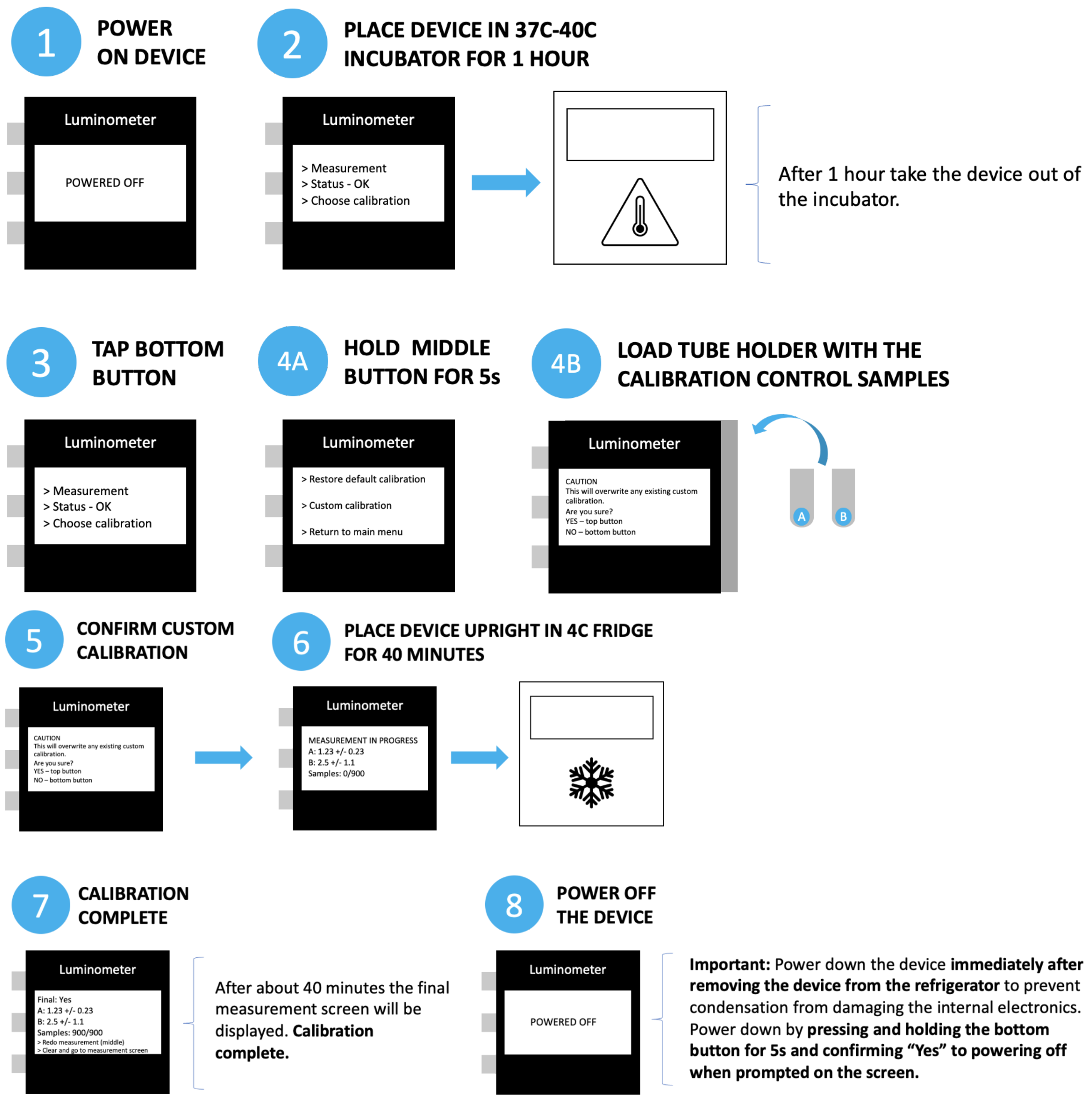


***Figure 16:*** *Graphical summary: Performing a temperature calibration*

##### Action - choosing a calibration

- From the main menu, tap the bottom button to enter the calibration screen. The following options will appear:
    - **(Top button) Restore default calibration -** this will reset the temperature calibration values to their factory values
    - **(Middle button) Custom calibration -** if a custom calibration has already been done, this will set the temperature calibration values to the custom ones. **Note:** if no calibration has been done yet, this line will display "Custom calibration - NONE" (pressing the middle button when it displays this line will simply return you to the main menu).
    - **(Bottom button) Return to main menu**

##### Action - Viewing System Status

- From the main menu screen, tap the middle button to enter the status/diagnostic screen.
- See Figure 17 for typical values displayed on the screen

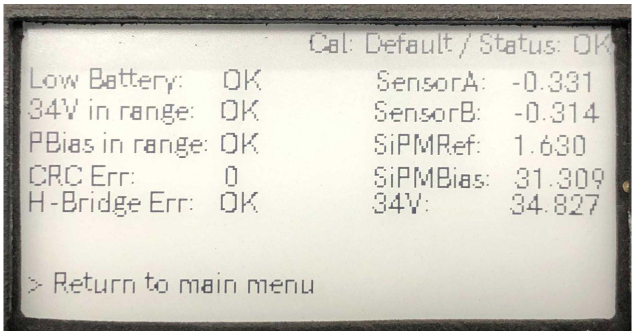


***Figure 17:*** *The device status screen showing normal values*

- The left column displays OK/ERR status for certain elements of the electronics:
    - Low battery: shows battery charge status
    - 34V in range: shows whether the sensor's power supply is operational
    - PBias in range: shows whether the photodetector bias voltage is within the correct range.
    - Cyclic Redundancy Check (CRC) error count: displays the number of communication errors that have occurred in the most recent measurement. If this number is non-zero there is not necessarily a problem with the device. However, if the CRC error level becomes too high, an error will be raised and shown on the main menu.
    - H-Bridge Err: Indicates the status of the shutter driver chip.
- The right column displays numerical values for several system parameters:

  - SensorA: Raw voltage from sensor A. If the luminometer is closed up with no sample, this should read somewhere in the range -0.34 V to 0.1 V, depending on ambient temperature. If the rear access lid or the tube holder is not fully-secured, these values will change.
  - SensorB: Raw voltage from sensor B. If the luminometer is closed up with no sample, this should read somewhere in the range -0.34 V to 0.1 V, depending on ambient temperature. If the rear access lid or the tube holder is not fully-secured, these values will change.
  - SiPM Ref: The silicon photomultiplier reference voltage should be between 1.62 V and 1.64 V.
  - SiPMBias: The silicon photomultiplier bias voltage should be between 30 V and 32 V.
  - 34V: The power supply feeding the SiPMBias system.
    - **NOTE:** If the rear access lid and/or the tube holder is not fully closed, these values may be pulled into an invalid range due to excessive light hitting the sensors (Figure 18). If an invalid value is observed for any of the above parameters, try closing the device up and re-entering the menu. If the SiPMRef is still out of range, please contact CZ Biohub.

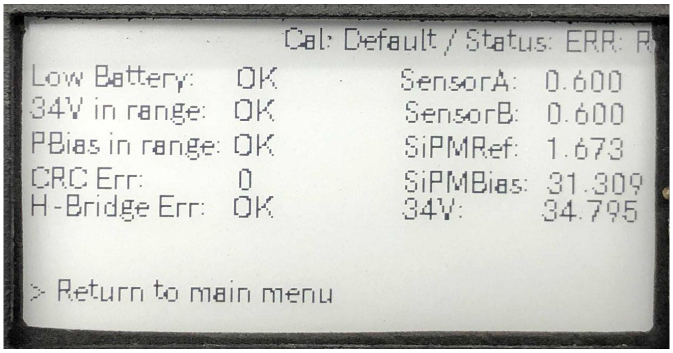


***Figure 18:*** *Error status triggered by excessive light hitting the sensors.*

- From the status screen, press the bottom button to return to the main menu.

### Troubleshooting

*ssh instructions:*

- ssh pi@freshlumi{NUM}.local (e.g pi@freshlumi1.local)
- pw: fresh1 (for all luminometers)
