## Supplementary material for "A handheld luminometer with sub-attomole limit of detection for distributed applications in global health": Mirzapur study data

| id | spluc_date | lumi_number | S1 | S1_err | S2 | S2_err | N1 | N1_err | N2 | N2_err | C1 | C1_err | C2 | C2_err | CTRL Avg | S_avg | N_avg | vax_date_1 | vax_date_2 | vax_date_3 | inf_date_1 | inf_date_2 | inf_date_3 |
|---|---|---|---|---|---|---|---|---|---|---|---|---|---|---|---|---|---|---|---|---|---|---|---|
| 1001 | 12-May-22 | 7 | 332.6 | 2.9 | 702.5 | 2.5 | 72.7 | 1.4 | 92.3 | 3.1 | 35.1 | 1.2 | 66.4 | 1.3 | 50.7 | 517.6 | 82.5 | 3/28/2021 | 1/10/2022 | 4/11/2022 | 7/5/2021 | 1/22/2022 | |
| 1002 | 12-May-22 | 7 | 152.6 | 2 | 180.8 | 2 | 376.3 | 2.3 | 407.5 | 2.9 | 35.1 | 1.2 | 66.4 | 1.3 | 50.7 | 166.7 | 391.9 | 3/4/2021 | 5/2/2021 | 3/5/2022 | 1/19/2022 | 6/2/2020 | |
| 1003 | 12-May-22 | 7 | 79.2 | 1.9 | 138.8 | 2.5 | -1.9 | 1.3 | 2.9 | 1.3 | 35.1 | 1.2 | 66.4 | 1.3 | 50.7 | 109 | 0.5 | 4/13/2021 | 6/4/2021 | 1/22/2022 | 11/23/2020 | | |
| 1004 | 12-May-22 | 7 | 75.2 | 1.7 | 61.2 | 1.6 | 180.7 | 1.9 | 164.2 | 2.1 | 35.1 | 1.2 | 66.4 | 1.3 | 50.7 | 68.2 | 172.5 | 9/4/2021 | 10/31/2021 | | 1/31/2022 | | |
| 1005 | 12-May-22 | 7 | 1306.8 | 5.4 | 994.7 | 3.6 | 29.4 | 2.1 | 18 | 0.9 | 35.1 | 1.2 | 66.4 | 1.3 | 50.7 | 1150.7 | 23.7 | 7/18/2021 | 8/18/2021 | 3/12/2022 | 1/23/2022 | | |
| 1006 | 18-May-22 | 7 | 401.3 | 1.9 | 468 | 3.1 | 32 | 1.6 | 45.4 | 1.5 | 53.4 | 1.5 | 50.4 | 1.2 | 51.9 | 434.7 | 38.7 | 2/10/2021 | 4/8/2021 | 1/5/2022 | 1/24/2022 | | |
| 1007 | 18-May-22 | 7 | 464.9 | 2 | 489.9 | 2.1 | 250.7 | 1.7 | 479.4 | 2.6 | 53.4 | 1.5 | 50.4 | 1.2 | 51.9 | 477.4 | 365 | 2/11/2021 | 4/10/2021 | | 6/15/2020 | 5/11/2021 | 1/25/2022 |
| 1008 | 18-May-22 | 7 | 720 | 2.6 | 533.1 | 2.9 | 453 | 2 | 379.4 | 1.5 | 53.4 | 1.5 | 50.4 | 1.2 | 51.9 | 626.6 | 416.2 | 9/13/2021 | 10/11/2021 | | | | |
| 1009 | 31-May-22 | 7 | 265.1 | 1.9 | 272.3 | 1.7 | -0.4 | 1.9 | 11.7 | 2.8 | 32.6 | 1.4 | 46.5 | 2.4 | 39.6 | 268.7 | 5.6 | 2/11/2021 | 4/10/2021 | 1/6/2022 | 5/23/2020 | 8/4/2020 | |
| 1010 | 31-May-22 | 7 | 213.7 | 1.5 | 182.1 | 1.6 | 3 | 1.8 | 5.1 | 2.1 | 32.6 | 1.4 | 46.5 | 2.4 | 39.6 | 197.9 | 4.1 | 2/11/2021 | 4/13/2021 | 1/5/2022 | 7/15/2020 | 7/1/2021 | |
| 1011 | 31-May-22 | 7 | 107.9 | 1.5 | 112.5 | 2 | 306.2 | 2.1 | 395.9 | 2 | 32.6 | 1.4 | 46.5 | 2.4 | 39.6 | 110.2 | 351.1 | 8/18/2021 | 9/20/2021 | 4/12/2022 | | | |
| 1012 | 31-May-22 | 7 | 189.1 | 2.3 | 177.2 | 2.7 | 274.8 | 2.1 | 268.8 | 2.2 | 32.6 | 1.4 | 46.5 | 2.4 | 39.6 | 183.2 | 271.8 | 2/14/2021 | 4/13/2021 | | 1/29/2022 | | |
| 1013 | 31-May-22 | 7 | 154.5 | 2.2 | 194.4 | 4.1 | -5.1 | 2.4 | -1.4 | 1.7 | 32.6 | 1.4 | 46.5 | 2.4 | 39.6 | 174.4 | -3.3 | 2/13/2021 | 5/12/2021 | 1/11/2022 | 11/3/2020 | | |
| 1014 | 31-May-22 | 7 | 2701.6 | 3.3 | 2657.6 | 3.2 | 192.6 | 1.5 | 193.8 | 1.7 | 32.6 | 1.4 | 46.5 | 2.4 | 39.6 | 2679.6 | 193.2 | 8/1/2021 | 9/1/2021 | 5/15/2022 | 7/23/2020 | | |
| 1015 | 31-May-22 | 7 | 349.7 | 2.1 | 370 | 2 | -1 | 1.7 | 5.1 | 1.6 | 32.6 | 1.4 | 46.5 | 2.4 | 39.6 | 359.8 | 2.1 | 4/3/2021 | 5/30/2021 | 1/11/2022 | 1/15/2021 | | |
| 1016 | 31-May-22 | 7 | 301.5 | 2.2 | 458.2 | 2.6 | -4.8 | 1.9 | 0.6 | 1.8 | 32.6 | 1.4 | 46.5 | 2.4 | 39.6 | 379.8 | -2.1 | 4/15/2021 | 6/15/2021 | 1/10/2022 | 4/24/2021 | | |
| 1017 | 13-Jun-22 | 7 | 224.4 | 1.7 | 238 | 3 | 62.6 | 1.1 | 74 | 1.5 | 30.5 | 1.1 | 32.8 | 1.6 | 31.6 | 231.2 | 68.3 | 2/11/2021 | 4/11/2021 | 1/20/2022 | 6/24/2021 | | |
| 1018 | 13-Jun-22 | 7 | 76.7 | 1.5 | 113.9 | 1.3 | 34.7 | 1.7 | 49.6 | 1.6 | 30.5 | 1.1 | 32.8 | 1.6 | 31.6 | 95.3 | 42.1 | 7/29/2021 | 9/2/2021 | | 1/15/2022 | | |
| 1019 | 13-Jun-22 | 7 | 322.2 | 1.5 | 280.2 | 1.7 | 744.3 | 4.5 | 925.6 | 6.3 | 30.5 | 1.1 | 32.8 | 1.6 | 31.6 | 301.2 | 834.9 | 3/25/2021 | 5/22/2022 | | 10/15/2020 | 7/5/2021 | |
| 1020 | 13-Jun-22 | 7 | 108.4 | 1.4 | 126.5 | 1.8 | 165.9 | 2.1 | 150.5 | 2.7 | 30.5 | 1.1 | 32.8 | 1.6 | 31.6 | 117.4 | 158.2 | 2/19/2021 | 3/31/2022 | | 1/15/2022 | | |
| 1021 | 13-Jun-22 | 7 | 476.8 | 1.3 | 463.9 | 1.7 | 54.8 | 2 | 54.4 | 3.2 | 30.5 | 1.1 | 32.8 | 1.6 | 31.6 | 470.3 | 54.6 | 7/13/2021 | 8/16/2021 | 4/21/2021 | 3/23/2021 | | |
| 1022 | 13-Jun-22 | 7 | 497.3 | 2.2 | 520.6 | 3.2 | 236.5 | 2.1 | 193.1 | 1.5 | 30.5 | 1.1 | 32.8 | 1.6 | 31.6 | 508.9 | 214.8 | 2/23/2021 | 4/24/2021 | 3/5/2022 | 7/25/2020 | 1/29/2022 | 5/5/2022 |
| 1023 | 13-Jun-22 | 7 | 142.9 | 1.8 | 203.6 | 1.6 | 71.5 | 2.1 | 100.6 | 1.5 | 30.5 | 1.1 | 32.8 | 1.6 | 31.6 | 173.3 | 86 | 2/16/2021 | 4/15/2021 | 3/3/2022 | 7/29/2020 | 1/20/2022 | |
| 1024 | 13-Jun-22 | 7 | 324.3 | 2.6 | 353 | 1.8 | 7.3 | 1.5 | 16.9 | 1.5 | 30.5 | 1.1 | 32.8 | 1.6 | 31.6 | 338.7 | 12.1 | 2/16/2021 | 5/19/2021 | 2/22/2022 | 9/28/2021 | 4/19/2020 | |
| 1025 | 13-Jun-22 | 7 | 14.7 | 2 | 21.2 | 1.8 | 469.9 | 1.5 | 423.7 | 1.6 | 30.5 | 1.1 | 32.8 | 1.6 | 31.6 | 18 | 446.8 | | | | | | |
| 1026 | 13-Jun-22 | 7 | 1186.1 | 2 | 1288.1 | 2.7 | 129.7 | 1.7 | 151.1 | 1.7 | 30.5 | 1.1 | 32.8 | 1.6 | 31.6 | 1237.1 | 140.4 | 7/13/2021 | 8/16/2021 | 3/30/2022 | 4/28/2020 | 8/20/2020 | 1/25/2022 |
| 1027 | 13-Jun-22 | 7 | 1237.5 | 3 | 1680.5 | 3.4 | 49.5 | 2.2 | 71.6 | 1.5 | 30.5 | 1.1 | 32.8 | 1.6 | 31.6 | 1459 | 60.6 | 10/9/2021 | 11/20/2021 | 4/12/2022 | 1/16/2022 | | |
| 1028 | 13-Jun-22 | 7 | 153.6 | 1.5 | 188.9 | 2.5 | 16.4 | 1.7 | 30.8 | 2 | 30.5 | 1.1 | 32.8 | 1.6 | 31.6 | 171.2 | 23.6 | 4/5/2021 | 6/10/2021 | 3/14/2022 | 6/15/2020 | 1/15/2022 | |
| 1029 | 18-Jun-22 | 7 | 30.9 | 2 | 42.1 | 2.3 | 4 | 2 | 11.1 | 2.8 | 29.6 | 1.7 | 42.2 | 1.7 | 35.9 | 36.5 | 7.5 | 2/16/2021 | 3/18/2021 | | 7/4/2021 | | |
| 1030 | 18-Jun-22 | 7 | 172 | 1.8 | 124.2 | 2.6 | 1117.2 | 2.1 | 1146.4 | 2 | 29.6 | 1.7 | 42.2 | 1.7 | 35.9 | 148.1 | 1131.8 | 8/16/2021 | 9/15/2021 | 3/22/2022 | 9/1/2020 | 3/15/2021 | 1/20/2022 |
| 1031 | 18-Jun-22 | 7 | 564.8 | 1.8 | 713.5 | 2.3 | 35.9 | 1.6 | 48.9 | 3.1 | 29.6 | 1.7 | 42.2 | 1.7 | 35.9 | 639.1 | 42.4 | 7/14/2021 | 8/16/2021 | 3/22/2022 | 7/21/2020 | 3/21/2021 | |
| 1032 | 18-Jun-22 | 7 | 2502.6 | 3.5 | 2087.4 | 3.6 | 1484.3 | 2.1 | 1195 | 3 | 29.6 | 1.7 | 42.2 | 1.7 | 35.9 | 2295 | 1339.6 | 2/15/2021 | 4/15/2021 | 3/24/2022 | 4/15/2020 | 7/15/2020 | 1/16/2022 |
| 1033 | 18-Jun-22 | 7 | 130.1 | 1.5 | 141.6 | 2.1 | 280.3 | 2.1 | 324.3 | 2.7 | 29.6 | 1.7 | 42.2 | 1.7 | 35.9 | 135.9 | 302.3 | 3/15/2021 | 5/6/2021 | | 7/3/2021 | 1/30/2022 | |
| 1034 | 18-Jun-22 | 7 | 553.3 | 1.6 | 544.7 | 1.5 | 57.4 | 2 | 76.2 | 2.9 | 29.6 | 1.7 | 42.2 | 1.7 | 35.9 | 549 | 66.8 | 8/5/2021 | 9/9/2021 | 3/31/2022 | 5/4/2020 | | |
| 1035 | 18-Jun-22 | 7 | 143.3 | 1.7 | 145.3 | 2.2 | 918 | 3.7 | 951.1 | 3.4 | 29.6 | 1.7 | 42.2 | 1.7 | 35.9 | 144.3 | 934.6 | | | | 6/7/2020 | 1/26/2022 | |
| 1036 | 18-Jun-22 | 7 | 124 | 1.9 | 213.4 | 1.4 | 2 | 1.5 | 20.6 | 1.2 | 29.6 | 1.7 | 42.2 | 1.7 | 35.9 | 168.7 | 11.3 | 2/13/2021 | 4/15/2021 | 1/10/2022 | 7/3/2021 | | |
| 1037 | 18-Jun-22 | 7 | 808 | 3.5 | 1056.8 | 3.5 | 226.7 | 2.3 | 195.6 | 2.3 | 29.6 | 1.7 | 42.2 | 1.7 | 35.9 | 932.4 | 211.1 | 4/5/2021 | 6/3/2021 | 3/14/2022 | 6/21/2021 | 1/23/2022 | |
| 1038 | 19-Jun-22 | 7 | 41.2 | 1.1 | 35.6 | 1.8 | -1.4 | 1.3 | 13.4 | 1.7 | 30.6 | 1.2 | 28.3 | 1.2 | 29.5 | 38.4 | 6 | 2/17/2021 | 5/17/2021 | 1/12/2022 | 7/15/2020 | 9/29/2020 | 7/17/2021 |
| 1039 | 19-Jun-22 | 7 | 124.6 | 1.8 | 128.3 | 1.5 | 4.9 | 1.6 | 14.6 | 2 | 30.6 | 1.2 | 28.3 | 1.2 | 29.5 | 126.4 | 9.7 | 2/19/2021 | 4/20/2021 | 1/4/2022 | 3/15/2021 | | |
| 1040 | 19-Jun-22 | 7 | 172.9 | 1.5 | 209.7 | 2.7 | 146.2 | 2 | 161.7 | 2.2 | 30.6 | 1.2 | 28.3 | 1.2 | 29.5 | 191.3 | 153.9 | 7/4/2021 | 8/4/2021 | 3/13/2022 | | | |
| 1042 | 19-Jun-22 | 7 | 272.7 | 1.6 | 223.9 | 2.7 | 17.2 | 1.1 | 21.7 | 2.9 | 30.6 | 1.2 | 28.3 | 1.2 | 29.5 | 248.3 | 19.5 | 2/15/2021 | 4/13/2021 | 1/20/2022 | | | |
| 1043 | 19-Jun-22 | 7 | 838.2 | 2.4 | 1055.9 | 4.2 | 214.5 | 1.3 | 244.8 | 2 | 30.6 | 1.2 | 28.3 | 1.2 | 29.5 | 947 | 229.7 | 11/17/2021 | 12/15/2021 | 4/20/2022 | 10/24/2020 | 1/19/2022 | |
| 1044 | 19-Jun-22 | 7 | 268.2 | 1.8 | 292.6 | 2.7 | 463.5 | 1.7 | 496.6 | 2.7 | 30.6 | 1.2 | 28.3 | 1.2 | 29.5 | 280.4 | 480.1 | 2/15/2021 | 4/13/2021 | 1/20/2022 | 8/10/2020 | 1/26/2022 | |
| 1045 | 20-Jul-22 | 7 | 73.3 | 1.4 | 59.8 | 2.4 | 1127.4 | 3 | 918.6 | 3.4 | 25.5 | 1.3 | 38.9 | 1.6 | 32.2 | 66.5 | 1023 | 10/7/2021 | 11/18/2021 | | 1/15/2022 | | |
| 1046 | 20-Jul-22 | 7 | 785.3 | 2.5 | 1027.5 | 2.7 | 165.5 | 1.9 | 138.4 | 2.4 | 25.5 | 1.3 | 38.9 | 1.6 | 32.2 | 906.4 | 151.9 | 8/23/2021 | 9/20/2021 | 3/31/2022 | | | |
| 1047 | 20-Jul-22 | 7 | 434.8 | 1.7 | 485.9 | 1.5 | 105.5 | 2 | 11.3 | 2.2 | 25.5 | 1.3 | 38.9 | 1.6 | 32.2 | 460.3 | 58.4 | 11/18/2021 | 12/20/2021 | 5/9/2022 | 7/28/2020 | 1/17/2022 | |
| 1048 | 20-Jul-22 | 7 | 219.5 | 2.2 | 208.3 | 2.2 | 5248.2 | 14.7 | 6425.1 | 14.2 | 25.5 | 1.3 | 38.9 | 1.6 | 32.2 | 213.9 | 5836.6 | 2/20/2021 | 4/22/2021 | 1/11/2022 | 12/15/2020 | 6/24/2020 | |
| 1049 | 20-Jul-22 | 7 | 388.1 | 1.8 | 415 | 2.4 | 2178.3 | 4.6 | 2930.1 | 5.7 | 25.5 | 1.3 | 38.9 | 1.6 | 32.2 | 401.6 | 2554.2 | 1/22/2021 | 5/17/2021 | 1/13/2022 | 4/27/2020 | | |
| 2001 | 27-Jun-22 | 1 | 9 | 1.3 | 20.8 | 1.3 | 19.4 | 1.7 | 15.8 | 1 | 34.5 | 1.4 | 49.5 | 1.3 | 42 | 14.9 | 17.6 | 2/11/2021 | 3/11/2021 | | 1/24/2022 | | |
| 2002 | 27-Jun-22 | 1 | 11.1 | 1.1 | 10.3 | 1.4 | 35.3 | 2.6 | 73.4 | 1.4 | 34.5 | 1.4 | 49.5 | 1.3 | 42 | 10.7 | 54.4 | 8/9/2021 | 9/9/2021 | 4/19/2022 | | | |
| 2003 | 27-Jun-22 | 1 | 26.3 | 1.7 | 35 | 1.2 | 182.7 | 1.5 | 218 | 1.8 | 34.5 | 1.4 | 49.5 | 1.3 | 42 | 30.6 | 200.4 | 2/17/2021 | 4/17/2021 | | 6/16/2021 | | |
| 2004 | 27-Jun-22 | 1 | 26.7 | 1.5 | 30.4 | 1.5 | 12.5 | 1.5 | -1 | 1.4 | 34.5 | 1.4 | 49.5 | 1.3 | 42 | 28.5 | 5.7 | 2/11/2021 | 4/10/2021 | 1/15/2022 | | | |
| 2005 | 27-Jun-22 | 1 | 18.5 | 2.8 | 10.1 | 1.9 | 211.2 | 1.5 | 275.4 | 1.8 | 34.5 | 1.4 | 49.5 | 1.3 | 42 | 14.3 | 243.3 | 9/30/2021 | 11/2/2021 | 4/10/2022 | | | |
| 2006 | 27-Jun-22 | 1 | 24.5 | 2.1 | 26.5 | 1.3 | 183.3 | 2.5 | 182.1 | 1.4 | 34.5 | 1.4 | 49.5 | 1.3 | 42 | 25.5 | 182.7 | 2/13/2021 | 4/11/2021 | 1/20/2022 | 6/26/2021 | 1/27/2022 | |
| 2007 | 27-Jun-22 | 1 | 13.6 | 1.4 | 15.8 | 1.4 | 358.2 | 2.4 | 332.2 | 1.7 | 34.5 | 1.4 | 49.5 | 1.3 | 42 | 14.7 | 345.2 | 2/7/2021 | 4/10/2021 | 1/17/2022 | 7/20/2021 | | |
| 2008 | 27-Jun-22 | 1 | 99 | 2.6 | 92.7 | 1.3 | 49.7 | 3.3 | 49 | 1.8 | 34.5 | 1.4 | 49.5 | 1.3 | 42 | 95.8 | 49.4 | 2/10/2021 | 4/10/2021 | 1/18/2022 | 2/4/2022 | | |
| 2009 | 27-Jun-22 | 1 | 54.6 | 2.3 | 84.5 | 2.6 | 85.7 | 2.4 | 155.4 | 2.3 | 41.6 | 1.8 | 32.2 | 1.7 | 36.9 | 69.6 | 120.5 | 2/11/2021 | 4/10/2021 | 1/18/2022 | 1/28/2021 | 2/10/2022 | |
| 2010 | 27-Jun-22 | 1 | 18.6 | 3.4 | 41 | 1.8 | 102.4 | 2.2 | 116.6 | 2.9 | 41.6 | 1.8 | 32.2 | 1.7 | 36.9 | 29.8 | 109.5 | 2/11/2021 | 4/10/2021 | 1/11/2022 | 1/29/2022 | | |
| 2011 | 27-Jun-22 | 6 | 26.1 | 2.9 | 24.8 | 1.2 | 22.3 | 2 | 17.3 | 1.2 | 34.5 | 1.4 | 49.5 | 1.3 | 42 | 25.4 | 19.8 | 2/14/2021 | 4/4/2021 | 1/17/2022 | 8/5/2021 | | |
| 2012 | 27-Jun-22 | 6 | 190.7 | 3.5 | 182.3 | 1.7 | 94.7 | 1.7 | 85.9 | 1.8 | 34.5 | 1.4 | 49.5 | 1.3 | 42 | 186.5 | 90.3 | 2/11/2021 | 4/17/2021 | 1/16/2022 | 6/23/2021 | | |
| 2013 | 27-Jun-22 | 1 | 142.8 | 1.6 | 120.2 | 1.4 | 47.5 | 1.5 | 44 | 1.3 | 44.2 | 1.9 | 42.1 | 1.5 | 43.1 | 131.5 | 45.7 | 2/11/2021 | 4/11/2021 | 1/22/2022 | | | |
| 2021 | 27-Jun-22 | 6 | 30.9 | 2.6 | 56.9 | 1.5 | -0.4 | 2.1 | 10.1 | 2.8 | 41.6 | 1.8 | 32.2 | 1.7 | 36.9 | 43.9 | 4.8 | 2/9/2021 | 7/8/2021 | 2/15/2022 | 4/1/2021 | | |
| 2022 | 27-Jun-22 | 6 | -20.7 | 2.5 | 11.9 | 1.5 | -19.4 | 1.5 | -2.5 | 2.2 | 41.6 | 1.8 | 32.2 | 1.7 | 36.9 | -4.4 | -10.9 | 2/9/2021 | 4/12/2021 | 1/24/2022 | 8/7/2021 | | |
| 2023 | 27-Jun-22 | 6 | 2.2 | 2 | 14.1 | 3.1 | 35.8 | 2.8 | 55.8 | 2.4 | 41.6 | 1.8 | 32.2 | 1.7 | 36.9 | 8.1 | 45.8 | 2/10/2021 | 4/10/2021 | 1/18/2022 | 2/7/2022 | | |
| 2024 | 27-Jun-22 | 6 | -18.3 | 2.1 | 13.7 | 1.8 | -1.1 | 2.1 | 11.1 | 2.1 | 41.6 | 1.8 | 32.2 | 1.7 | 36.9 | -2.3 | 5 | 2/10/2021 | 4/10/2021 | 1/18/2022 | | | |
| 2025 | 27-Jun-22 | 6 | -0.9 | 2.3 | 34 | 1.9 | 120.8 | 2.9 | 160 | 3.4 | 41.6 | 1.8 | 32.2 | 1.7 | 36.9 | 16.6 | 140.4 | 2/15/2021 | 8/16/2021 | 6/5/2022 | | | |
| 2026 | 27-Jun-22 | 6 | 382.2 | 2.1 | 609.1 | 2.2 | 179.3 | 2.2 | 157.3 | 3.1 | 41.6 | 1.8 | 32.2 | 1.7 | 36.9 | 495.7 | 168.3 | 8/13/2021 | 10/19/2021 | 6/8/2022 | 1/24/2022 | | |
| 2027 | 27-Jun-22 | 6 | -3 | 2.4 | 61.6 | 2.1 | 11.1 | 2.1 | 41 | 2.5 | 41.6 | 1.8 | 32.2 | 1.7 | 36.9 | 29.3 | 26 | 2/15/2021 | 4/22/2021 | 6/5/2022 | | | |
| 2028 | 27-Jun-22 | 6 | 32.4 | 2.3 | 35.2 | 1.9 | 3 | 2.1 | 27.5 | 2.1 | 41.6 | 1.8 | 32.2 | 1.7 | 36.9 | 33.8 | 15.3 | 2/11/2021 | 9/23/2021 | | | | |
| 2029 | 27-Jun-22 | 6 | 31.7 | 2.8 | 30.1 | 2 | 16.9 | 2.3 | 30.7 | 2.4 | 41.6 | 1.8 | 32.2 | 1.7 | 36.9 | 30.9 | 23.8 | 2/11/2021 | 4/10/2021 | 2/13/2022 | | | |
| 2030 | 27-Jun-22 | 6 | -15.2 | 2.7 | 1.7 | 2.7 | 124.2 | 2.4 | 135.8 | 3.4 | 41.6 | 1.8 | 32.2 | 1.7 | 36.9 | -6.8 | 130 | 9/13/2021 | 10/10/2021 | | | | |
| 2031 | 27-Jun-22 | 6 | 181.3 | 2.9 | 224.2 | 2.7 | 56.4 | 3.2 | 78.1 | 2.2 | 41.6 | 1.8 | 32.2 | 1.7 | 36.9 | 202.8 | 67.3 | 2/11/2021 | 4/10/2021 | 1/24/2022 | | | |
| 2032 | 27-Jun-22 | 6 | 0.9 | 2.6 | 46.6 | 1.8 | 135.4 | 2.6 | 184.8 | 2.4 | 41.6 | 1.8 | 32.2 | 1.7 | 36.9 | 23.7 | 160.1 | 2/24/2021 | 4/28/2021 | 3/5/2022 | | | |
| 2033 | 27-Jun-22 | 1 | 48.7 | 1.5 | 62.6 | 1.3 | 392.8 | 2.2 | 361.4 | 2.2 | 44.2 | 1.9 | 42.1 | 1.5 | 43.1 | 55.6 | 377.1 | 2/17/2022 | 3/29/2022 | | | | |
| 2034 | 27-Jun-22 | 1 | 40.5 | 1.7 | 37.6 | 1.1 | 179.9 | 1.2 | 212.4 | 1.4 | 44.2 | 1.9 | 42.1 | 1.5 | 43.1 | 39 | 196.2 | 2/22/2021 | 4/24/2021 | 3/14/2022 | | | |
| 2035 | 27-Jun-22 | 1 | 79.7 | 1.3 | 99.9 | 1.6 | 175.7 | 1.8 | 203.6 | 1.9 | 44.2 | 1.9 | 42.1 | 1.5 | 43.1 | 89.8 | 189.6 | 2/11/2021 | 4/25/2021 | 3/30/2022 | | | |
| 2036 | 27-Jun-22 | 1 | 1303 | 4.9 | 1278.2 | 4.3 | 4.9 | 1.5 | 4 | 1.1 | 44.2 | 1.9 | 42.1 | 1.5 | 43.1 | 1290.6 | 4.5 | 2/16/2021 | 5/11/2021 | 2/14/2022 | | | |
| 2037 | 27-Jun-22 | 1 | 70.5 | 1.2 | 77.6 | 1.2 | 629.3 | 3 | 823.2 | 2.7 | 44.2 | 1.9 | 42.1 | 1.5 | 43.1 | 74 | 726.3 | 2/10/2021 | 8/17/2021 | 3/13/2022 | | | |
| 2039 | 27-Jun-22 | 1 | 16.7 | 1.8 | 10.8 | 1.5 | 12.2 | 1.7 | 4.7 | 1.1 | 44.2 | 1.9 | 42.1 | 1.5 | 43.1 | 13.8 | 8.4 | 2/11/2021 | 8/12/2021 | 3/3/2022 | | | |
| 2040 | 27-Jun-22 | 1 | 289.6 | 2.1 | 304 | 2.2 | 3.2 | 1.3 | -6.8 | 1.4 | 44.2 | 1.9 | 42.1 | 1.5 | 43.1 | 296.8 | -1.8 | 2/11/2021 | 4/10/2021 | 5/30/2022 | | | |
| 2042 | 27-Jun-22 | 1 | 27.7 | 1.8 | 25.1 | 1.3 | 6.5 | 1.6 | 0.8 | 1.8 | 44.2 | 1.9 | 42.1 | 1.5 | 43.1 | 26.4 | 3.6 | 2/11/2021 | 4/10/2021 | 1/28/2022 | | | |
| 2043 | 27-Jun-22 | 1 | 24.3 | 1.2 | 22.1 | 1.4 | 85.3 | 1.6 | 77.7 | 1.2 | 44.2 | 1.9 | 42.1 | 1.5 | 43.1 | 23.2 | 81.5 | 2/11/2021 | 4/12/2021 | 1/27/2022 | | | |
| 2044 | 27-Jun-22 | 1 | 64.8 | 1.6 | 34.3 | 1.5 | 3.7 | 1.6 | 10.4 | 1.4 | 44.2 | 1.9 | 42.1 | 1.5 | 43.1 | 49.6 | 7.1 | 4/12/2021 | 6/27/2021 | | | | |

| | | | | | | | | | | | | | | | | | | | | | |
|---|---|---|---|---|---|---|---|---|---|---|---|---|---|---|---|---|---|---|---|---|---|
| 2045 | 27-Jun-22 | 1 | 11.8 | 1.3 | 15.6 | 1.1 | 29.6 | 1.2 | 53 | 1.5 | 44.2 | 1.9 | 42.1 | 1.5 | 43.1 | 13.7 | 41.3 | 2/10/2021 | 4/12/2021 | 1/27/2022 | |
| 2046 | 27-Jun-22 | 1 | 324.3 | 2.8 | 704.6 | 3.5 | 3.5 | 2.7 | 16.4 | 2.7 | 44.2 | 1.9 | 42.1 | 1.5 | 43.1 | 514.4 | 10 | 2/22/2021 | 4/24/2021 | 3/16/2022 | 1/23/2022 |
| 2047 | 27-Jun-22 | 1 | 150.1 | 2.5 | 155.3 | 2.1 | 1284 | 3.2 | 1407.3 | 4.3 | 44.2 | 1.9 | 42.1 | 1.5 | 43.1 | 152.7 | 1345.7 | 2/10/2021 | 2/19/2021 | 3/16/2021 | 2/10/2022 |
| 2048 | 27-Jun-22 | 1 | 6 | 2.5 | 3 | 3.5 | 31 | 2.4 | 65.7 | 2.6 | 44.2 | 1.9 | 42.1 | 1.5 | 43.1 | 4.5 | 48.3 | 2/10/2021 | 4/10/2021 | 1/27/2022 | |
| 2049 | 27-Jun-22 | 1 | 100.9 | 2.1 | 125.3 | 2.1 | -24.8 | 2.8 | -2.4 | 3.2 | 44.2 | 1.9 | 42.1 | 1.5 | 43.1 | 113.1 | -13.6 | 2/11/2021 | 4/19/2021 | 1/30/2022 | |
| 2050 | 20-Jun-22 | 1 | -4.2 | 2.3 | 13.1 | 2.2 | -27.1 | 2 | -22.8 | 3.5 | 44.2 | 1.9 | 42.1 | 1.5 | 43.1 | 4.4 | -24.9 | 2/11/2021 | 4/12/2021 | 1/27/2022 | |
| 2051 | 27-Jun-22 | 1 | -13.5 | 3.3 | 14.3 | 3.2 | -12.5 | 2.7 | -0.7 | 3.8 | 25.1 | 2 | 32 | 2.1 | 28.6 | 0.4 | -6.6 | 2/16/2021 | 4/19/2021 | | |
| 2053 | 27-Jun-22 | 1 | 7.6 | 2.6 | 5.8 | 2.9 | 54.1 | 2.4 | 66.6 | 2.5 | 25.1 | 2 | 32 | 2.1 | 28.6 | 6.7 | 60.3 | 2/22/2021 | 4/24/2021 | | |
| 2054 | 27-Jun-22 | 6 | 785.7 | 3.3 | 406.6 | 3.3 | 888.4 | 4.5 | 776.8 | 4.4 | 25.1 | 2 | 32 | 2.1 | 28.6 | 596.1 | 832.6 | 2/10/2021 | 10/14/2021 | | |
| 3001 | 20-Jun-22 | 6 | 12.4 | 1.5 | 13.8 | 0.8 | 31.1 | 1 | 46.6 | 1.1 | 72.5 | 1.1 | 71.5 | 1 | 72 | 13.1 | 38.8 | 9/28/2021 | 10/28/2021 | | |
| 3002 | 20-Jun-22 | 6 | 116.6 | 1.3 | 135 | 1.3 | 183.4 | 1.2 | 190.9 | 1.1 | 72.5 | 1.1 | 71.5 | 1 | 72 | 125.8 | 187.2 | 9/28/2021 | | | |
| 3003 | 20-Jun-22 | 6 | 8.9 | 1.1 | 14.6 | 1.1 | 6.3 | 1 | 10.8 | 1 | 72.5 | 1.1 | 71.5 | 1 | 72 | 11.7 | 8.5 | 11/30/2021 | 12/28/2021 | | |
| 3004 | 20-Jun-22 | 6 | 17.2 | 1.2 | 22 | 1 | 110 | 1.1 | 99 | 1 | 72.5 | 1.1 | 71.5 | 1 | 72 | 19.6 | 104.5 | 9/23/2021 | 10/23/2021 | | |
| 3005 | 20-Jun-22 | 6 | 50.7 | 1 | 56.1 | 1.3 | 147.1 | 1.3 | 118.9 | 1 | 72.5 | 1.1 | 71.5 | 1 | 72 | 53.4 | 133 | 9/28/2021 | 10/28/2021 | 6/5/2022 | |
| 3006 | 20-Jun-22 | 6 | 1111.3 | 2.2 | 1131.9 | 2.2 | 24.2 | 1.2 | 26.4 | 1.1 | 72.5 | 1.1 | 71.5 | 1 | 72 | 1121.6 | 25.3 | 11/3/2021 | 12/6/2021 | 6/6/2022 | |
| 3007 | 20-Jun-22 | 6 | 183.4 | 1 | 203 | 1.3 | 11.8 | 1 | 5.8 | 1 | 72.5 | 1.1 | 71.5 | 1 | 72 | 193.2 | 8.8 | 7/17/2021 | 8/19/2021 | 3/2/2022 | |
| 3008 | 20-Jun-22 | 6 | 448.7 | 1.1 | 488.6 | 0.9 | 204.7 | 0.8 | 143.2 | 1 | 72.5 | 1.1 | 71.5 | 1 | 72 | 468.6 | 174 | 10/3/2021 | 11/4/2021 | 5/17/2022 | |
| 3009 | 20-Jun-22 | 6 | 1177.5 | 1.3 | 1312 | 1.6 | 98.1 | 0.8 | 23.6 | 1.1 | 72.5 | 1.1 | 71.5 | 1 | 72 | 1244.8 | 60.9 | 9/19/2021 | 10/23/2021 | 6/6/2022 | |
| 3010 | 20-Jun-22 | 6 | 1720.9 | 2.5 | 1444.9 | 2.5 | 36.1 | 1.1 | 39 | 0.9 | 72.5 | 1.1 | 71.5 | 1 | 72 | 1582.9 | 37.5 | 7/28/2021 | 8/28/2021 | | |
| 3011 | 20-Jun-22 | 6 | 2.8 | 1.2 | -0.7 | 1.2 | 4.7 | 1.1 | 12.5 | 1 | 72.5 | 1.1 | 71.5 | 1 | 72 | 1 | 8.6 | | | | |
| 3012 | 20-Jun-22 | 6 | 1092.8 | 1.3 | 474.6 | 1.2 | 62.8 | 1.5 | 66.1 | 1 | 72.5 | 1.1 | 71.5 | 1 | 72 | 783.7 | 64.4 | 11/10/2021 | 12/8/2021 | 6/6/2022 | |
| 3013 | 20-Jun-22 | 6 | 817 | 1.3 | 2413.8 | 2.2 | 772.6 | 1.6 | 688.3 | 1.5 | 72.5 | 1.1 | 71.5 | 1 | 72 | 1615.4 | 730.5 | 11/10/2021 | 12/8/2021 | 6/6/2022 | |
| 3014 | 20-Jun-22 | 6 | 134.5 | 1 | 169.1 | 1.2 | 79.3 | 1 | 87.1 | 1.1 | 72.5 | 1.1 | 71.5 | 1 | 72 | 151.8 | 83.2 | 2/27/2021 | 3/29/2021 | | |
| 3015 | 20-Jun-22 | 6 | 63.1 | 1 | 43.7 | 1.3 | 135.7 | 1.2 | 100.4 | 1.1 | 72.5 | 1.1 | 71.5 | 1 | 72 | 53.4 | 118 | 4/7/2022 | 6/13/2022 | | |
| 3016 | 20-Jun-22 | 6 | 151.8 | 1 | 150.7 | 1 | 27.8 | 1.2 | 26.5 | 1 | 72.5 | 1.1 | 71.5 | 1 | 72 | 151.2 | 27.2 | 2/27/2022 | 6/7/2022 | | |
| 3017 | 20-Jun-22 | 6 | 125.9 | 1 | 98.5 | 0.8 | 18.4 | 1.2 | 14.2 | 1.3 | 72.5 | 1.1 | 71.5 | 1 | 72 | 112.2 | 16.3 | 2/26/2022 | 3/29/2022 | | |
| 3018 | 20-Jun-22 | 6 | 330.2 | 1.2 | 322 | 1.3 | 22.9 | 0.9 | 20 | 1.1 | 72.5 | 1.1 | 71.5 | 1 | 72 | 326.1 | 21.5 | 2/26/2022 | 3/29/2022 | | |
| 3019 | 20-Jun-22 | 6 | 353.3 | 1.1 | 338.7 | 1 | 284.1 | 1.2 | 314.1 | 1.1 | 72.5 | 1.1 | 71.5 | 1 | 72 | 346 | 299.1 | 2/26/2022 | 3/29/2022 | | |
| 3020 | 20-Jun-22 | 6 | 37.4 | 1 | 50.4 | 1 | 0.7 | 1.2 | 3.4 | 1 | 72.5 | 1.1 | 71.5 | 1 | 72 | 43.9 | 2 | 3/29/2022 | | | |
| 3021 | 20-Jun-22 | 6 | 72.6 | 1.1 | 80.6 | 1.1 | 1.5 | 1 | 2.3 | 1.1 | 72.5 | 1.1 | 71.5 | 1 | 72 | 76.6 | 1.9 | 3/30/2022 | | | |
| 3022 | 20-Jun-22 | 6 | 447.2 | 1.3 | 635.1 | 1.5 | 102.6 | 1.1 | 27.9 | 1.2 | 72.5 | 1.1 | 71.5 | 1 | 72 | 541.1 | 65.2 | 11/9/2021 | 12/9/2021 | 6/5/2022 | |
| 3026 | 20-Jun-22 | 6 | 22.2 | 1.2 | 22.8 | 1 | 215.8 | 1.2 | 158 | 1.2 | 72.5 | 1.1 | 71.5 | 1 | 72 | 22.5 | 186.9 | 2/26/2022 | 3/28/2022 | | |
| 3027 | 20-Jun-22 | 6 | 36.5 | 0.9 | 42.7 | 0.8 | 214 | 1.5 | 225.8 | 1.4 | 72.5 | 1.1 | 71 | 1 | 71.7 | 39.6 | 219.9 | 4/25/2022 | | | |
| 3028 | 20-Jun-22 | 6 | 15.3 | 1.2 | 29.8 | 1.1 | 2.5 | 1.3 | -9 | 1.3 | 72.5 | 1.1 | 71 | 1 | 71.7 | 22.6 | -3.2 | 1/10/2022 | 3/9/2022 | | |
| 3029 | 20-Jun-22 | 6 | 770 | 2 | 197 | 1.4 | 62.2 | 0.9 | 50.1 | 1.1 | 72.5 | 1.1 | 71 | 1 | 71.7 | 483.5 | 56.1 | 10/31/2021 | 12/6/2021 | | |
| 3030 | 20-Jun-22 | 6 | 23.7 | 1.3 | 3 | 1.2 | 34.3 | 1.2 | 31.3 | 1 | 45.5 | 1.3 | 64.8 | 1 | 55.1 | 13.3 | 32.8 | 9/28/2021 | 10/28/2021 | | |
| 3031 | 20-Jun-22 | 6 | 11.3 | 1.2 | 13.8 | 1.1 | 44.6 | 0.7 | 71.8 | 1.3 | 45.5 | 1.3 | 64.8 | 1 | 55.1 | 12.6 | 58.2 | 10/28/2021 | 11/30/2021 | | |
| 3032 | 20-Jun-22 | 6 | 36.9 | 1 | 43.6 | 0.8 | 1662.9 | 1.6 | 1630.3 | 1.7 | 45.5 | 1.3 | 64.8 | 1 | 55.1 | 40.3 | 1646.6 | 12/15/2021 | | | |
| 3033 | 20-Jun-22 | 6 | 90.8 | 1 | 95.2 | 1.1 | 59.1 | 0.9 | 126.3 | 1.1 | 45.5 | 1.3 | 64.8 | 1 | 55.1 | 93 | 92.7 | 12/15/2021 | 3/10/2022 | | |
| 3034 | 20-Jun-22 | 6 | 557 | 1.3 | 468.6 | 1.2 | 1121.2 | 1.4 | 920.1 | 0.9 | 45.5 | 1.3 | 64.8 | 1 | 55.1 | 512.8 | 1020.6 | 10/17/2021 | 11/23/2021 | | |
| 3035 | 20-Jun-22 | 6 | 422 | 1.1 | 433.5 | 1.2 | 382.7 | 1 | 311.5 | 1.3 | 45.5 | 1.3 | 64.8 | 1 | 55.1 | 427.7 | 347.1 | 11/9/2021 | 12/8/2021 | 6/5/2022 | |
| 3036 | 20-Jun-22 | 6 | 79.6 | 1.2 | 82.2 | 1.2 | 397.8 | 1.1 | 433.5 | 1.2 | 45.5 | 1.3 | 64.8 | 1 | 55.1 | 80.9 | 415.7 | 11/9/2021 | 12/8/2021 | 6/5/2022 | |
| 3037 | 20-Jun-22 | 6 | 716.8 | 1.7 | 1283.8 | 2.1 | 37.9 | 0.9 | 47.4 | 1 | 45.5 | 1.3 | 64.8 | 1 | 55.1 | 1000.3 | 42.6 | 9/19/2021 | 10/23/2021 | 6/6/2022 | |
| 3038 | 20-Jun-22 | 6 | 382.3 | 1.4 | 417 | 1.2 | 21.7 | 0.9 | 18.1 | 1.3 | 45.5 | 1.3 | 64.8 | 1 | 55.1 | 399.7 | 19.9 | 11/10/2021 | 12/8/2021 | 6/6/2022 | |
| 3071 | 20-Jun-22 | 7 | 21.9 | 1.3 | 23.9 | 10 | 82.9 | 1.2 | 77 | 1.2 | 25.4 | 1.1 | 29.7 | 1.7 | 27.6 | 22.9 | 79.9 | 1/19/2022 | 3/16/2022 | | |
| 3072 | 20-Jun-22 | 7 | 94.3 | 1.3 | 55.2 | 1.3 | 2.4 | 1.1 | 16.1 | 1.4 | 25.4 | 1.1 | 29.7 | 1.7 | 27.6 | 74.7 | 9.3 | 1/19/2022 | 3/16/2022 | | |
| 3073 | 20-Jun-22 | 7 | 34.8 | 1 | 33.6 | 1 | 2.6 | 1.1 | 8.1 | 0.9 | 25.4 | 1.1 | 29.7 | 1.7 | 27.6 | 34.2 | 5.3 | 4/19/2021 | 6/19/2021 | 3/20/2022 | |
| 3074 | 20-Jun-22 | 7 | 28.2 | 1.2 | 34.5 | 1.5 | 20.8 | 1 | 35 | 1.4 | 25.4 | 1.1 | 29.7 | 1.7 | 27.6 | 31.4 | 27.9 | 1/19/2022 | 3/16/2022 | | |
| 3075 | 20-Jun-22 | 1 | 181.2 | 1 | 145.1 | 1.3 | 263.8 | 1.1 | 240.5 | 1 | 25.4 | 1.1 | 29.7 | 1.7 | 27.6 | 163.2 | 252.1 | 1/19/2022 | 3/16/2022 | | |
| 3076 | 20-Jun-22 | 7 | 1320.5 | 2 | 542 | 1.2 | 21 | 1.1 | 59.5 | 1.2 | 25.4 | 1.1 | 29.7 | 1.7 | 27.6 | 931.3 | 40.2 | 10/28/2021 | 11/25/2021 | 6/9/2022 | |
| 3077 | 20-Jun-22 | 7 | 931.1 | 1.9 | 933.5 | 6.2 | 809.2 | 1.3 | 26.8 | 0 | 25.4 | 1.1 | 29.7 | 1.7 | 27.6 | 932.3 | 418 | 10/28/2021 | 11/25/2021 | 6/9/2022 | |
| 3078 | 20-Jun-22 | 1 | 332.1 | 1.2 | 516.7 | 1.4 | 725.7 | 1.3 | 717.9 | 1.5 | 25.4 | 1.1 | 29.7 | 1.7 | 27.6 | 424.4 | 721.8 | 10/4/2021 | 11/6/2021 | 6/9/2022 | |
| 3079 | 20-Jun-22 | 7 | -9 | 1 | 3.8 | 1.4 | 83.3 | 1.4 | 147.6 | 1.2 | 25.4 | 1.1 | 29.7 | 1.7 | 27.6 | -2.6 | 115.5 | 9/29/2021 | 11/1/2021 | 4/18/2022 | |
| 3080 | 20-Jun-22 | 8 | 164.1 | 1.2 | 74.2 | 1.1 | 966.7 | 1.9 | 990 | 1.6 | 42.7 | 1.1 | 54.9 | 1.9 | 48.8 | 119.2 | 978.3 | 1/18/2022 | 3/18/2022 | | |
| 3081 | 20-Jun-22 | 7 | 46.3 | 1.3 | 93 | 1.4 | 306.5 | 1.1 | 375.1 | 14.8 | 25.4 | 1.1 | 29.7 | 1.7 | 27.6 | 69.7 | 340.8 | 8/8/2021 | 9/8/2021 | 5/9/2022 | |
| 3082 | 20-Jun-22 | 7 | 93 | 1.5 | 148.8 | 1.6 | 10.8 | 1.4 | 7.9 | 1.8 | 46.4 | 2 | 61.4 | 1.2 | 53.9 | 120.9 | 9.3 | 6/10/2021 | 10/29/2021 | | |
| 3084 | 20-Jun-22 | 7 | -1.6 | 0.9 | 20 | 0.8 | 117.3 | 1.1 | 59 | 1.1 | 45.5 | 1.3 | 64.8 | 1 | 55.1 | 9.2 | 88.1 | 2/5/2022 | 3/7/2022 | | |
| 3085 | 20-Jun-22 | 7 | 258.1 | 1 | 318.3 | 1.5 | 103.2 | 1 | 133.6 | 1.1 | 45.5 | 1.3 | 64.8 | 1 | 55.1 | 288.2 | 118.4 | 9/28/2021 | 10/28/2021 | 6/9/2022 | |
| 3086 | 20-Jun-22 | 7 | 304.6 | 1.1 | 556.6 | 1.4 | 708 | 1.4 | 479.2 | 1.1 | 45.5 | 1.3 | 64.8 | 1 | 55.1 | 430.6 | 593.6 | 11/2/2021 | 12/2/2021 | 6/8/2022 | |
| 3087 | 20-Jun-22 | 7 | -3.6 | 1.1 | 24.2 | 1.1 | 154.3 | 1.3 | 291.7 | 1.3 | 46.4 | 2 | 61.4 | 1.2 | 53.9 | 10.3 | 223 | 12/11/2021 | 3/20/2022 | | |
| 3088 | 20-Jun-22 | 7 | 19 | 1.3 | 20.4 | 1.3 | 161 | 1.4 | 475.4 | 1.4 | 46.4 | 2 | 61.4 | 1.2 | 53.9 | 19.7 | 318.2 | 11/2/2021 | 1/13/2022 | 6/9/2022 | |
| 3089 | 20-Jun-22 | 7 | 394.2 | 1.1 | 498.3 | 1.1 | 6.3 | 1 | 12.8 | 1.5 | 46.4 | 2 | 61.4 | 1.2 | 53.9 | 446.2 | 9.5 | 9/28/2021 | 11/28/2021 | 5/19/2022 | |
| 3090 | 20-Jun-22 | 7 | 9.4 | 0.9 | -279 | 1.1 | -10.6 | 1.2 | 1 | 1 | 25.4 | 1.1 | 29.7 | 1.7 | 27.6 | -134.8 | -4.8 | 1/11/2022 | 2/13/2022 | | |
| 3091 | 20-Jun-22 | 7 | 197.7 | 1.1 | 20.4 | 1.5 | 123.1 | 1.1 | 19.4 | 1 | 25.4 | 1.1 | 29.7 | 1.7 | 27.6 | 109 | 71.2 | 2/11/2021 | 5/3/2021 | 3/2/2022 | |
| 3092 | 20-Jun-22 | 7 | 9.9 | 1.3 | 6.1 | 1.6 | 7 | 1.7 | 9.2 | 2.3 | 46.4 | 2 | 61.4 | 1.2 | 53.9 | 8 | 8.1 | 9/28/2021 | | | |
| 3094 | 20-Jun-22 | 7 | 35.2 | 1.3 | 38 | 1.9 | 105.4 | 2.3 | 96.8 | 1.7 | 46.4 | 2 | 61.4 | 1.2 | 53.9 | 36.6 | 101.1 | | | | |
| 3096 | 20-Jun-22 | 1 | 36.4 | 1.3 | 45 | 1 | 224.9 | 1.3 | 198.2 | 1.1 | 50.4 | 1 | 34.8 | 0.9 | 42.6 | 40.7 | 211.6 | 2/20/2022 | | | |
| 3097 | 20-Jun-22 | 1 | 118.6 | 1.3 | 104.1 | 0.8 | 1.3 | 1 | 2.4 | 1 | 50.4 | 1 | 34.8 | 0.9 | 42.6 | 111.4 | 1.9 | 3/1/2021 | 7/18/2021 | 1/8/2022 | |
| 3098 | 20-Jun-22 | 1 | 111.7 | 1.2 | 102.7 | 1.1 | 882.5 | 1.7 | 778.1 | 1.5 | 50.4 | 1 | 34.8 | 0.9 | 42.6 | 107.2 | 830.3 | 7/24/2021 | 8/24/2021 | 3/30/2022 | |
| 3099 | 20-Jun-22 | 1 | 99.1 | 2 | 159.8 | 1 | 592.5 | 1.4 | 693 | 1.1 | 50.4 | 1 | 34.8 | 0.9 | 42.6 | 129.5 | 642.7 | 7/24/2021 | 8/24/2021 | 3/30/2022 | |
| 3100 | 20-Jun-22 | 1 | 0.4 | 1 | 1.4 | 1.3 | 3.1 | 1.1 | 2.5 | 1.1 | 50.4 | 1 | 34.8 | 0.9 | 42.6 | 0.9 | 2.8 | | | | |
| 3101 | 20-Jun-22 | 1 | 360.7 | 2.1 | 60.7 | 1.3 | 202.2 | 2.2 | 149.7 | 1.1 | 50.4 | 1 | 34.8 | 0.9 | 42.6 | 210.7 | 175.9 | 10/26/2021 | 12/1/2021 | 6/7/2022 | |
| 3102 | 20-Jun-22 | 8 | 13.6 | 1.1 | 8.4 | 1.8 | 5.6 | 1.1 | 0.1 | 1.5 | 50.4 | 1 | 34.8 | 0.9 | 42.6 | 11 | 2.8 | 1/15/2022 | 3/15/2022 | | |
| 3104 | 20-Jun-22 | 8 | -6.7 | 1 | 0 | 1.2 | 106.3 | 1 | 71.9 | 1 | 66.4 | 1.2 | 67.5 | 1.8 | 67 | -3.3 | 89.1 | 8/19/2021 | 9/20/2021 | 3/28/2022 | |
| 3105 | 20-Jun-22 | 8 | 64 | 1.3 | 68.3 | 1.1 | 329.3 | 0.9 | 428.5 | 1.1 | 66.4 | 1.2 | 67.5 | 1.8 | 67 | 66.2 | 378.9 | 8/8/2021 | 9/8/2021 | 3/30/2022 | |
| 3106 | 20-Jun-22 | 8 | -98 | 1 | 41.4 | 1.7 | 22.7 | 1.1 | 31.4 | 1.3 | 66.4 | 1.2 | 67.5 | 1.8 | 67 | -28.3 | 27.1 | 1/19/2022 | 3/19/2022 | | |
| 3107 | 20-Jun-22 | 8 | 11.6 | 1.3 | 13.9 | 3 | 1 | 1.5 | 7.7 | 1.9 | 46.4 | 2 | 61.4 | 1.2 | 53.9 | 12.8 | 4.4 | 9/23/2021 | 3/3/2022 | | |
| 3108 | 20-Jun-22 | 8 | 957 | 5.8 | 681.8 | 2.2 | 27.5 | 2 | 28.9 | 1.6 | 46.4 | 2 | 61.4 | 1.2 | 53.9 | 819.4 | 28.2 | 9/23/2021 | 10/23/2021 | 6/12/2022 | |
| 3109 | 20-Jun-22 | 8 | 15.1 | 1.9 | 12.6 | 1.5 | 329.2 | 2.2 | 315.9 | 1.9 | 46.4 | 2 | 61.4 | 1.2 | 53.9 | 13.9 | 322.5 | 2/20/2022 | 4/4/2022 | | |
| 3110 | 20-Jun-22 | 8 | 103.6 | 1.3 | 41.8 | 1.2 | 350 | 1.4 | 333.9 | 1 | 66.4 | 1.2 | 67.5 | 1.8 | 67 | 72.7 | 341.9 | 1/17/2022 | 3/16/2022 | | |
| 3111 | 20-Jun-22 | 8 | 106.4 | 1.4 | 63.8 | 0.9 | 42.9 | 1.5 | 3.6 | 1.1 | 66.4 | 1.2 | 67.5 | 1.8 | 67 | 85.1 | 23.2 | 4/18/2021 | 8/9/2021 | 3/20/2022 | |
| 3112 | 20-Jun-22 | 8 | 78.2 | 1.2 | 97.7 | 1.5 | 29.9 | 1.1 | 50.6 | 1.1 | 66.4 | 1.2 | 67.5 | 1.8 | 67 | 88 | 40.3 | 7/28/2021 | 8/26/2021 | 3/29/2022 | |
| 3113 | 20-Jun-22 | 8 | -11 | 1.5 | 11.2 | 1.2 | 2.2 | 1.2 | 8 | 1.8 | 66.4 | 1.2 | 67.5 | 1.8 | 67 | 0.1 | 5.1 | 1/19/2022 | 3/16/2022 | | |
| 3114 | 20-Jun-22 | 8 | 32.3 | 1.2 | 46.2 | 1.4 | 76.8 | 1.1 | 64.4 | 1.6 | 66.4 | 1.2 | 67.5 | 1.8 | 67 | 39.2 | 70.6 | 8/7/2021 | 9/16/2021 | 5/7/2022 | |

| | | | | | | | | | | | | | | | | | | | | | |
|---|---|---|---|---|---|---|---|---|---|---|---|---|---|---|---|---|---|---|---|---|---|
| 3115 | 20-Jun-22 | 8 | 272.9 | 1 | 231.6 | 1.6 | -3.5 | 1.1 | 8 | 1.9 | 66.4 | 1.2 | 67.5 | 1.8 | 67 | 252.2 | 2.2 | 12/7/2021 | 2/7/2022 | 6/10/2022 | |
| 3116 | 20-Jun-22 | 8 | 97.2 | 1.1 | 65.9 | 1.8 | -13.1 | 1.4 | -6.9 | 1.8 | 66.4 | 1 | 67.5 | 1.8 | 67 | 81.6 | -10 | 4/19/2021 | 8/9/2021 | 2/19/2022 | |
| 3117 | 20-Jun-22 | 8 | 105.1 | 0.9 | 113.5 | 1.4 | 32.9 | 1.2 | 39.6 | 1.8 | 66.4 | 1.2 | 67.5 | 1.2 | 67 | 109.3 | 36.2 | 2/9/2021 | 4/11/2021 | 1/15/2022 | |
| 3118 | 20-Jun-22 | 8 | -1.9 | 1.4 | 2.5 | 1.1 | 198.3 | 1.3 | 158.1 | 1.1 | 66.4 | 1.2 | 67.5 | 1.8 | 67 | 0.3 | 178.2 | 9/28/2021 | 10/28/2021 | 3/29/2022 | |
| 3119 | 20-Jun-22 | 8 | 36.7 | 1 | 79.1 | 1.4 | 549.9 | 1.3 | 376 | 1.2 | 42.7 | 1.1 | 54.9 | 1.9 | 48.8 | 57.9 | 462.9 | | | | |
| 3120 | 20-Jun-22 | 8 | 2.3 | 0.9 | 41.2 | 1.1 | 138.1 | 1.3 | 215.2 | 1 | 42.7 | 1.1 | 54.9 | 1.9 | 48.8 | 21.8 | 176.6 | 2/26/2022 | 3/28/2022 | | |
| 3121 | 20-Jun-22 | 8 | 175.8 | 1.8 | 95.2 | 1 | 51 | 1 | 60.4 | 1.7 | 42.7 | 1.1 | 54.9 | 1.9 | 48.8 | 135.5 | 55.7 | 10/5/2021 | 11/18/2021 | 3/28/2022 | |
| 3122 | 20-Jun-22 | 8 | 0.6 | 1.1 | 3.6 | 1.1 | 56.1 | 1 | 89.8 | 1.8 | 42.7 | 1.1 | 54.9 | 1.9 | 48.8 | 2.1 | 73 | 12/1/2021 | 3/6/2022 | | |
| 3123 | 20-Jun-22 | 8 | 69.6 | 1.3 | 79.3 | 0.9 | -0.4 | 1.2 | 7 | 1.1 | 42.7 | 1.1 | 54.9 | 1.9 | 48.8 | 74.5 | 3.3 | 1/23/2022 | 3/30/2022 | | |
| 3124 | 20-Jun-22 | 8 | 30 | 1.2 | 46.3 | 1.2 | 172.1 | 0.9 | 136.3 | 1.1 | 42.7 | 1.1 | 54.9 | 1.9 | 48.8 | 38.1 | 154.2 | 8/23/2021 | 10/4/2021 | | |
| 3125 | 20-Jun-22 | 8 | 560.5 | 1.4 | 605.6 | 10.9 | -18.1 | 1.4 | -19.6 | 1.1 | 42.7 | 1.1 | 54.9 | 1.9 | 48.8 | 583 | -18.8 | 12/1/2021 | 2/2/2022 | 6/9/2022 | |
| 3126 | 20-Jun-22 | 8 | 108.2 | 1.3 | 129.8 | 1.3 | 372.6 | 1.2 | 533.6 | 1.5 | 42.7 | 1.1 | 54.9 | 1.9 | 48.8 | 119 | 453.1 | 1/27/2022 | 3/24/2022 | | 7/4/2021 |
| 3127 | 20-Jun-22 | 8 | 1.5 | 1.4 | 17 | 1.3 | 105.3 | 0.9 | 361.9 | 2.5 | 42.7 | 1.1 | 54.9 | 1.9 | 48.8 | 9.2 | 233.6 | 2/14/2021 | 4/16/2021 | | |
| 3128 | 20-Jun-22 | 8 | 63.2 | 1.2 | 34.9 | 1.3 | 17.9 | 1.5 | 21.5 | 1.6 | 42.7 | 1.1 | 54.9 | 1.9 | 48.8 | 49.1 | 19.7 | 1/15/2022 | 3/14/2022 | | |
| 3129 | 20-Jun-22 | 8 | 66.9 | 0.9 | 122.5 | 1.4 | 234.8 | 1.2 | 255.9 | 1.2 | 42.7 | 1.1 | 54.9 | 1.9 | 48.8 | 94.7 | 245.3 | 7/13/2021 | 8/14/2021 | 2/22/2022 | |
| 3130 | 20-Jun-22 | 1 | 8.9 | 1 | 12.3 | 0.9 | 9.5 | 1.2 | 3.8 | 0.9 | 50.4 | 1 | 34.8 | 0.9 | 42.6 | 10.6 | 6.6 | 7/27/2021 | 8/25/2021 | 3/5/2022 | |
| 3131 | 20-Jun-22 | 1 | 361 | 1.3 | 424.6 | 1.8 | 246.2 | 1 | 280.1 | 1.3 | 50.4 | 1 | 34.8 | 0.9 | 42.6 | 392.8 | 263.2 | 10/24/2021 | 11/25/2021 | 6/2/2022 | |
| 3132 | 20-Jun-22 | 1 | 334.5 | 1.1 | 837.2 | 2.2 | 205.5 | 1.4 | 188.9 | 1.2 | 50.4 | 1 | 34.8 | 0.9 | 42.6 | 585.8 | 197.2 | 10/10/2021 | 11/10/2021 | 6/2/2022 | |
| 3133 | 20-Jun-22 | 1 | 102.9 | 2.5 | 97.7 | 1.2 | 51 | 1.1 | 102 | 1.1 | 50.4 | 1 | 34.8 | 0.9 | 42.6 | 100.3 | 76.5 | 2/20/2021 | 8/16/2021 | 2/20/2022 | |
| 3134 | 20-Jun-22 | 1 | 69.8 | 1 | 46.8 | 1.4 | 21.8 | 1.5 | 19.5 | 1.2 | 50.4 | 1 | 34.8 | 0.9 | 42.6 | 58.3 | 20.6 | 11/28/2021 | 3/22/2022 | | |
| 3135 | 20-Jun-22 | 1 | 13.6 | 1.4 | 15.2 | 1.1 | 228.8 | 2.4 | 240.7 | 1.1 | 50.4 | 1 | 34.8 | 0.9 | 42.6 | 14.4 | 234.8 | 2/26/2022 | 3/28/2022 | | |
| 3136 | 20-Jun-22 | 8 | 8.9 | 0.9 | 3.2 | 1.2 | 395.2 | 1.4 | 480.4 | 1.4 | 46.4 | 2 | 61.4 | 1.2 | 53.9 | 6 | 437.8 | 10/24/2021 | 11/24/2021 | | |
| 3137 | 20-Jun-22 | 8 | 12.1 | 0.9 | 14.7 | 0.9 | 88 | 1.1 | 55.6 | 1.4 | 46.4 | 2 | 61.4 | 1.2 | 53.9 | 13.4 | 71.8 | 1/23/2022 | | | |
| 3138 | 20-Jun-22 | 8 | -1 | 1.1 | -16.2 | 1.1 | 2.1 | 1 | -0.9 | 1.2 | 46.4 | 2 | 61.4 | 1.2 | 53.9 | -8.6 | 0.6 | 10/24/2021 | 11/24/2021 | | |
| 3139 | 20-Jun-22 | 8 | 9.4 | 1 | 8.4 | 1 | 86.4 | 1.2 | 108.6 | 1.2 | 46.4 | 2 | 61.4 | 1.2 | 53.9 | 8.9 | 97.5 | 1/23/2022 | 3/23/2022 | | |
| 3140 | 20-Jun-22 | 8 | 140.1 | 0.9 | 160.2 | 1.2 | 3.6 | 1.2 | 11.3 | 1 | 50.4 | 1 | 34.8 | 0.9 | 42.6 | 150.2 | 7.4 | 1/23/2022 | 3/20/2022 | | |
| 3141 | 20-Jun-22 | 6 | 29.5 | 1.1 | 34.1 | 1 | 21.6 | 1 | 26.4 | 0.9 | 72.5 | 1.1 | 71.5 | 1 | 72 | 31.8 | 24 | 8/3/2021 | 9/12/2021 | 3/13/2022 | |
| 3142 | 20-Jun-22 | 8 | 11.9 | 0.9 | -4.4 | 2 | 18.4 | 1 | 10.4 | 1.3 | 46.4 | 2 | 61.4 | 1.2 | 53.9 | 3.7 | 14.4 | 2/26/2022 | 3/28/2022 | | |
| 3143 | 20-Jun-22 | 8 | 2 | 1.1 | 8.8 | 1.3 | 118.5 | 1.2 | 174.8 | 1.3 | 46.4 | 2 | 61.4 | 2 | 53.9 | 5.4 | 146.6 | 2/2/2022 | 3/29/2022 | | |
| 3144 | 20-Jun-22 | 8 | -9.7 | 0.8 | -19.9 | 1 | -9.7 | 0.8 | 43.1 | 1 | 46.4 | 2 | 61.4 | 2 | 53.9 | -14.8 | 16.7 | 2/26/2022 | 3/28/2022 | | |
| 3145 | 20-Jun-22 | 8 | -13.1 | 1.1 | 99.1 | 1.4 | 9.3 | 1.1 | -18.5 | 1 | 46.4 | 2 | 61.4 | 2 | 53.9 | 43 | -4.6 | 9/28/2021 | 11/28/2021 | | |
| 3146 | 20-Jun-22 | 8 | -6 | 1.2 | 79.2 | 1.5 | 0.8 | 1.2 | 4.8 | 1.3 | 46.4 | 2 | 61.4 | 2 | 53.9 | 36.6 | 2.8 | 8/25/2021 | 9/25/2021 | 3/22/2022 | |
| 3147 | 20-Jun-22 | 8 | 104.7 | 1 | 35.3 | 1 | 359.3 | 1.3 | 1.6 | 1.2 | 46.4 | 2 | 61.4 | 2 | 53.9 | 70 | 180.5 | 11/28/2021 | 2/23/2022 | | |
| 3148 | 20-Jun-22 | 8 | 19.3 | 1 | -23.2 | 1.6 | 447.7 | 1.2 | 12.1 | 2.7 | 46.4 | 2 | 61.4 | 2 | 53.9 | -2 | 229.9 | 1/19/2022 | 3/16/2022 | | |
| 3149 | 20-Jun-22 | 8 | -3.1 | 1.1 | 10.5 | 1.3 | -13 | 1.6 | -8.6 | 1.2 | 46.4 | 2 | 61.4 | 2 | 53.9 | 3.7 | -10.8 | 11/28/2021 | 2/23/2022 | | |
| 3150 | 20-Jun-22 | 8 | 24.6 | 0.9 | 3.4 | 1.2 | 250.4 | 1 | 65.2 | 1.4 | 46.4 | 2 | 61.4 | 2 | 53.9 | 14 | 157.8 | 1/23/2022 | 2/20/2022 | | |
| 3151 | 20-Jun-22 | 8 | 9.4 | 1.2 | 7.6 | 1.2 | 44.6 | 1 | 21.4 | 1.1 | 46.4 | 2 | 61.4 | 2 | 53.9 | 8.5 | 33 | 9/28/2021 | 10/28/2021 | | |
