## Supplementary info for "A handheld luminometer with sub-attomole limit of detection for distributed applications in global health"

#### Supplementary Design Details

A cloud-based mechanical CAD model of the design is provided here:
https://tinyurl.com/3sh6myts

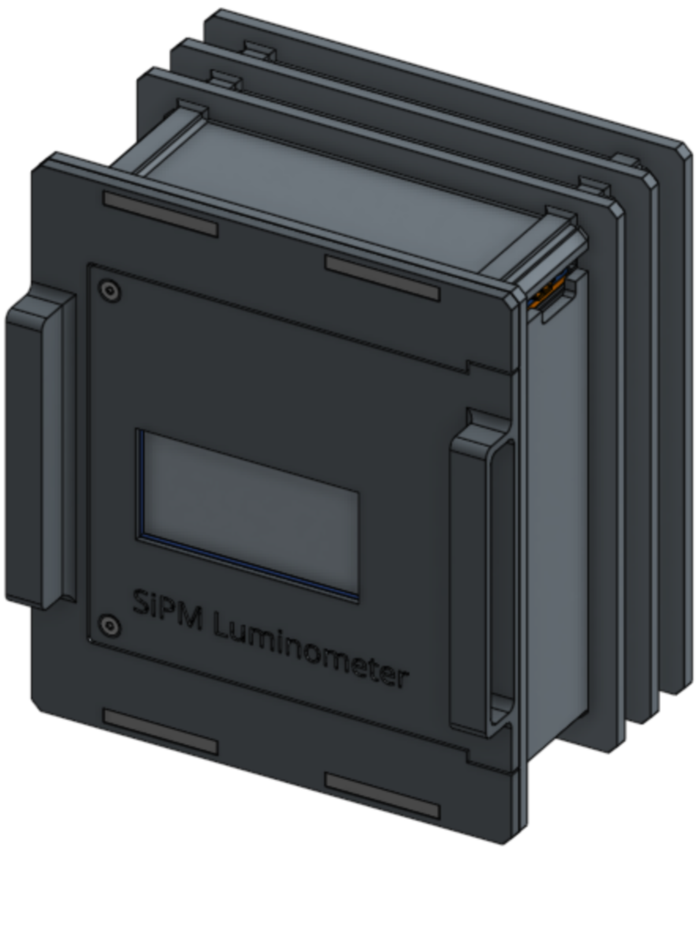

### Supplementary Datasets

Spreadsheets are provided for each dataset below.

#### Supplementary data 1: Firefly luciferase titration

Raw data is provided for the firefly luciferase titrations in figure 2a. Separate tabs denote data from the Bright-Glo assay condition and the Luciferase Assay System (LAS).

#### Supplementary data 2: Fleet normalization with nanoluciferase in whole blood

The full set of luminometers was used to perform a simultaneous titration of nanoluciferase in order to normalize the sensitivity of all devices. A background of whole blood was used to simulate the spLUC assay condition.

#### Supplementary data 3: Bangladesh serology study data

All raw data from the Mirzapur serology study are provided, along with participant self-reported data.

#### Supplementary data 4: Bill of Materials

A full bill of materials is provided for the luminometer. The table is divided into three sections:

1. Assembly items
2. Digital PCB - Itemized
3. Analog PCR - Itemized

Note that in the “Assembly items” table, there are columns for ‘Internal part number’ and also for ‘Vendor part number’. When there is only a vendor part number, this is a commercial item that was used as-is in the assembly. When there is an internal part number, the item was fabricated in-house, and the corresponding vendor part number is typically the material used to fabricate the part. Note that we did not attempt an accurate cost estimate for in-house 3D-printed and laser-cut parts. Typically, these parts were very small and had minimal material cost, usually less than $1.

### Supplementary Movies

#### Supplementary movie 1:

Computer animation of the shutter motion, shown with all components hidden except for the shutter system, tube holder, and sample tubes. Note that in reality the actuation motion occurs very rapidly (motion is complete after approximately 30 ms), while the stationary phase for open and closed positions is one second in duration. The animation only demonstrates the motion profile.

#### Supplementary movie 2:

Computer animation of the shutter motion, with only the shutter system and electronics displayed.

### Supplementary Notes

#### Supplementary Note 1: *Etendue* and optical collection efficiency

The principle of *Etendue* states that the product of surface area and solid angle of a projected emitting surface does not decrease via transmission through any series of optical elements [1]:

$$dU = n^2 dA\, cos(\theta)\, d\Omega$$

Where *dU* is the differential unit of *Etendue*, *n* is the refractive index of the immersed medium, *dA* is the differential unit of surface area, θ is the projected angle, and $d\Omega$ is the differential unit of solid angle. The implications place practical limits on how much light can be collected by an optical sensor with finite surface area and collection angle, given the source's optical emission characteristics. If the source is larger than the sensor, then a de-magnified image could be created at the sensor to match its surface area. However, the conservation of Etendue states that to reduce the area of the image, the acceptance angle at the sensor must be proportionally larger than the collection angle at the source.

Equivalently, the radiance of the original emission source (photon flux per unit volume per unit solid angle) cannot be increased by the use of lenses and/or mirrors. Collection optics therefore cannot play a beneficial role in focusing luminescence emission onto a small detector, beyond the result of positioning the sensor immediately adjacent to the sample. However, a reflecting surface can be used to redirect light emitted in the opposite direction from the sensor, which would otherwise be lost.

By contrast, in fluorescence microscopy, since dye molecules generate photons from a nanoscopic volume, optics with large magnification factors can be used to collect and focus emission onto microscopic detectors with extremely low dark current -- for example, a small cluster of cooled EMCCD or sCMOS camera pixels. Since the dark current of such cooled scientific camera pixels is typically less than one electron per pixel per second [2], the detector noise is insignificant in comparison to photon shot noise from the signal itself. Since the emission volume of a typical luminescence sample is tens to hundreds of µL (1µL = 1 $mm^3$) in volume, similar use of magnification would require impractically large detectors with proportionally large dark currents. The challenges of luminescence detection are therefore distinct from other low-light applications such as single molecule fluorescence microscopy.

#### Supplementary note 2: Optimal sensor size

The optimal sensor size is influenced by the Lambertian emission profile of the sample, the solid angle subtended by the sensor with respect to the sample, and the dark current's dependence on sensor area. Under the assumption that dark current is proportional to area, we extend equation 1 from the main text to include an integrated Lambertian collection profile (uniform emission per unit solid angle, projected onto the plane of the sensor) of the form $(1 - cos(\theta))$, where θ is the half-angle subtended by the sensor. This equation approximates SNR (under the assumption that dark current is much larger than signal current), up to a proportionality constant:

$$SNR \sim \frac{1 - cos(\theta)}{\sqrt{2y^2}}$$

Where y is the half-width of the sensor, and $r_o$ is the distance between the sample and the sensor. Here, we simplify the calculation by temporarily considering the emitter to be a point source (a proper integral over the sample will have the effect of broadening the peak). By substituting $cos(\theta) = r/h$, where $h = \sqrt{(y^2 + r^2)}$ is the hypotenuse of the solid angle, we get:

$$SNR \sim \frac{1}{y} - \frac{r}{y\sqrt{(y^2 + r^2)}}$$

Substituting $\eta = \frac{y}{r}$, we obtain the unitless representation proportional to the SNR:

$$SNR \sim \left( \frac{1}{\eta} - \frac{1}{\eta\sqrt{\eta^2 + 1}} \right)$$

The derivative of the function with respect to y can be set to zero to solve for the optimal value of y, or solved graphically. A result emerges, suggesting that optimal SNR is achieved by using a sensor marginally larger than the sample size. Solved graphically, the optimal sensor-to-sample size ratio is approximately 1.2.

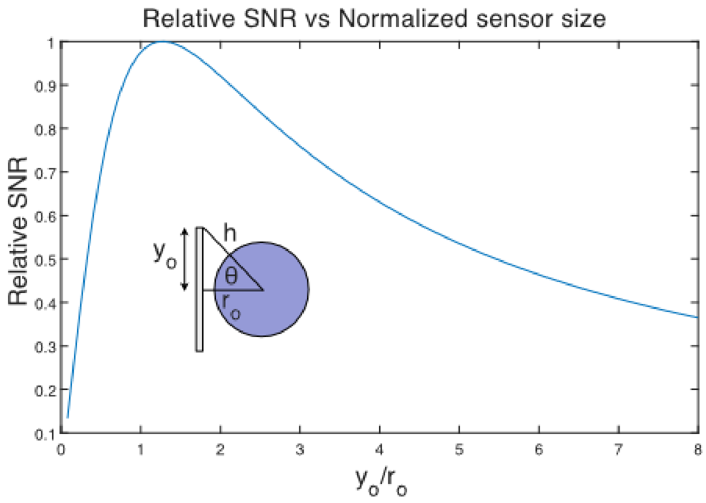

### Supplementary Tables

#### Supplementary table 1: Summary of commercial and academic luminometer Limits of Detection (LODs)

LODs are quoted either in terms of moles of luciferase, moles of ATP, radiometric power, or other units, with all conversions dependent on several assumptions. Conversion between the two luciferase-based systems of reporting depends on the enzyme used (nanoluciferase vs firefly luciferase) and the assay's kinetic conditions. Assuming flash-mode kinetics, the catalytic rate $k_{cat}$ of firefly luciferase was previously reported to be approximately 1.6 $s^{-1}$ [3]. Note also that conversion between enzymatic standards and radiometric power depends on both assay conditions as well as collection efficiency and detector size. Smaller detectors can exhibit superior radiometric LODs due to their lower dark current, while light collection efficiency and LOD for enzymatic signals will be reduced, as detailed in Supplementary Note 2. We estimate that 1E-17 moles of ATP is approximately equivalent to 1.2E-19 moles of firefly luciferase, with each assay operating in flash mode kinetics and the other component in saturation, and with a signal half-life of 30 seconds, although differing assumptions will produce differing results.

All the above factors must be considered when comparing across methods of reporting, none of which capture every aspect relevant to all applications.

| **Make/ref #** | **Model/Author** | **Sensor** | **Portable** | **Battery** | **Limit of detection** | **Cost** |
|---|---|---|---|---|---|---|
| Promega | GloMax | PMT | No | No | 3 E-21 moles of luciferase | $16,313 |
| Berthold | Junior | PMT | Semi[a] | Yes | 5 E-17 moles of ATP | $8,000 |
| BMG | Omega | PMT | No | No | 2 E-17 moles of ATP | $13,560 |
| Tecan | Infinite M-Plex | PMT | No | No | 1.2 E-17 moles of ATP | $36,000 |
| 3M | LX25 | Not reported | Yes | Yes | 5 E-16 moles of ATP | $3,409 |
| [4] | Bunce et al. | Film | No | No | 5E-13 moles of luminol | Not reported |
| [5] | Porakishvili et al. | Cooled CCD | No | No | Conversion not known | £3,000 |
| [6] | Bunce et al. | PIN Photodiode | Yes | Yes | 5E-13 moles of ATP | Not reported |
| [7] | Roda et al. | Smartphone | Yes | Yes | 1E-4 moles/L lactate | Cost of phone |
| [8] | Kim et al. | Smartphone | Yes | Yes | 1-10 pW | Cost of phone |
| [9] | Li et al | SiPM (1 $mm^2$) | Not reported | No | 0.6 fW [b,c,d] | Not reported |

| | | | | | | |
|---|---|---|---|---|---|---|
| [10] | Jung et al. | SiPM (9 $mm^2$) | Yes | Yes | 100 fW[b,c] | Not reported |
| [11] | Calabretta et al. | SiPM (1.7 $mm^2$) | Yes | No | 9E-15 moles of luciferase[b] | Not reported |
| [12] | Baszczyk et al. | SiPM (1 $mm^2$) | Yes | No | 73 fW [b,d] | Not reported |
| **This work** | **CZ Biohub SF** | **SiPM (36 $mm^2$)** | **Yes[e]** | **Yes** | **1.6E-19 moles of luciferase or 1 fW** | **< $1,000** |

[a] Mass is 2 kg and a suitcase-sized carrying case is required for transport.
[b] Sensor active area is small, limiting applications in luminescence detection.
[c] Active cooling used
[d] Computed from quoted limit of detection in counts per second, using a wavelength of 460 nm.
[e] Mass is 515 g, and outer dimensions are 108 x 122 x 80 mm (w x h x d)

#### Supplementary table 2: Data from UCSF volunteer study

| Participant | Log Spluc RLU | LFA | ELISA |
|---|---|---|---|
| 1 | 2.200134 | 0.377057 | 0.5309 |
| 2 | 3.49643 | 4.37561 | 0.7924 |
| 3 | 2.954085 | 3.278838 | 0.53265 |
| 4 | 3.113676 | 3.28483 | 0.575241 |
| 5 | 2.953201 | 3.513158 | 0.8648 |
| 6 | 3.009275 | 4.721729 | 0.72615 |
| 7 | 2.614875 | 1.563393 | 0.381143 |
| 8 | 2.95082 | 3.803675 | 0.8206 |
| 9 | 3.2499 | 6.565445 | 0.9295 |
| 10 | 1.924395 | 0.931394 | 0.22045 |
| 11 | 2.684152 | 2.76165 | 0.66555 |
| 12 | 2.963569 | 1.818232 | 0.705 |
| 13 | 1.513414 | 0.015569 | 0.0069 |
| 14 | 3.165569 | 6.498282 | 0.9146 |
| 15 | 3.243311 | 3.609574 | 1.0823 |
| 16 | 3.440545 | 5.960265 | 1.0014 |
| 17 | 2.199416 | 0.923422 | 0.27795 |
| 18 | 3.581251 | 7.708171 | 0.9794 |
| 19 | 2.955012 | 2.063568 | 0.8963 |
| 20 | 2.908417 | 2.52082 | 0.7193 |
| 21 | 2.617891 | 2.78174 | 0.3493 |
| 22 | 1.31212 | 0.156827 | 0.01195 |
| 23 | 3.697681 | 4.984286 | 1.17285 |
| 24 | 2.668437 | 2.21548 | 0.51135 |
| 25 | 2.827742 | 2.978032 | 0.79475 |

| | | | |
|---|---|---|---|
| 26 | 2.864987 | 3.394816 | * |

*Insufficient blood volume from participant

### Supplementary Figures

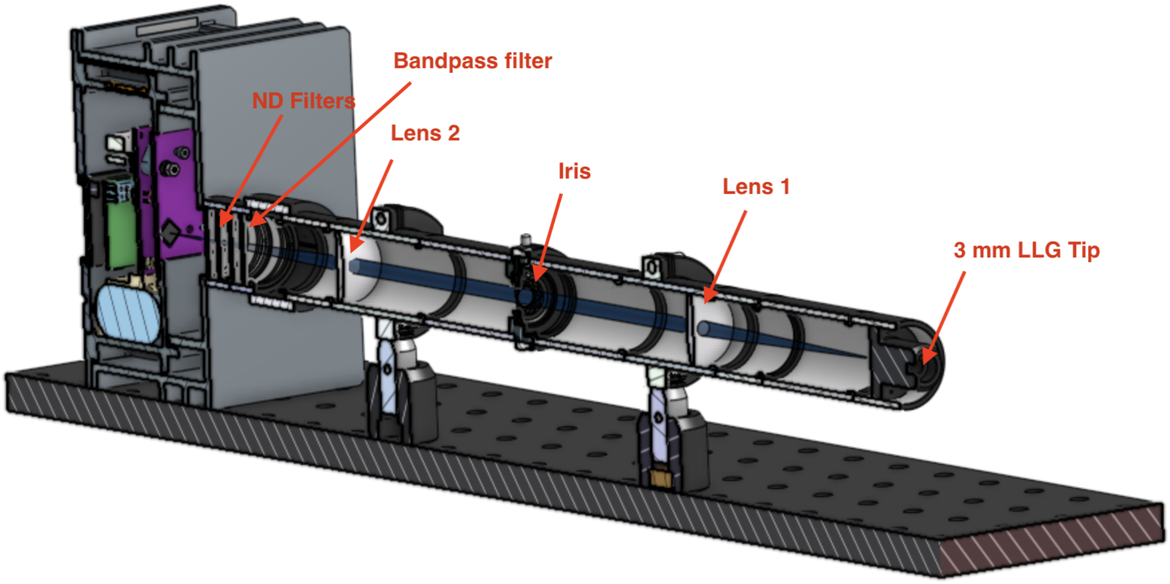


**Supplementary Figure 1: Cutaway view of the radiometric test station**
A 3D CAD model of the radiometric test setup is shown as a cross-section view. The optical path consisted of a 4f relay, imaging the tip of a 3 mm Liquid Light Guide (LLG) onto the luminometer sensor at 1× magnification. Variable numbers of neutral density filters were added or removed as required to vary the optical power, along with the lamp's percentage output setting (0-100%). The lens relay was constructed using two 100 mm focusing lenses (Thorlabs LA1509-A), and an adjustable iris (fixed at 2 mm) to restrict the NA of the optical path. A 25 nm FWHM spectral bandpass filter (Semrock FF02-475/50-25) was employed to approximately match the wavelength of the lamp's spectrum to that of nanoluciferase emission (460 nm). The incident optical power at the luminometer sensor was varied between 0.1 fW to nearly 1E5 fW.

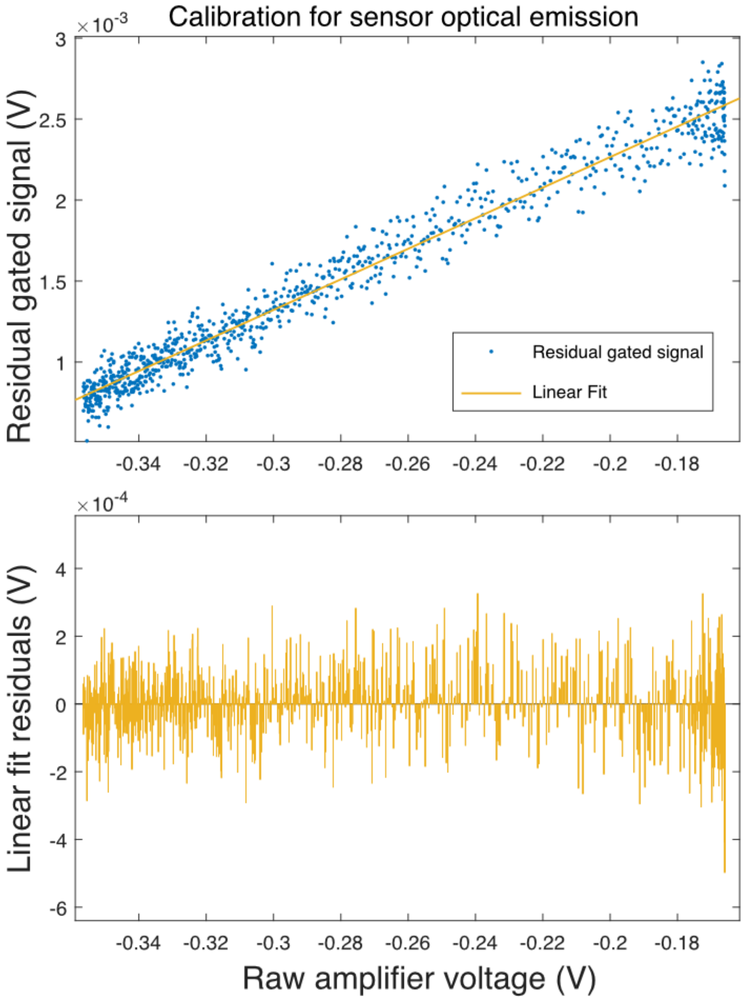


**Supplementary Figure 2: Sensor externally-coupled crosstalk (ECC) calibration.** Top: Scatter plot of residual gated signal, collected during an ambient temperature ramp from 40°C to 4°C, under dark conditions. The residual signal due to the ECC effect is observed to be directly proportional to the raw amplifier voltage, which is in turn linearly proportional to sensor's dark current. The shutter flag modulates the ECC coupling ratio from the sensor back to itself, by blocking and unblocking the reflective sample cavity. The magnitude of the effect represents approximately 150 RLU of correlated error over the recorded temperature range. **Bottom:** Residuals from the linear fit to the data, showing no evidence of bias across the range of sampled temperatures.

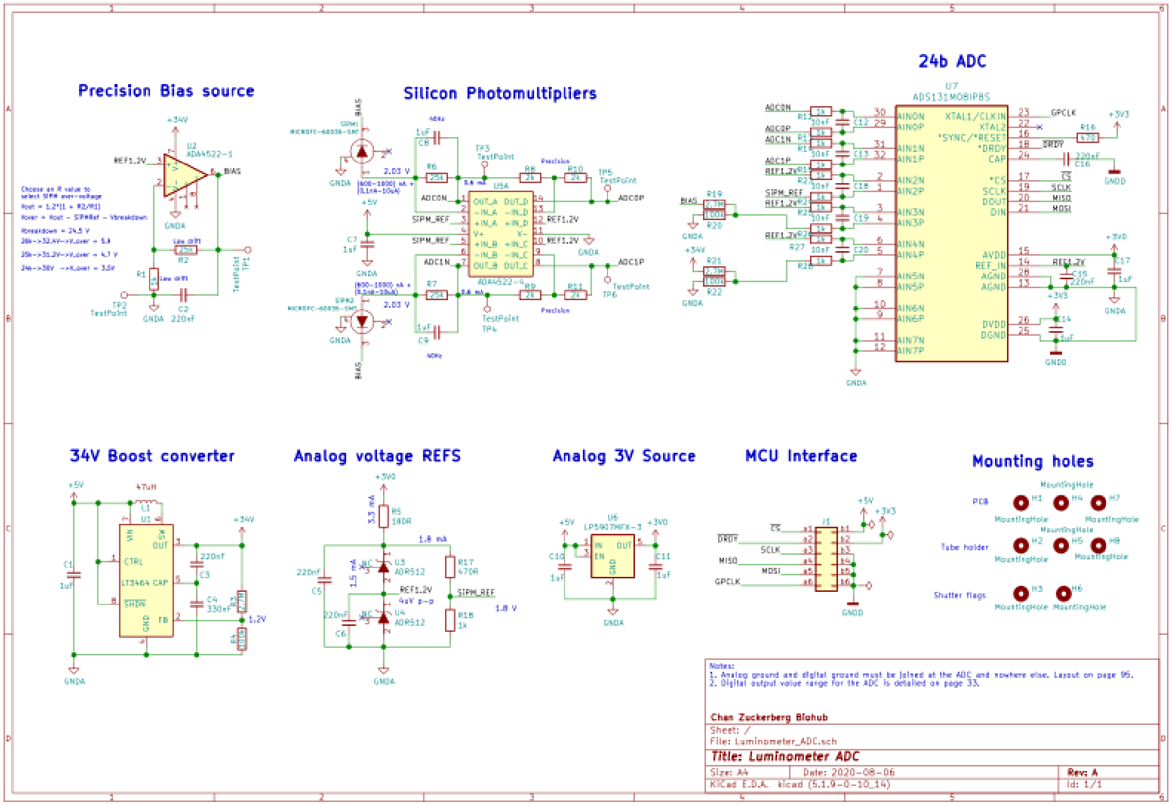


**Supplementary figure 3: Schematic diagram of the analog sensor PCB**. Clockwise, from upper left: the precision bias source uses a zero-drift amplifier and precision low-drift resistors to set the SiPM bias voltage, which controls gain and photon detection efficiency of the sensors. This bias source is powered by an on-board boost converter to generate a 34V supply. The SiPM sensors themselves are read out by fully-differential, zero-drift transimpedance amplifiers, and routed with short traces to a 24-bit differential-input ADC with “global chop” DC bias elimination and digital over-sampling for anti-aliasing. The transimpedance amplifier gain was chosen to position the maximum expected signals near the point of saturation of the amplifier, with parallel capacitors low-pass filtering the bandwidth of the amplified signal. Precision voltage reference sources were used for both the ADC and the differential amplifier. The ADC digitized the signals using an SPI protocol routed directly to a board-to-board interconnect.

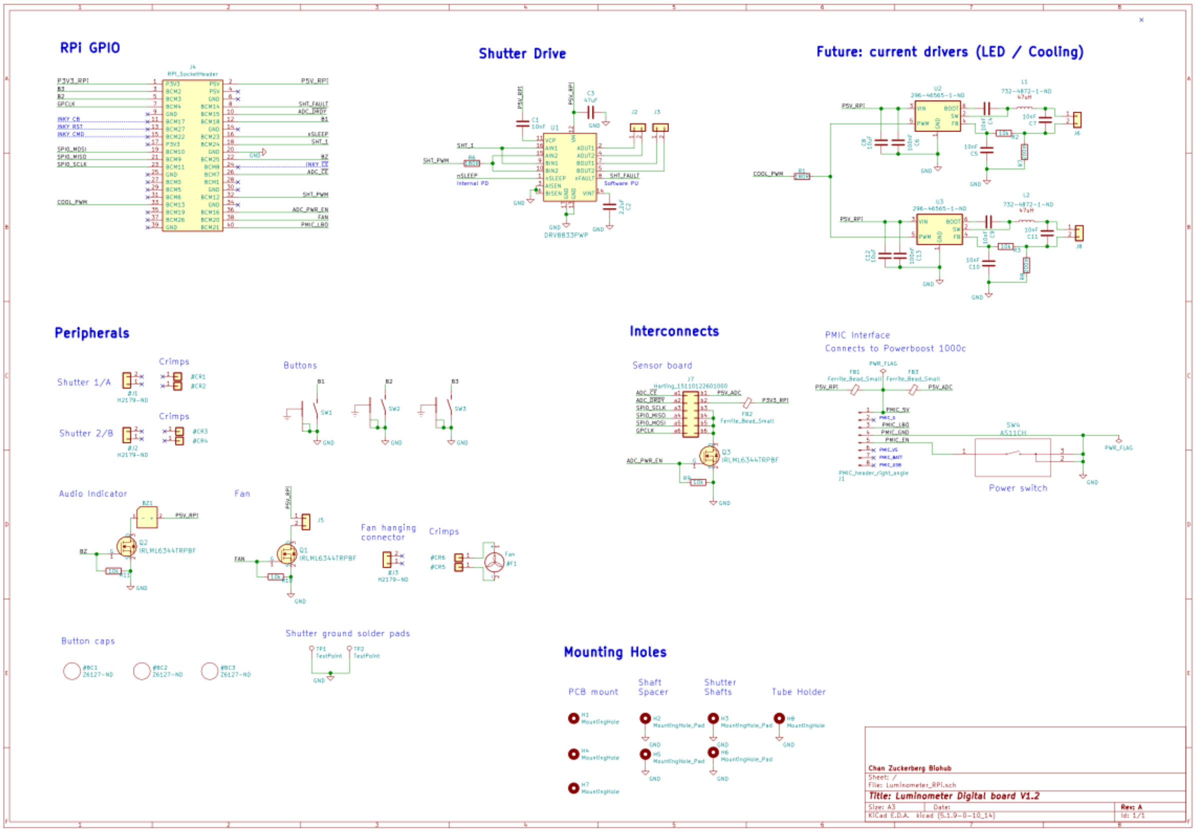


**Supplementary figure 4: Schematic diagram of the digital PCB.** Clockwise, from upper left: Connection to the Raspberry Pi zero GPIO header, with labeled pins. H-Bridge circuit for driving the mechanical shutter system. Unused: Buck converter LED driver chips for potential future usage in fluorescence applications, or sensor temperature stabilization (Peltier). Mechanical interconnects: Board to board interconnect (J7) connecting to the analog sensor board, and jumper cables (J1) to the power management board, and the power switch (SW4). Peripherals include: shutter jumper cables (#J1, #J2), user interface buttons (SW1-3), audio indicator (BZ1), fan connector (J5) and its cable (#J3), button caps (#BC1-2) and ground electrodes for the shutter system (TP1-2). Mounting holes were used for PCB mechanical mounting.

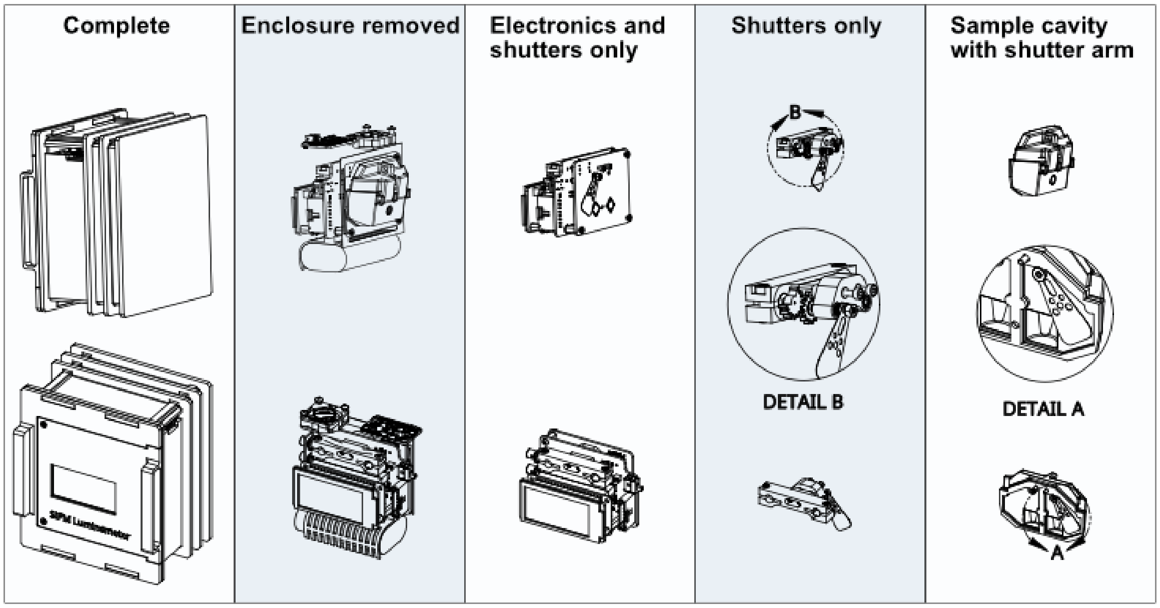


**Supplementary figure 5: Pictorial overview of the luminometer's mechanical design.** The operating principle of the device is illustrated by displaying individual sub-modules. Complete: the completed luminometer is shown from the outside. Enclosure removed: all electronics, tube holder, shutters, power management board, and battery are shown.

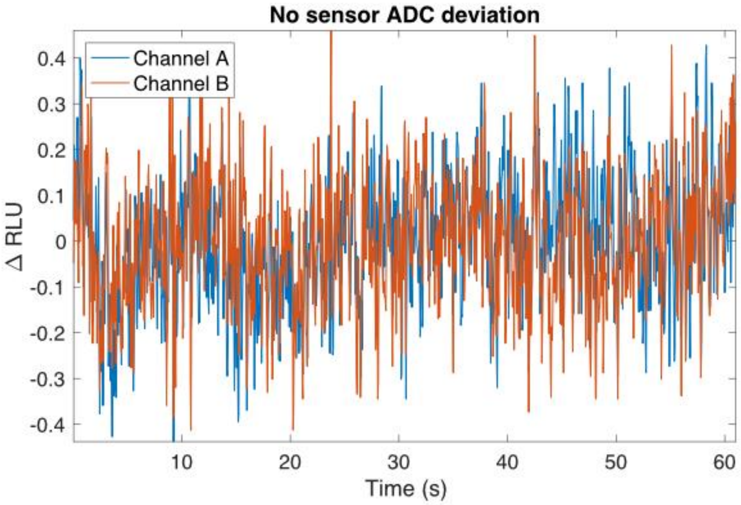


**Supplementary figure 6: The baseline noise of the readout electronics is substantially lower than the limit of detection.** Measurements of the transimpedance amplifier noise were acquired prior to installation of the SiPM sensors, in order to characterize the baseline noise in the absence of sensor current. The resulting traces exhibited a total standard deviation on the order of 0.1 RLU at a sampling rate of 20 Hz. With temporal averaging over a period of 30-60 seconds, this noise is further reduced and is more than 100-fold lower than any expected luminescence signals (> 1 RLU). The ADC discretization limit was 7.2 nV, or 0.003 RLU.

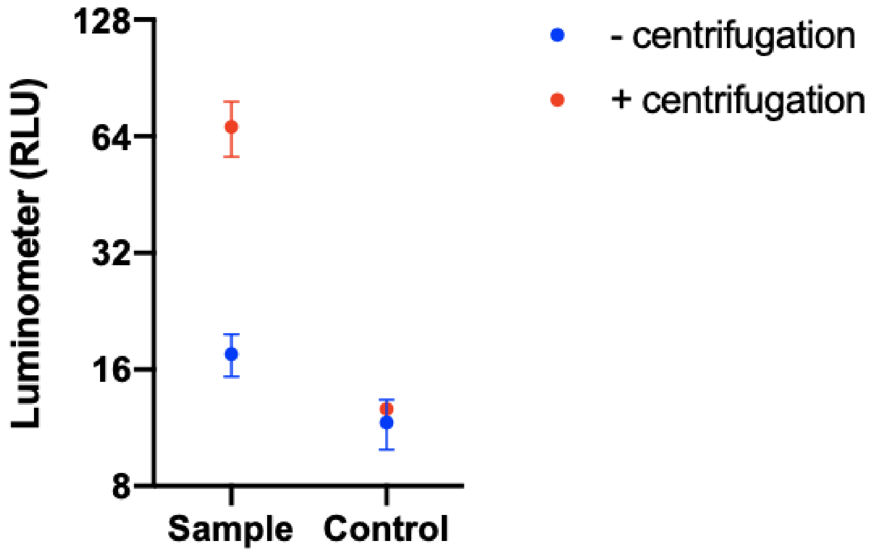


**Supplementary figure 7: Centrifugation of whole blood increases signal in hand-held luminometer.** The spLUC assay was run as previously described and the RLU signal was compared with and without centrifugation of the red cells. Centrifugation increased the signal by 5-fold for this sample from a vaccinated volunteer.

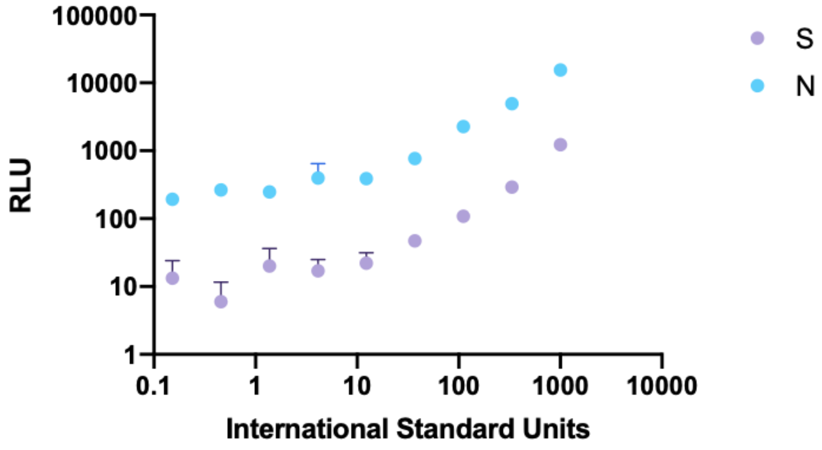


**Supplementary figure 8: Limit of detection of the International Serology Standard on the lab plate reader.** A dilution series of the international serology standard was performed on the

Tecan M200 lab plate-based luminometer. The limit of detection for the IS titration was 37 IS units for the S sensor and 12 IS units for the N sensor

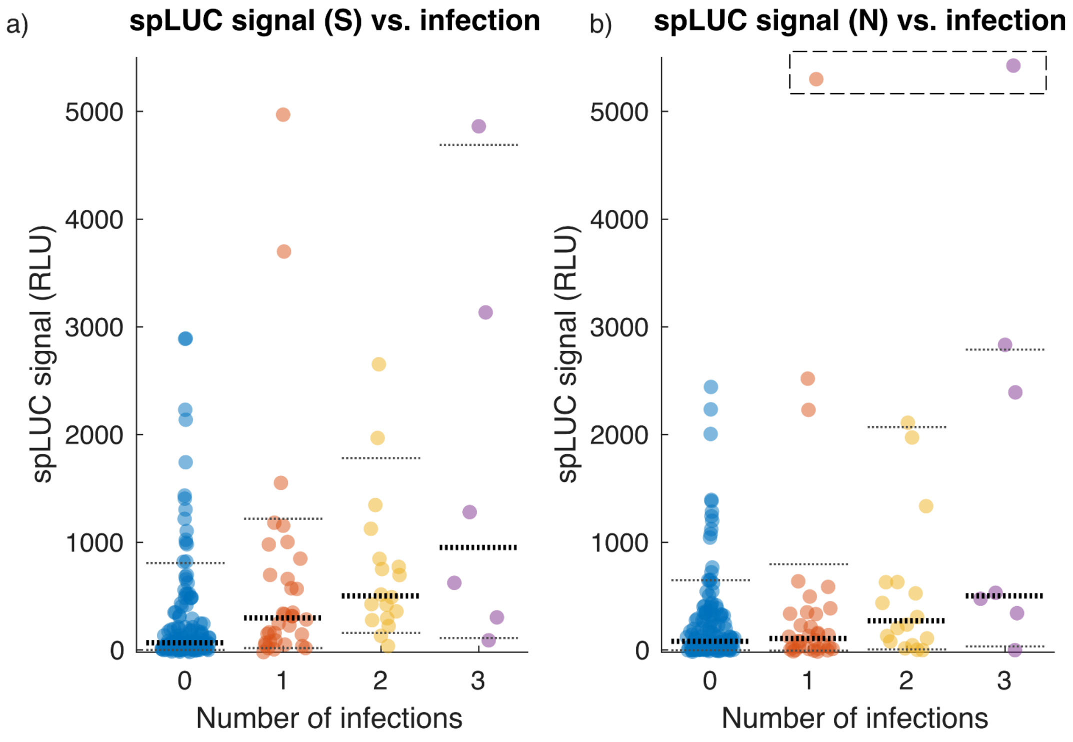


**Supplementary figure 9: Bangladesh spLUC results vs. self-reported infection status**. a) S signal vs. self-reported number of infections. b) N signal vs. self-reported number of infections. In both plots, bold dotted lines denote the median of each distribution, and fine dotted lines denote the 10th and 90th percentiles of each distribution. In b), two outlier data points (the same points from Figure 5c in the main text) were artificially lowered into the display range (dashed box), which was limited to 5500 for clarity.

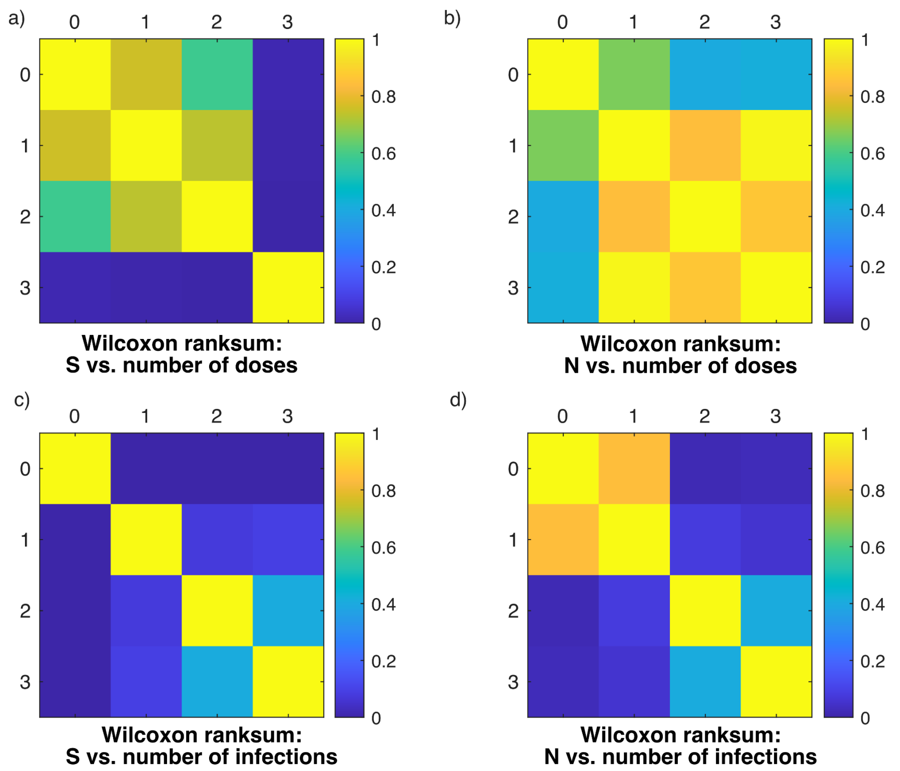


**Supplementary figure 10: Wilcoxon ranksum coefficients comparing spLUC distributions across vaccination and infection status.** Ranksum coefficients denote the P-values of a two-sided Wilcoxon ranksum test for the median similarity of the sampled distributions, where a value of 1.0 indicates an identical distribution median. Diagonal entries in each matrix compare the same data to itself and all have a value of 1.0. Off-diagonal entries compare distributions from sets of participants with different reported numbers of either doses (a and b) or infections (c and d). Coefficients for the S sensor are shown in a) and c), and for the N sensor in b) and d).
